# Integrating genetic regulation and single-cell expression with GWAS prioritizes causal genes and cell types for glaucoma

**DOI:** 10.1101/2022.05.14.22275022

**Authors:** Andrew R. Hamel, Wenjun Yan, John M. Rouhana, Aboozar Monovarfeshani, Xinyi Jiang, Puja A. Mehta, Jayshree Advani, Yuyang Luo, Qingnan Liang, Skanda Rajasundaram, Arushi Shrivastava, Katherine Duchinski, Sreekar Mantena, Jiali Wang, Tavé van Zyl, Louis R. Pasquale, Anand Swaroop, Puya Gharahkhani, Anthony P. Khawaja, Stuart MacGregor, International Glaucoma Genetics Consortium (IGGC), Rui Chen, Veronique Vitart, Joshua R. Sanes, Janey L. Wiggs, Ayellet V. Segrè

## Abstract

Primary open-angle glaucoma (POAG), characterized by retinal ganglion cell death, is a leading cause of irreversible blindness worldwide; however, the molecular and cellular causes are not well understood. Elevated intraocular pressure (IOP) is a major risk factor, but many patients have normal IOP. Colocalization and Mendelian randomization analysis of >240 POAG and IOP GWAS loci and of overlapping eQTLs and sQTLs in 49 GTEx tissues and retina prioritized causal genes for 60% of loci. These genes were enriched in pathways implicated in extracellular matrix organization, cell adhesion, and vascular development. Analysis of single-nucleus RNA-seq of glaucoma-relevant eye tissues revealed that the colocalizing genes and genome-wide POAG and IOP associations were enriched in specific cell types in the aqueous outflow pathways, retina, optic nerve head, peripapillary sclera, and choroid. This study nominated IOP-dependent and independent regulatory mechanisms, genes, and cell types that may contribute to POAG pathogenesis.

## Introduction

Primary open-angle glaucoma (POAG) is the leading cause of irreversible blindness worldwide among people over the age of 55^1^. It is characterized by progressive optic neuropathy, caused by the gradual death of retinal ganglion cells (RGCs) that transmit visual information from the outer retina to the brain via the optic nerve (myelinated RGC axons)^2^. Elevated intraocular pressure (IOP) is a major risk factor for POAG^3^ and is primarily caused by decreased outflow of the aqueous humor from the ocular anterior segment. Decreased outflow may be due to abnormal function of structures in the anterior segment of the eye, consisting of the trabecular meshwork (TM)^4^ and Schlemm’s canal (SC)^5^ in the conventional outflow pathway, and the ciliary muscle and iris in the uveoscleral (unconventional) pathway^6^. However, about one third of patients with POAG display optic nerve degeneration in the absence of abnormally high IOP measurements (normal tension glaucoma (NTG))^7^. Conversely, many people with elevated IOP do not develop glaucoma, suggesting that other processes, including increased RGC susceptibility to normal IOP, might also lead to optic nerve damage. Currently, neuroprotective therapies are lacking, and medications that reduce IOP have limited effectiveness^2^. Gaining a better understanding of the molecular and cellular causes of POAG in the anterior and posterior segments of the eye could suggest novel therapeutic targets.

A recent multi-ethnic genome-wide association study (GWAS) meta-analysis of 34,179 POAG cases and 349,321 controls of European, Asian, and African ancestries identified 127 risk loci associated with POAG^8^, explaining ∼9% of POAG heritability, and a meta-analysis of the European subset identified 68 POAG loci^8^, some of which were not uncovered in the cross-ancestry meta-analysis. Furthermore, a GWAS meta-analysis of IOP performed on 139,555 individuals, primarily of European descendant^9^, has identified 133 independent associations in 112 loci, largely overlapping with two other studies^10, 11^. The IOP variants’ effect sizes and direction of effect are highly correlated with their effect on POAG risk^8, 9^, and together they explain 9-17% of IOP heritability. Vertical-cup-to-disc ratio (VCDR), central corneal thickness, and corneal hysteresis, a measure of the viscoelastic damping of the cornea, have also been associated with POAG risk, and large GWAS meta-analyses have uncovered 70-200 genetic associations for these traits^12–23^.

Identifying putative causal genes and cell types underlying the genetic associations with POAG and its related traits is challenging. As with other complex traits, a majority of associated variants lie in noncoding regions and are enriched for regulatory effects^24–26^. Due to linkage disequilibrium (LD), the discovered associations typically tag multiple variants and genes, making it hard to pinpoint the implicated causal gene(s) from sequence alone. Furthermore, genetic regulatory effects in relevant ocular tissues are limited, reported to date only in retinal tissues^27–30^, and have not yet been detected at cellular resolution in other parts of the eye. Nevertheless, through single-cell or single-nucleus RNA-sequencing (sc/snRNA-seq), human cell atlases and cellular level transcriptomes have been generated for various non-diseased eye tissues relevant to POAG pathogenesis, including retina^31–33^, the aqueous humor outflow pathways^34, 35^, six tissues in the anterior chamber^36^, and the optic nerve head (ONH), where RGCs pass to exit the eye, the optic nerve, and surrounding posterior tissues^37^. Using a method we recently developed, ECLIPSER^38, 39^, we show that cell type-specific enrichment of genes mapped to GWAS loci of complex diseases and traits can help identify cell types of action for diseases in relevant tissues^38, 39^.

In this study, we have combined expression quantitative trait loci (eQTLs) and splicing QTLs (sQTLs) in 49 (non-ocular) tissues from the Genotype-Tissue Expression (GTEx) Project^26^, retinal eQTLs^31, 32^, retinal Hi-C data^40^, and single-cell expression from glaucoma-relevant eye tissues^33, 36, 37^ with POAG and IOP genetic associations to identify regulatory mechanisms, genes, pathways, and cell types that may play an important role in POAG etiology.

## Results

An overview of the analytical steps and approaches taken are described in Fig. 1 and Supplementary Note.

**Figure 1.**
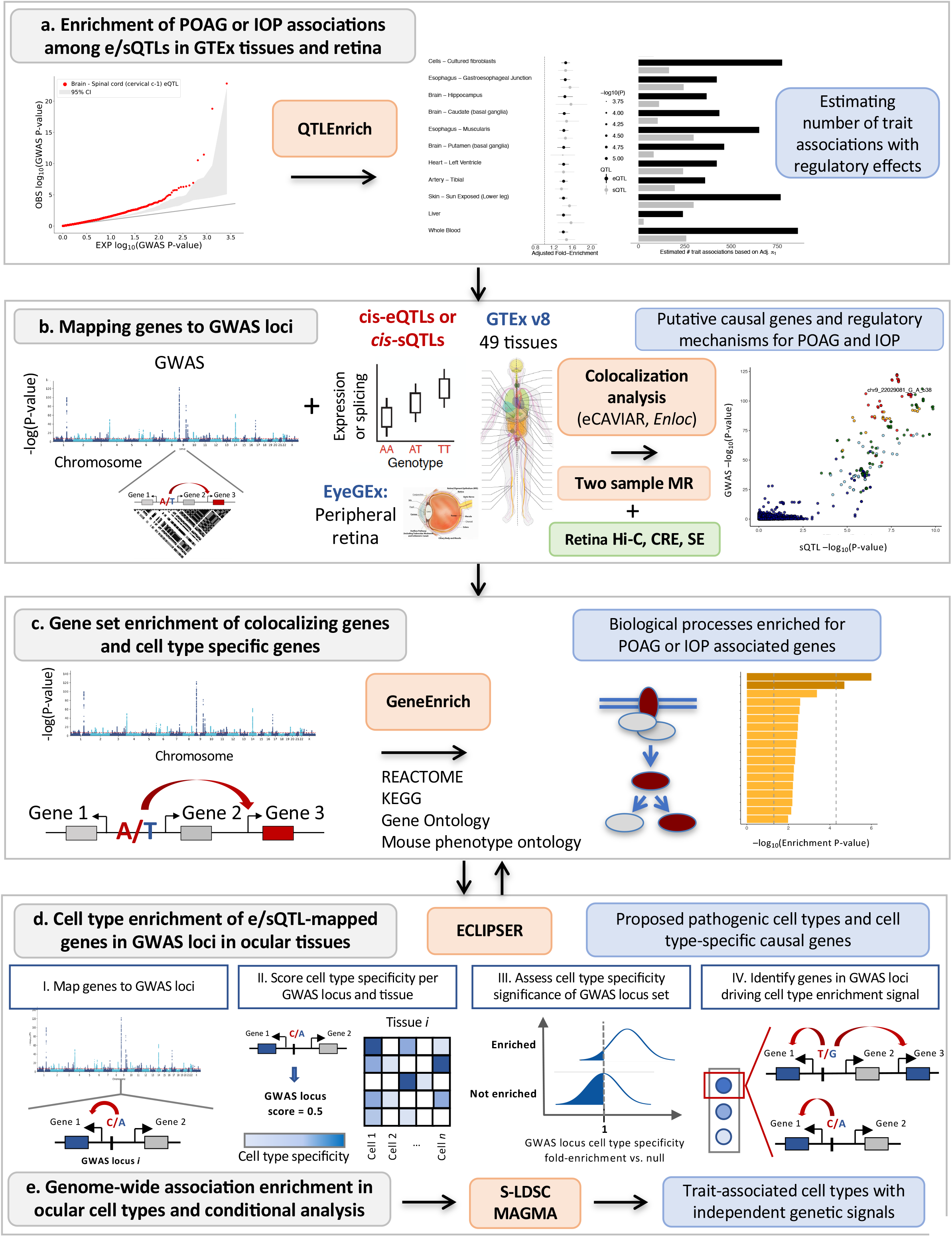
Analysis workflow from POAG and IOP GWAS to causal regulatory mechanisms, genes, pathways, and cell types. **a**, POAG and IOP associations genome-wide (known and modest associations) were tested for enrichment among e/sQTLs in GTEx tissues and retina compared to permuted null sets of variants matched on confounding factors, using *QTLEnrich*. In cases where enrichment was found, the lower bound number of e/sQTLs in a given tissue, likely to be true trait associations was estimated using an empirically derived, true positive rate (1) approach. **b,** Putative causal genes were prioritized per known POAG and IOP GWAS locus by applying two colocalization methods to all e/sQTLs from 49 GTEx tissues and retina eQTLs that overlapped each locus. A Manhattan plot of the POAG cross-ancestry GWAS meta-analysis was plotted with QMplot (URLs). **c,** All target genes of significantly colocalizing e/sQTLs per trait were tested for enrichment in signaling and metabolic pathways (Reactome, KEGG), gene ontologies and mouse phenotype ontologies using *GeneEnrich*. **d,** Significantly colocalizing e/sGenes were tested for enrichment in specific cell types in single nucleus RNA-seq data of glaucoma-relevant eye tissues, using ECLIPSER. Cell type specific genes were defined with cell type fold-change >1.3 and FDR<0.1 per tissue. Cell type specificity significance per GWAS locus set for a given trait was assessed against a null distribution of GWAS loci associated with unrelated, non-ocular traits, using a Bayesian Fisher’s exact test. Genes mapped to GWAS loci with a cell type specificity score above the 95th percentile of null locus scores were proposed as contributing to the trait in the enriched cell type. **e,** Cell type enrichment for the POAG and IOP GWAS was corroborated using two regression-based methods that assess cell type specificity of trait associations considering all associations genome-wide: stratified-LD score regression and MAGMA.

### POAG and IOP associations enriched among eQTLs and sQTLs

To assess the relevance of eQTLs and sQTLs to POAG risk and IOP variation, we tested whether *cis*-eQTLs and *cis*-sQTLs (e/sQTLs) from 49 GTEx (v8) tissues^26^ and peripheral retina *cis*-eQTLs^27, 28^ were enriched for POAG or IOP associations (GWAS P<0.05) using *QTLEnrich*^24, 26^ that adjusts for confounding factors and tissue sample size (Methods and Fig. 1a). We found significant enrichment of multiple POAG and IOP associations (both genome-wide significant and subthreshold) among eQTLs and sQTLs in most of the 49 GTEx tissues and in retina (Bonferroni-corrected P<5×10^-4^) (Fig. 2a, b and Supplementary Tables 1-3). Many of the top enriched GTEx tissues contain cell types that may be pathogenic to glaucoma (Supplementary Note). The relative contribution of sQTLs to POAG and IOP, as measured by adjusted fold-enrichment and estimated true positive rate, was larger than the relative contribution of eQTLs to these traits (One-sided Wilcoxon rank sum test P<1.5×10^-11^ and P<0.03, respectively; Supplementary Fig. 1a-c, and Supplementary Tables 1-3), as observed with other complex traits^26^. The absolute number of eQTLs proposed to contribute to POAG and IOP (average 258 to 606 per tissue) was 2-fold larger than that of sQTLs (average 124 to 320 per tissue), likely due to the larger discovery rate of eQTLs compared to sQTLs^26^ (Supplementary Fig. 1a-c and Supplementary Table 1). The target genes of eQTLs or sQTLs with top-ranked POAG or IOP GWAS p-values (P<0.05) were enriched in metabolic and cellular processes (Methods; Supplementary Tables 4-5; Supplementary Note).

**Figure 2.**
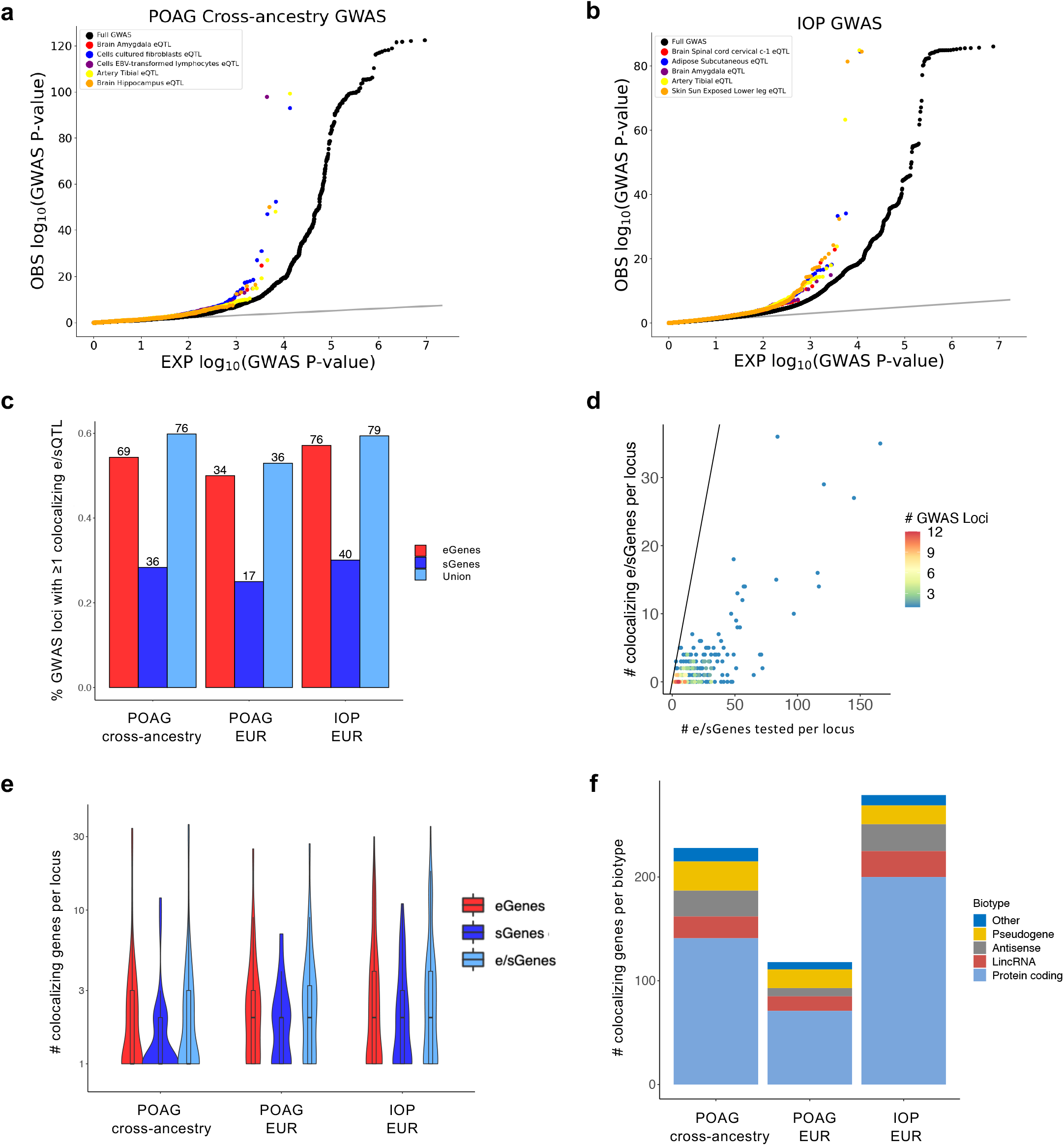
Enrichment and colocalization analysis of eQTLs and sQTLs with POAG and IOP associations. **a, b** Quantile-quantile (Q-Q) plots of POAG cross-ancestry (**a**) and IOP (**b**) GWAS -log_10_ (P-value) compared to expectation for the best eQTL per eGene sets (eVariants with FDR < 0.05) of the top enriched tissues based on adjusted fold-enrichment (colored points), compared to all variants in the GWAS (black points). Grey line represents the diagonal. **c**, Histogram of percent of GWAS loci with at least one colocalizing e/sQTL (eCAVIAR CLPP > 0.01 and/or *enloc* RCP > 0.1) for the three traits (POAG cross-ancestry, POAG European (EUR) ancestry subset, and IOP European ancestry). Numbers above the bars represent the number of loci with at least one colocalizing e/sQTL. Red, dark blue, and light blue bars indicate percentage of loci with at least one colocalizing eGene, sGene, or both, respectively. **d**, Scatter plot comparing unique number of e/sGenes per locus that significantly colocalized per locus versus unique number of e/sGenes tested per locus. Points are color-coded by number of GWAS loci. The black line represents the diagonal. **e**, Violin plots showing the distribution of the unique number of colocalizing eGenes (red), sGenes (dark blue), or both (light blue) per locus for the three GWAS tested. The center line in the box plots contained within each violin plot shows the median and the box edges depict the interquartile range. **f**, Stacked histogram showing the number of colocalizing e/sGenes per gene biotype for each GWAS. Protein coding (light blue), lincRNA (brown), antisense (grey), pseudogenes (yellow), and other (dark blue).

### Colocalization analysis of POAG and IOP GWAS loci with *cis-*e/sQTLs

Given the widespread e/sQTL enrichment of POAG and IOP associations, we used the e/sQTLs in all 49 GTEx tissues and retina eQTLs to propose putative causal genes that may underlie genome-wide significant loci for these traits. We applied two colocalization methods, eCAVIAR^41^ and *enloc*^42^, to 127 POAG loci from a large cross-ancestry GWAS meta-analysis^8^, 68 POAG loci from a European (EUR) subset meta-analysis^8^ (POAG EUR), and 133 IOP loci from a primarily European GWAS meta-analysis^9^ (IOP) (variant list in Supplementary Table 6; Methods and Fig. 1a), and any e/sQTLs that overlapped each GWAS locus LD interval (Methods). The results are presented per trait and colocalization method in Supplementary Tables 7-12 and summarized in Supplementary Table 13. We defined a “comprehensive set” of putative causal genes and regulatory mechanisms for POAG and IOP as those e/sGenes that were significant with at least one of the colocalization methods (Colocalization posterior probability (CLPP) > 0.01 for eCAVIAR, and regional colocalization probability (RCP) > 0.1 for *enloc* (see Methods); Supplementary Table 13), filtering out potential false positives (See Methods and examples in Supplementary Fig. 3). The largest number of colocalizing e/sGenes was found in tibial nerve, adipose, skin, artery, and fibroblasts, among other tissues, many of which contain cell types relevant to the pathogenicity of glaucoma (Supplementary Fig. 4). Eighteen retina eQTLs colocalized with 13 POAG and/or IOP loci (Column AH in Supplementary Table 13). The number of significantly colocalizing e/sGenes per tissue significantly correlated with tissue sample size (Pearson’s R^2^=0.72, P=1×10^-14^, Supplementary Fig. 4) that is also associated with the number of detected e/sQTLs per tissue^26^. This suggests that e/sQTL discovery power is a driving factor in tissue identity of the colocalizing e/sQTLs. We therefore primarily considered the causal genes proposed by the colocalization analysis and not the associated tissues, in downstream analyses.

We found that 58% of all GWAS loci tested significantly colocalized with at least one eQTL and/or sQTL based on eCAVIAR and/or *enloc*: 60% (76) of 127 cross-ancestry POAG GWAS loci, 53% (36) of 68 European POAG loci and 59% (79) of 133 IOP loci (Fig. 2c and Supplementary Table 14). About 55% and 29% of GWAS loci colocalized with ≥1 eQTL and ≥1 sQTL, respectively. For 21% of the POAG and IOP GWAS loci (69 loci total), significant colocalization was found for the same e/sGene with eCAVIAR and *enloc* (‘high confidence set’ listed in Table 1 and Supplementary Table 15). The GWAS-e/sQTL colocalization analysis significantly reduced the number of putative causal genes per GWAS locus for POAG and IOP from an average of 22.8 ± 1.8 genes tested per LD interval (range: 3-166, median=15) to an average of 3.5 ± 0.4 genes per locus (range: 1-36, median of 1 or 2 genes per locus per trait; Fig. 2d,e and Supplementary Table 16). eQTLs and sQTLs nominated an average of 3 and 2 causal genes per locus, respectively, with partial overlap of target genes between the colocalizing eQTLs and sQTLs (Fig. 2e and Supplementary Table 16). 60-72% of the colocalizing e/sGenes per trait were protein-coding and 18-20% were noncoding RNA genes, half of which were lincRNAs and half antisense genes (Fig. 2f, Supplementary Tables 17-18, and Supplementary Note). A single causal gene was proposed for 80 (42%) of the POAG and IOP loci with significantly colocalizing e/sQTLs (Supplementary Tables 19-20), 49 (61%) of which are the nearest gene to the lead GWAS variant. In 30.3% (23/76), 16.7% (6/36) and 31.7% (25/79) of the POAG cross-ancestry, POAG EUR and IOP GWAS loci, respectively, with significant colocalization results, the colocalizing e/sGenes were not the nearest gene to the lead GWAS variant. In total, 228, 118, and 279 genes, including previously suggested and novel ones, are candidate causal genes for POAG cross-ancestry risk, POAG EUR risk, and IOP variation, respectively (Supplementary Table 21), with a total of 459 genes proposed from the combined datasets.

**Table 1.**
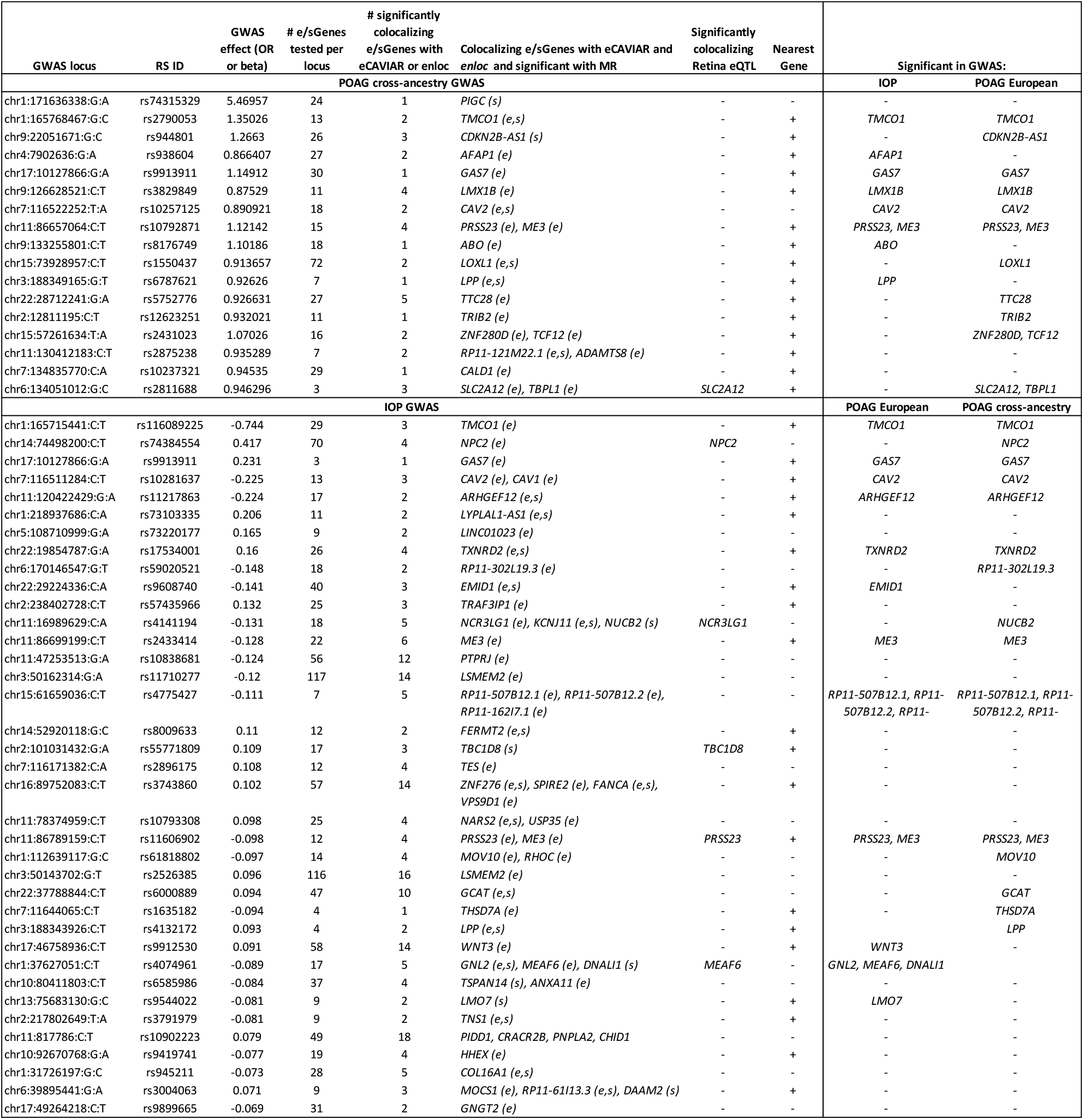
List of high-confidence colocalizing expression and splicing QTLs with POAG and IOP GWAS loci.

### Colocalizing e/sGenes of top POAG and IOP GWAS loci and direction of regulatory effect on disease risk

In addition to prioritizing causal genes and regulatory mechanisms that may contribute to POAG and IOP, colocalizing e/sQTLs propose the direction of effect of altered gene expression or splicing on disease risk or trait variation (examples for top POAG and IOP GWAS signals in Fig. 3 and Supplementary Fig. 5). For example, an eQTL and sQTL acting on *TMCO1* and an eQTL acting on *TMCO1’*s antisense, *RP11-466F5.8*, in the opposite direction, colocalized with the second strongest association with POAG (rs2790053, odds ratio (OR)=1.35; CLPP=0.92-1) and the top IOP association (rs116089225, beta = -0.744; CLPP=0.87-1) (Fig. 4a-e and Supplementary Fig. 6). Decreased expression of *TMCO1* and increased expression of *RP11-466F5.8* are proposed to lead to increased IOP levels and increased POAG risk (Fig. 4, Supplementary Fig. 6, and Supplementary Table 13). Furthermore, an alternative splice donor site in exon 4, the first exon in the *TMCO1* mRNA in GTEx Cells Cultured fibroblasts, leads to a longer exon 4 (Fig. 4f,g) that is associated with decreased POAG risk (Supplementary Tables 7 and 13). *TMCO1* is expressed in different cell types in the anterior and posterior parts of the eye, including lymphatic and fibroblast cells in the conventional and unconventional outflow pathways, vascular and immune cells in the anterior and posterior segments, and macroglial cells in the retina (Supplementary Figure 7). Other examples include *ANGPT1* and *ANGPT2*, involved in vascular biology, whose increased expression is proposed to reduce IOP levels (Supplementary Fig. 8 and Supplementary Table 13) that is consistent with the effect observed on IOP in *Angpt1*-knockout mice^43^.

**Figure 3.**
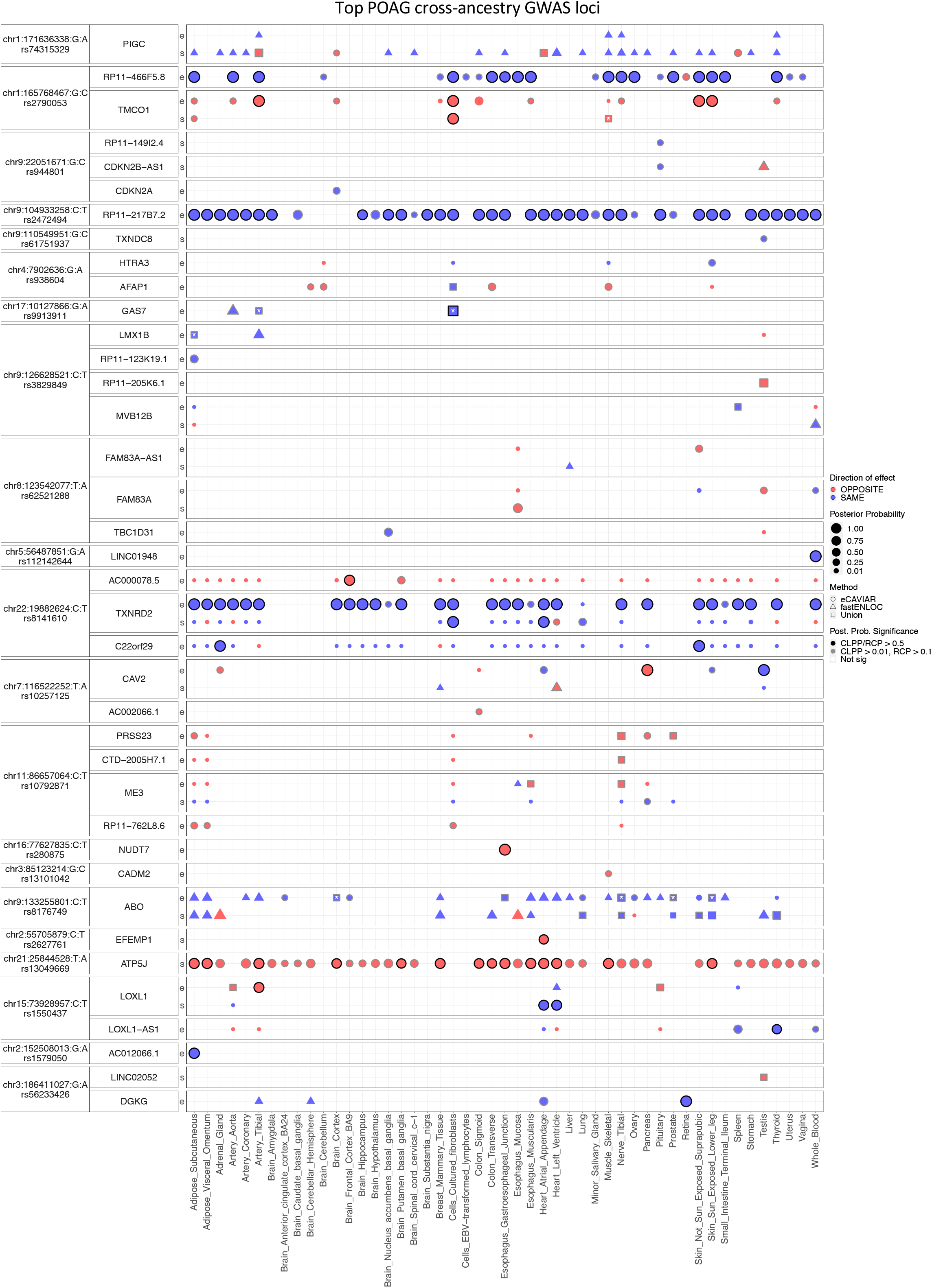
Colocalizing e/sQTLs in GTEx tissues and retina with top POAG GWAS loci. Genes with at least one significant colocalization result are shown for e/sQTLs tested across 49 GTEx tissues and peripheral retina for the top 21 POAG cross-ancestry GWAS loci. GWAS loci were ordered by absolute value of their effect size. Within each locus, genes were ordered based on their chromosome position. Bubble size is proportional to the maximum colocalization posterior probability of all e/sVariants tested for the given gene, QTL type and tissue combination. Points are color-coded by direction of effect (blue if increased expression or splicing increases POAG risk or vice versa; red if increased expression or splicing decreases POAG risk or vice versa). Shape of points indicates colocalization method used: circle (eCAVIAR), triangle (*enloc*), and square (tested in both methods; results shown for method with maximum posterior probability). Grey or black border denotes variant-gene-tissue-QTL combination that passed QC filtering (Methods) and a colocalization posterior probability cutoff above 0.01/0.1 (CLPP/RCP) or 0.5 (higher confidence), respectively. White or black asterisk in the square indicates whether the second method tested passed a posterior probability cutoff 0.01/0.1 (CLPP/RCP) or 0.5, respectively.

**Figure 4.**
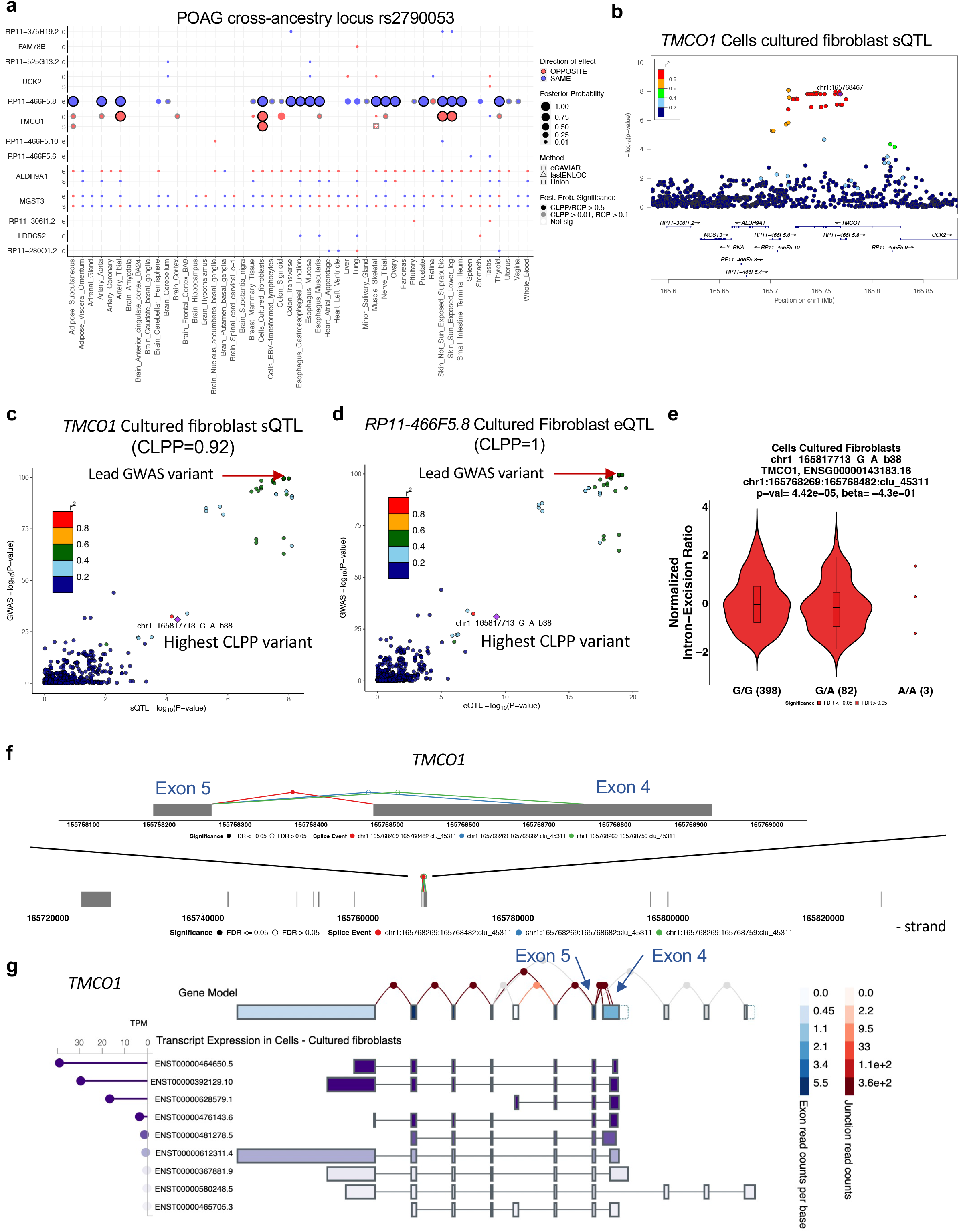
Examples of colocalizing e/sQTLs with top POAG and IOP GWAS loci. **a**, Colocalization results for all e/sGenes tested in the POAG cross-ancestry rs2790053 locus LD interval with 21 significant eQTL or sQTL result across 49 GTEx tissues and peripheral retina based on eCAVIAR and/or *enloc*. Genes were ordered by chromosome position. Size of points is proportional to the maximum colocalization posterior probability of all e/sVariants tested for the given gene, QTL type and tissue combination. Points are color-coded by direction of effect (blue if increased expression or splicing increases POAG risk or vice versa; red if increased expression or splicing decreases POAG risk or vice versa). Shape of points indicates colocalization method: circle (eCAVIAR), triangle (*enloc*), and square (tested in both methods; results shown for method with maximum posterior probability). Grey or black border denote variant-gene-tissue-QTL combination that passed QC filtering (Methods) and a colocalization posterior probability cutoff above 0.01/0.1 (CLPP/RCP) or 0.5, respectively. White or black asterisk in the square indicates whether the second method tested passed a posterior probability cutoff of 0.01/0.1 (CLPP/RCP) or 0.5, respectively. **b**, LocusZoom^96^ plot for *TMCO1* sQTL -log_10_(P-value) in GTEx Cells Cultured fibroblasts in the POAG cross-ancestry GWAS variant rs2790053 (chr1_165768467_C_G_b38) LD interval. Points are color-coded by LD (r^2^) relative to the lead GWAS variant. **c**,**d** LocusCompare plot of -log_10_(P-value) of the POAG cross-ancestry GWAS meta-analysis versus the -log_10_(P-value) of the Cells Cultured fibroblast sQTL or eQTL acting on *TMCO1* (**c**) or *RP11-466F5.8* (**d**), respectively. Points are color-coded based on LD (r^2^) relative to the variant with the highest eCAVIAR colocalization posterior probability (CLPP). **e,** Violin plot of normalized intron-excision ratio for chr1:165768269-165768482 computed with Leafcutter^95^ for *TMCO1* in Cells Cultured fibroblasts as a function of the genotype of the sVariant chr1_165817713_G_A_b38 (rs143863391) with the highest CLPP (0.92) for this sQTL and the POAG cross-ancestry GWAS locus rs2790053. The effect size of the sQTL relative to the alternative allele (*beta* = -0.43) is in opposite direction relative to the GWAS variant (*beta* = 0.25), suggesting that decreased splicing between chr1:165768269-165768482 increases POAG risk. **f,** Gene model for *TMCO1* in GTEx fibroblasts on the negative strand showing all intron excision splicing events detected with Leafcutter in *TMCO1*. A zoom in of the splicing event (chr1:165768269-165768482; red), an alternative splice donor site on exon 4 (red versus blue or green), associated with the sVariant chr1_165817713_G_A_b38 that colocalized with POAG risk is shown. A longer exon 4 in *TMCO1* is associated with decreased risk of POAG. **g,** *TMCO1* gene model and transcripts expressed in Cells Cultured fibroblasts taken from the GTEx portal (URLs). In the gene model, exon boxes are color-coded by exon read counts per base (blue) and lines connecting exons are color-coded by exon-exon junction read counts (red). All splicing events observed in the tissue are shown, including the alternative splice event between exon 4 and exon 5 whose genetic regulation colocalized with POAG (**c**). Below the gene model, transcript expression in Cells Cultured fibroblasts in Transcripts per Million (TPM), computed with RSEM^110^, is shown in descending order.

### Colocalizing genes for shared and distinct POAG and IOP loci

Of 50 overlapping POAG and IOP GWAS loci, 39 (78%) of the loci had at least one significant colocalization result for both traits, and in all cases at least one common gene was implicated (Supplementary Table 13 and Table 1). In most of the cases (95%), the relative direction of effect of the colocalizing e/sQTLs on IOP was consistent with IOP’s effect on POAG risk, proposing IOP-dependent mechanisms for POAG risk. For example, decreased *GAS7* expression or increased *ABO* expression were associated with both increased IOP levels and increased POAG risk (Supplementary Tables 7-13 and Supplementary Figs. 9-10). e/sGenes that colocalize with POAG loci not associated with IOP (48 loci; Column N in Supplementary Table 13) may suggest IOP-independent mechanisms.

To prioritize regulatory variants and genes that may affect POAG independent of IOP, we integrated retina Hi-C loops and/or epigenetically derived *cis*-regulatory elements (CREs) and super-enhancers (SEs) (Supplemental Data 4 in Marchal *et al*.^40^) with the POAG-only loci (Methods). In 17 of the loci, 21 colocalizing e/sQTLs was supported as a potential causal mechanism by retina Hi-C loops (6 loci), CREs (16 loci) and/or SEs (5 loci) (Supplementary Table 22). This includes the strongest normal tension glaucoma (NTG) association (9p21)^8,44^ in the POAG cross-ancestry (rs944801 OR=1.26) and POAG EUR (rs6475604 OR=1.3) GWAS meta-analyses that colocalized with a *CDKN2A* eQTL in brain cortex, *CDKN2B-AS1 and RP11-149I2.4* sQTLs in pituitary, and *CDKN2B* eQTL in skeletal muscle (Supplementary Table 13). The POAG risk variants and colocalizing e/sQTLs in this locus overlapped retinal CREs (Fig. 5a). These results imply that increased expression of *CDKN2A,* decreased expression of *CDKN2B*, and exon skipping in *CDKN2B-AS1* may increase POAG risk (Fig. 3, Supplementary Figs. 5a and 11). Other examples, involving a retina *SLC2A12* eQTL overlapping a retinal CRE, and e/sQTLs acting on *RERE* and its antisense, *RERE-AS1*, that are physically linked via retina chromatin loops to the *RERE* transcription start site (TSS), are shown in Fig. 5b and c (see also Supplementary Table 22). Notably, *RERE* expression is enriched in oligodendrocytes in the optic nerve head and optic nerve (False discovery rate (FDR)=0.07) and in retinal pigment epithelium (RPE) and S-cones in the macula (FDR=0.06), as shown below (Supplementary Table 38; Supplementary Fig. 18f, g).

**Figure 5.**
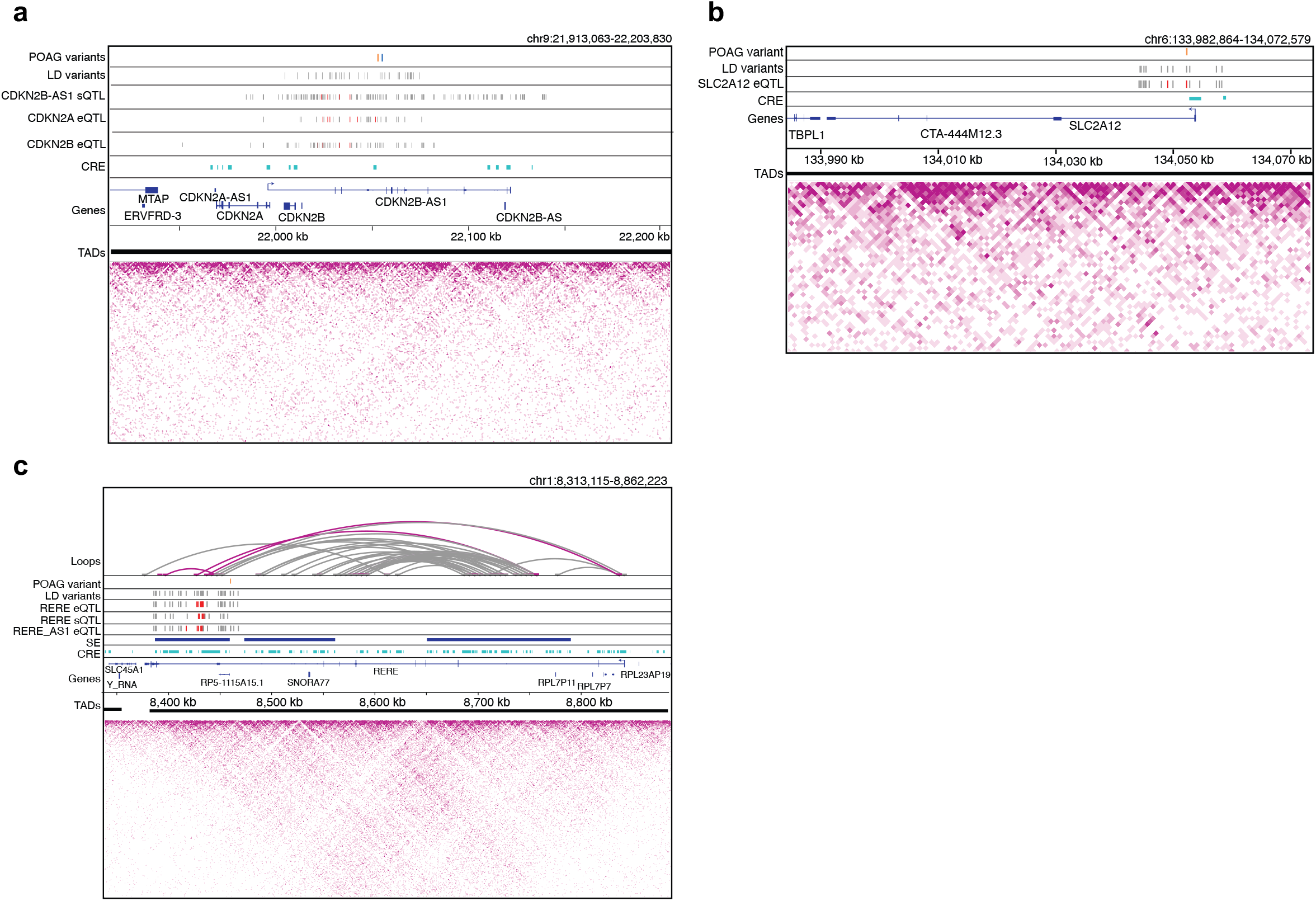
Chromatin loops and regulatory elements in human retina support effect of colocalizing e/sQTLs on POAG risk. **a**, Retina CREs (cyan) derived from epigenetic data overlapping e/sVariants that colocalized with POAG associations in the *CDKN2A/B* locus. The lead POAG variants from the cross-ancestry (blue line) and European subset (orange line) GWAS are shown in the top track, followed by their linkage disequilibrium (LD) proxy variants (r^2^>0.8) in the track below. The significantly colocalizing *CDKN2B-AS1* sVariants in Pituitary, *CDKN2A* eVariants in Brain Cortex, and *CDKN2B* eVariants in Skeletal Muscle are represented by red lines, and the grey lines represent LD proxy variants that are also significant e/sQTLs (FDR<0.05) for the corresponding gene and tissue. **b,** Retina CREs (cyan) overlapping retina *SLC2A12 e*Variants that colocalized with the POAG cross-ancestry association. Tracks display the lead POAG cross-ancestry GWAS variant (orange) and its LD proxy variants (grey), followed by the significantly colocalizing *SLC2A12* retina eVariants (red) with their LD proxy eVariants that are also significant eQTLs at FDR<0.05 (grey). The CRE overlaps the promoter of *SLC2A12.* **c,** Retina chromatin loops from Hi-C data, SEs (blue), and CREs (cyan) shown for the *RERE* POAG locus. Tracks display the lead POAG cross ancestry GWAS variant (orange) and its LD proxy variants (grey), followed by significantly colocalizing *RERE* eVariants in Nerve Tibial, *RERE* sVariants in fibroblast cells, and *RERE-AS1* eVariants in Adipose Subcutaneous (red), and their LD proxy variants that are also significant e/sQTLs (FDR<0.05) for the corresponding gene and tissue (grey). Magenta loops have one foot that overlaps or is in LD with the POAG variant and colocalizing e/sQTLs. In all panels, LD proxy variants were computed at r^2^>0.8, TADs are represented as solid black lines, and the magenta heatmaps represent Hi-C physical contact maps. CRE, Cis-regulatory element; SE, Super-enhancer; TAD, Topologically associating domain.

### Colocalizing genes in population-specific and cross-ancestry POAG loci

For all 59 POAG EUR loci also found in the POAG cross-ancestry meta-analysis, at least one common colocalizing e/sGene was found for both the EUR and cross-ancestry GWAS (Supplementary Table 13). One such example is *EFEMP1*, in which rare mutations have been associated with a Mendelian form of glaucoma^45^. Colocalization analysis suggests that skipping of exons 6 and 7 in *EFEMP1* may be protective for POAG (Supplementary Fig. 12). Of the 9 loci found only in the POAG EUR GWAS, two loci colocalized with eQTLs acting on several genes each, including genes involved in the extracellular matrix (*EMID1*) and vascular endothelial growth (*ANGPTL2*), respectively, both of which also colocalized with IOP (Supplementary Table 23). In addition, three associations in the POAG cross-ancestry meta-analysis demonstrated significant allelic heterogeneity among the three populations (European, East Asian, and African American)^8^. e/sQTL colocalized with two of these loci. One is the 9p21 locus rs944801 with *CDKN2A/B* described above, which was significant in the European and Asian populations, but not the African population (P_heterogeneity=1.5 x 10^-8^) in which it is in lower frequency (AFR MAF=0.073, EUR MAF=0.42, EAS MAF=0.11 in gnomAD (URLs)). The other is a European-specific locus with the largest POAG odds ratio (rs74315329, OR=5.47). This variant is a nonsense mutation (p.Gln368Ter) in *MYOC*, known to cause juvenile-onset and adult-onset open-angle glaucoma with dominant inheritance^46^. This variant is 10-fold more common in the European population compared to the African population and is not found in the East Asian population (gnomAD; URLs). Of all the e/sQTL gene-tissue pairs that overlapped this locus targeting 24 genes, we identified an sQTL acting on *PIGC,* phosphatidylinositol glycan anchor biosynthesis class C, that significantly colocalized with the POAG cross-ancestry locus (spleen CLPP=0.12, and arterial tissues RCP=0.26-0.34; Supplementary Table 13 and Supplementary Fig. 13). Conditioning on the nonsense variant in *MYOC* that is likely the primary causal variant in the locus, we found that a secondary haplotype colocalized with the *PIGC* sQTL (Supplementary Tables 24-28; more details in Supplementary Note). These results suggest that decreased exon 2 skipping in *PIGC* or increased *PIGC* expression may lead to increased POAG risk (Supplementary Fig. 13). *PIGC* is among 10 colocalizing POAG genes enriched in oligodendrocytes in the optic nerve head and optic nerve (See below; FDR=0.07, Supplementary Table 38; Supplementary Fig. 18f, g).

### Mendelian randomization (MR) of colocalizing e/sQTLs

To provide additional support for a causal relationship between e/sQTLs and POAG and/or IOP, we applied two-sample MR to all the significantly colocalizing e/sQTL and GWAS locus pairs based on eCAVIAR and/or *enloc* (Methods). We found evidence for a causal relationship (FDR < 0.05) for 348 (75%) genes that were robust to the influence of horizontal pleiotropy, where pleiotropy-robust sensitivity could be performed (Supplementary Table 29). A high-confidence list of putative POAG and/or IOP causal genes based on colocalization analysis and MR is provided in Table 1. We found 239 e/sGenes to have significant MR associations with both POAG and IOP, including *TMCO1, GAS7,* and *LMX1B,* which colocalized with the largest association signals for both POAG and IOP GWAS loci (Supplementary Figs. 5, 6, 9), and *DGKG* and *NPC2*, whose retina eQTLs colocalized with POAG and IOP. Sixty-eight genes had significant MR associations with IOP but not POAG, such as *HLA-B* and *SLC7A6*, and 41 genes had significant MR associations with POAG but not IOP, such as *CDKN2B-AS1, RERE, and YAP1*, proposing high-confidence IOP-independent mechanisms (Supplementary Table 29 and Fig. 5). Since the MR analysis could not be applied to the larger, better-powered POAG cross-ancestry GWAS, as it requires a similar population background between the e/sQTL and GWAS studies (European in our case), our downstream analyses were applied to the more inclusive list of proposed causal genes based on the colocalization analysis (Supplementary Table 13).

**Figure 6.**
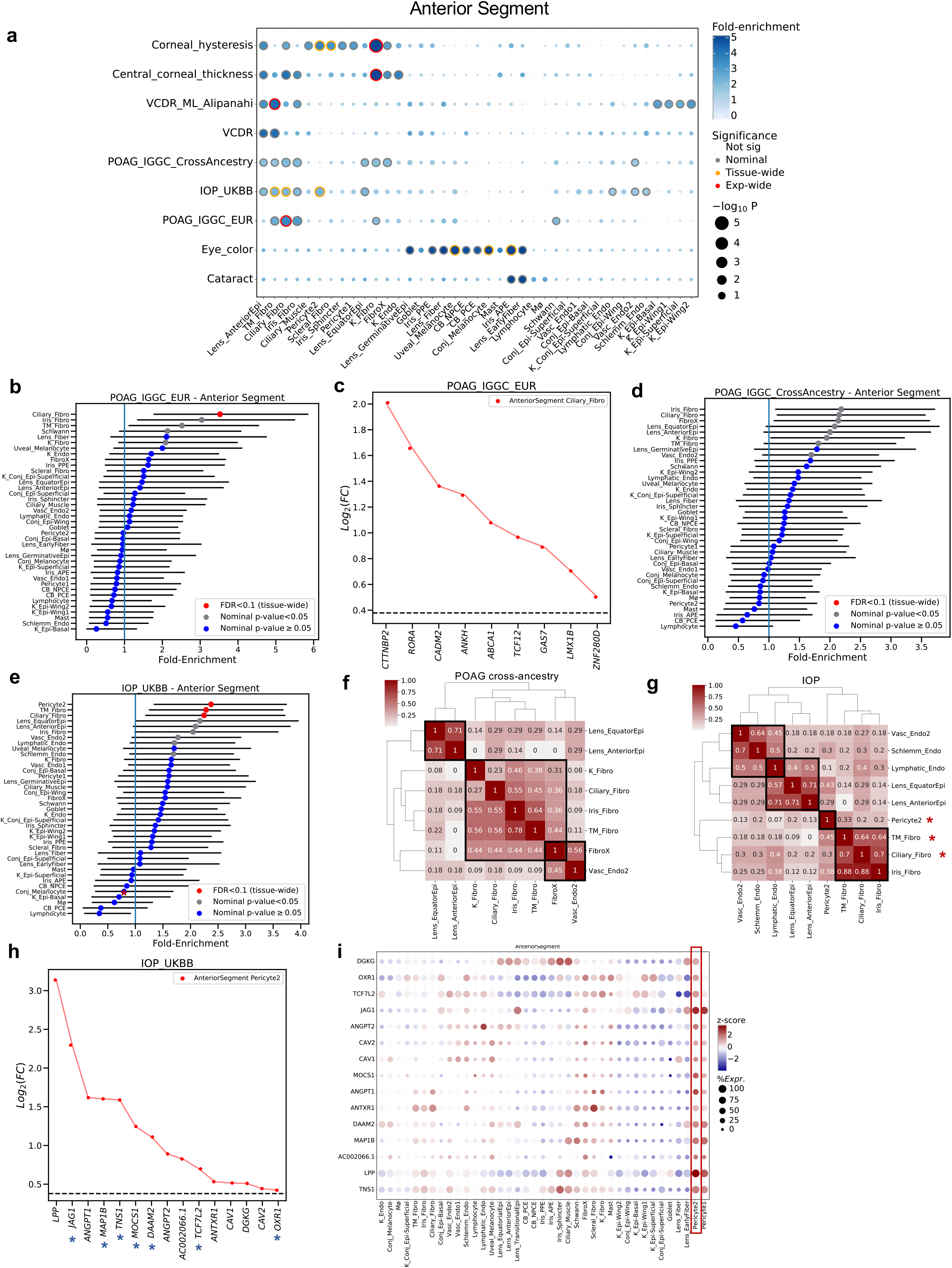
Cell type enrichment of e/sQTL-mapped genes for POAG, IOP and related trait loci in the anterior segment of the eye. **a**, Significance (circle size, -log10(P-value)) and fold-enrichment (circle color) of the cell type specificity of GWAS locus sets for POAG cross-ancestry, POAR European subset, IOP, central cornea thickness, corneal hysteresis, physician-defined vertical-cup-to-disc ratio (VCDR), and machine learning (ML)-defined VCDR (VCDR_ML_Alipanahi) are shown for each of the 39 cell types found in six tissues in the anterior segment of the eye^36^. Traits (rows) and cell types (columns) were clustered based on hierarchical clustering of the euclidean distance between GWAS locus set cell type-specificity enrichment scores. Red rings: experiment-wide significant (Benjamini Hochberg (BH) FDR<0.1); Yellow rings: tissue-wide significant (BH FDR<0.1); Grey rings: nominal significant (P<0.05). **b,d,e,** Cell type specificity fold-enrichment (x-axis) in the different anterior segment cell types ranked in descending order for the POAG European subset (**b**), POAG cross-ancestry (**d**), and IOP (**e**) GWAS locus sets. Error bars: 95% confidence intervals. Red: tissue-wide significant (FDR<0.1); Grey: nominal significant (P<0.05); Blue: non-significant (P≥0.05). **c,h,** Differential gene expression (log_2_(Fold-change), y axis) in the most strongly enriched cell type compared to all other cell types is shown for the set of genes (x axis) driving the enrichment signal of the POAG European GWAS loci in ciliary fibroblasts (**c**) and the IOP GWAS loci in pericytes (cluster 2) (**h**). Vertical dashed line represents log_2_(Fold-change) of 0.375 (FC=1.3) and FDR<0.1 that was used as the cell type-specificity enrichment cutoff. Asterisks denote genes in IOP loci not associated with POAG risk (**h**). **f-g**, Heatmap of fraction of genes that overlap between the e/sGenes driving the enrichment signal for top ranked cell types (P<0.05) in the anterior segment for POAG cross-ancestry (**f**) and IOP (**g**) GWAS loci. Numbers refer to the fraction of e/sGenes driving the cell type enrichment on each row that overlaps with the genes driving the cell type enrichment on the corresponding column. Hierarchical clustering was performed on both rows and columns using the euclidean distance between fractions. Red asterisks: tissue-wide BH FDR<0.1. **i,** Bubble map displaying the expression of the e/sGenes driving the IOP enrichment in pericytes across all cell types in the anterior segment. The colorbar represents gene expression z-scores computed by comparing each gene’s average expression in a given cell type to its per cell type average expression across all types divided by the standard deviation of all cell type expression averages. Bubble size is proportional to the percentage of cells expressing the given gene (log(TPK+1)>1). Cell type abbreviations are described in Supplementary Table 38.

### Enrichment of POAG and IOP colocalizing e/sGenes in biological processes

To gain biological insight into ways the implicated genes might contribute to glaucoma pathogenesis, we next tested whether the target genes of all the colocalizing e/sQTLs with POAG cross-ancestry, POAG EUR, or IOP GWAS loci were enriched in specific biological pathways, gene ontologies, or mouse phenotype ontologies, using *GeneEnrich* (Methods and Fig. 1c). Genes that colocalized with POAG cross-ancestry loci were significantly enriched in elastic fiber formation (Empirical P-value (P)<1×10^-5^, FDR<0.001) and extracellular matrix organization (P=3×10^-5^, FDR=0.012), and nominally enriched (P<0.05) in the transforming growth factor beta (TGF) receptor signaling pathway (P=3×10^-4^) and abnormal eye morphology (P=2.6×10^-3^), amongst others (Supplementary Table 30 and Supplementary Fig. 14a,b). Genes that colocalized with POAG EUR loci were nominally enriched (P < 4×10^-3^) in cellular senescence and cell cycle processes (e.g., Cyclin D-associated events in G1), lipid-related processes, such as apolipoprotein binding and decreased circulating high-density lipoprotein cholesterol level, and retina or neuronal related processes, including abnormal retina morphology, abnormal sensory neuron innervation pattern, and negative regulation of axon extension involved in axon guidance (Supplementary Table 31 and Supplementary Fig. 14c,d).

For the IOP genes, significant enrichment (P=2×10^-5^, FDR=0.025) was found in transcriptional regulation by *VENTX*, a gene that encodes a homeodomain-containing transcription factor (Supplementary Table 32 and Supplementary fig. 14e). *VENTX* and its IOP-colocalizing target genes driving the gene set enrichment signal (*ANAPC1*, *ANAPC7*, *AGO4*, *MOV10*, *TCF7L2*) were most highly expressed in immune cell types, lymphocytes and macrophages, in the single cell anterior segment and optic nerve head described below (Supplementary Fig. 14f). The IOP genes were also significantly enriched (FDR<0.15) in blood vessel morphogenesis and vasculature development, regulation of cytoskeletal organization, negative regulation of cellular component organization, and adherens junction (Supplementary Table 32 and Supplementary Fig. 14e). Since colocalization with multiple e/sQTLs was found for two GWAS loci in the HLA region on chromosome 6 associated with POAG and IOP (29 and 35 e/sGenes, respectively), likely due to high LD in the HLA region, we removed this region from the gene set enrichment analysis above to avoid inflating the results due to a single locus. When kept in, the endosomal vacuolar pathway, interferon gamma signaling, antigen presentation folding assembly and peptide loading of class I MHC, negative regulation of natural killer cell-mediated immunity, and cell aging were significantly enriched (FDR<0.1) for POAG genes (Supplementary Table 33), in addition to the gene sets above. The colocalizing POAG and IOP genes driving the gene set enrichment signals are listed in Supplementary Tables 30-33.

### Identifying pathogenic cell types for POAG and related ocular traits

To further relate the implicated genes to pathogenic mechanisms and cell types, we next tested whether the expression of POAG or IOP colocalizing e/sGenes was enriched in specific cell types in key eye tissues implicated in the pathophysiology of POAG. We first applied ECLIPSER^38, 39^ (Methods and Fig. 1d) to 228, 118, and 279 e/sGenes that colocalized with POAG cross-ancestry, POAG EUR and IOP GWAS loci, respectively (Supplementary Tables 13 and 34), and to cell type-specific expression from single nucleus (sn) RNA-seq of 13 tissues dissected from non-diseased human eyes: central cornea, corneoscleral wedge (CSW), trabecular meshwork (TM) including Schlemm’s canal, iris, ciliary body (CB), lens^36^ (all from anterior segment), peripheral and macular retina^33, 36^, the optic nerve head (ONH), optic nerve (ON), peripapillary sclera (PPS), peripheral sclera, and choroid^37^ (all from posterior segment) (Methods and Supplementary Tables 35-37). The cell type enrichment results are summarized in Supplementary Table 38.

In the anterior segment, we found significant enrichment (tissue-wide FDR<0.1) for POAG EUR loci in fibroblasts derived from the ciliary muscle (present in CB, CSW and TM^36^), annotated as ciliary fibroblasts in van Zyl *et al.*^36^, followed by fibroblasts derived from the iris root (present within the iris^36^), annotated as iris fibroblasts (Supplementary Table 38 and Fig. 6a,b). The ciliary muscle and iris are key tissues involved in the unconventional outflow pathway. These fibroblasts were also detected histologically within the TM where all three tissues meet and interweave at the iridocorneal angle^36^, implicating the conventional aqueous outflow pathway as well. Fig. 6c shows the e/sGenes driving the POAG EUR enrichment signal in ciliary fibroblasts. The POAG EUR genes were also modestly enriched in fibroblasts derived predominantly from the TM tissue (annotated as TM fibroblasts^36^) (P=0.014, FDR=0.18). For POAG cross-ancestry and IOP loci, we found supportive enrichment (P<0.05, FDR<0.23) in outflow pathway and cornea fibroblasts, vascular endothelium cells (cluster 2 derived from TM, CSW and CB tissues^36^), and lens epithelium, as detailed in Fig. 6a,d,e and Supplementary Table 38. Genes that colocalized with IOP loci were also significantly enriched (FDR<0.1) in pericytes (cluster 2 that localizes to the CSW^36^), and nominally enriched in lymphatic endothelium and Schlemm’s canal, whose dysfunction can lead to elevated IOP^47^ (Fig. 6e and Supplementary Table 38).

Clustering of the top-ranked anterior segment cell types (P<0.05) for POAG and IOP, separately, based on the overlap of genes driving the cell type enrichment, suggests three cell classes affecting POAG - fibroblasts, vascular endothelium, and lens epithelium; and three cell classes for IOP - fibroblasts, pericytes, and lymphatic endothelial cells (Fig. 6f, g). Between 45-78% of the genes driving the POAG enrichment signals in the outflow fibroblast cell types are common between the different fibroblasts (Fig. 6f), suggesting both shared and distinct genes acting in the conventional and unconventional outflow pathways. The IOP genes driving the enrichment signal in pericytes (Fig. 6h,i) were largely distinct from those enriched in vascular and fibroblast cell types (overlap 7-33%; Fig. 6g), and were enriched in vasculature development (P=3×10^-5^, FDR=0.075; Supplementary Table 40). On the other hand, the IOP genes driving the enrichment in TM fibroblasts (Supplementary Fig. 15c,d) were highly shared with the IOP genes enriched in ciliary and iris fibroblasts (overlap 64-88%; Fig. 6g). Notably, the enrichment of IOP genes in pericytes was specific to IOP (asterisks in Fig. 6h). When ECLIPSER was applied to genes that colocalized with IOP loci not associated with POAG, only the enrichment in pericytes remained (P=0.007) (Supplementary Table 41 and Supplementary Fig. 15e). Genes mapped to shared IOP and POAG loci were significantly enriched in ciliary and TM fibroblasts (FDR<0.01) and lymphatic or vascular endothelial cells (FDR=0.026). No enrichment was found for POAG-only loci in the anterior segment cell types, supporting IOP-dependent mechanisms in the anterior segment for POAG risk, as expected (Supplementary Table 41).

We next tested for enrichment of POAG and IOP colocalizing e/sGenes in retina snRNA-seq data (Methods). We found significant enrichment of POAG cross-ancestry genes in astrocytes and Müller glia cells (FDR<0.04; Supplementary Table 38, Fig. 7a and Supplementary Fig. 16a), which replicated (FDR<0.06) in a separate snRNA-seq study of the macula (Methods; Supplementary Table 38 and Supplementary Fig. 16g). Consistent results were found for POAG EUR genes (Supplementary Fig. 16b). A quarter (*YAP1, LPP, TRIB2)* of the 12 POAG cross-ancestry genes driving the astrocyte enrichment were common with Müller glia cells (Supplementary Fig. 16d, e), suggesting both shared and distinct processes between the two cell types. IOP genes were only nominally enriched in astrocytes (P=0.032; Supplementary Fig. 16c). By testing POAG- or IOP-only loci and shared loci, the POAG enrichment in retinal astrocytes and Müller glia cells appears to be independent of IOP (Supplementary Table 41 and Supplementary Fig. 17; more details in Supplementary Note). The POAG cross-ancestry genes were also enriched in RPE cells and S-cones in the macula (FDR=0.06). Of note, no significant enrichment was observed in RGCs (Fig. 7a), though some POAG colocalizing e/sGenes are expressed in RGCs (Supplementary Fig. 20).

**Figure 7.**
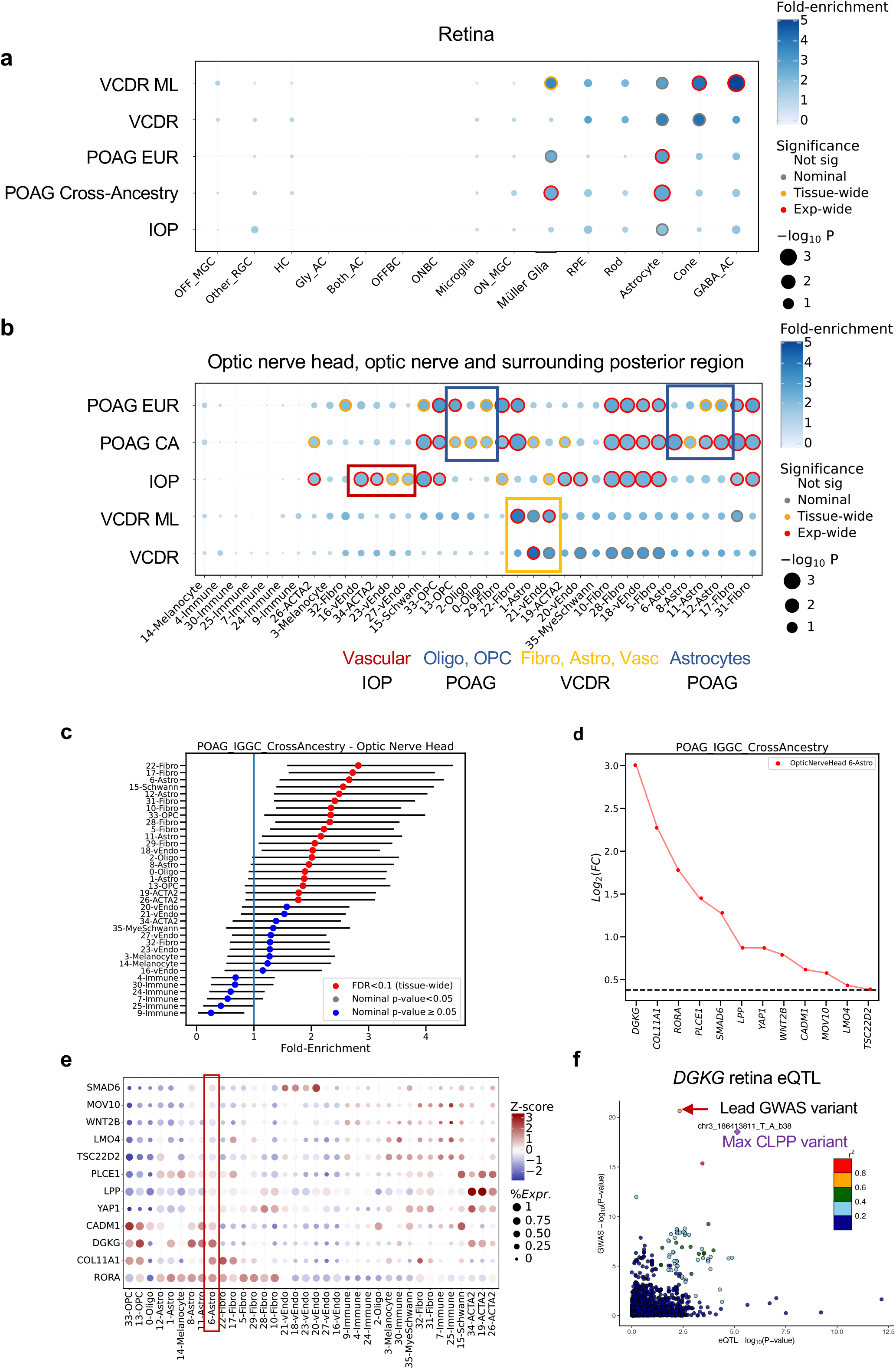
Cell type enrichment of e/sQTL-mapped genes for POAG, IOP and related trait loci in retina, optic nerve head and surrounding tissues. **a,b**, Significance (circle size, -log10(P-value)) and fold-enrichment (circle color) of the cell type specificity of GWAS locus sets for POAG cross-ancestry, POAR European subset, IOP, and physician (VCDR) and machine learning (ML)-defined VCDR (VCDR_ML_Alipanahi) are shown for each of the 15 cell types found in retina **(a)** and 36 cell types in optic nerve head (ONH), optic nerve (ON), peripapillary sclera, sclera and choroid **(b)**. Traits (rows) and cell types (columns) were clustered based on hierarchical clustering of the euclidean distance between GWAS locus set cell type-specificity enrichment scores. Red rings: experiment-wide significant (BH FDR<0.1); Yellow rings: tissue-wide significant (BH FDR<0.1); Grey rings: nominal significant (P<0.05). **c,** Cell type specificity fold-enrichment (x-axis) in the ONH and surrounding tissue cell types ranked in descending order for the POAG cross-ancestry GWAS locus set. **d,** Differential expression (log_2_(Fold-change)) in astrocytes, the most strongly enriched cell type for POAG cross-ancestry GWAS loci in the ONH, compared to all other cell types in the posterior tissues is shown for the set of genes driving the POAG enrichment signal in astrocytes. Horizontal dashed line represents log_2_(Fold-change) of 0.375 (FC=1.3) and FDR<0.1 that was used as the cell type-specificity enrichment cutoff. **e,** The expression profile of the e/sGenes driving the POAG cross-ancestry enrichment signal in astrocytes is shown across all cell types in the ONH and surrounding tissues. Color represents z-scores computed by comparing each gene’s average expression in a given cell type to the average expression across all cell types divided by its standard deviation of all cell type expression averages. Bubble size is proportional to the percentage of cells expressing the gene (log(TPK+1)>1). **f,** LocusCompare plot of -log_10_ (P-value) of the POAG cross-ancestry GWAS meta-analysis relative to -log_10_(P-value) of a retina eQTL acting on *DGKG* that significantly colocalized with the POAG GWAS locus rs56233426 (chr3_186411027_G_A). *DGKG* is the strongest astrocyte-specific gene driving the cell type enrichment signal for POAG loci. Points are color-coded based on LD (r^2^) relative to the eVariant with the highest colocalization posterior probability (CLPP=0.93). The red arrow is pointing to the lead POAG GWAS variant. Cell type abbreviations are described in Supplementary Table 38.

Finally, we tested for cell type-specific enrichment in the optic nerve head (ONH), optic nerve (ON) and adjacent posterior tissues (Methods; Supplementary Table 37). The strongest enrichment (FDR<0.01) of POAG cross-ancestry genes was found in fibroblasts primarily in the peripapillary sclera (PPS) that encompasses the ONH, followed by fibroblasts most abundant in the choroid, astrocytes that reside in the ONH and ON, Schwann cells in the choroid and PPS, oligodendrocyte precursor cells (OPCs) and oligodendrocytes in the ON and ONH (Supplementary Fig. 18f,g), and vascular endothelium cells primarily in the choroid (FDR<0.09; Fig 7b,c, Supplementary Table 38 and Supplementary Fig. 18a,d). POAG EUR genes showed similar enrichment patterns (Fig. 7b, Supplementary Fig. 18b and Supplementary Table 38). About half the genes driving the enrichment in astrocytes in ONH (Fig. 7d, e) and retina samples from separate donors were common (e.g., *DGKG, PLCE1, LPP*, *GAS7*, *YAP1*, and *COL11A1*; Supplementary Table 38). *DGKG*, diacylglycerol kinase gamma, whose retina-specific eQTL colocalized (CLPP=0.96) with POAG cross-ancestry association (Fig. 7f) displayed the strongest cell type specificity in ONH (Fig. 7d) and retinal astrocytes (Supplementary Fig. 16d).

IOP genes were most significantly enriched in vascular endothelial cells and fibroblasts primarily residing in the choroid, but also in the ONH and PPS (FDR<0.014), followed by Schwann cells in the choroid and PPS, vascular smooth muscle cells (19-ACTA2; Supplementary Fig. 18h,i) in the PPS and sclera, pericytes (26-ACTA2) in the ONH, PPS and choroid, and OPCs in the ON (FDR<0.06; Fig. 7b, Supplementary Table 38, and Supplementary Fig. 18a,c,e). The IOP genes driving enrichment in vascular endothelial cells in the ONH, choroid and posterior tissues were enriched in vasculature development and anchoring junction gene ontologies (FDR<0.06), and IOP genes enriched in pericytes in the ONH, choroid and PPS were enriched in TIE2 signaling (FDR=0.02), response to carbohydrate adhesion (FDR=0.13), and negative regulation of cell adhesion (FDR=0.13) (Supplementary Table 40). Notably, the enrichment in oligodendrocytes and OPCs was specific to POAG-only loci, and enrichment in vascular endothelium and mural cells was specific to IOP-only loci (Supplementary Table 41 and Supplementary Fig. 19).

The cell type expression profiles of all POAG cross-ancestry, POAG EUR and IOP colocalizing e/sGenes is shown in Supplementary Fig. 20, and a summary of the cell types and pathways in which each of the POAG and IOP colocalizing e/sGenes are enriched is presented in Supplementary Tables 42-44. Applying ECLIPSER to various negative control traits suggests that the cell type enrichment results are specific to glaucoma and not due to unaccounted confounding factors (Supplementary Tables 45-46, Fig. 6a, and Supplementary Fig. 21; Supplementary Note). Furthermore, the ECLIPSER cell type enrichment significance did not correlate with cell count per cell type in the single-nucleus datasets (Pearson’s R^2^<0.2, P>0.12; Supplementary Table 47).

To increase confidence in the POAG and IOP cell type enrichment results, we applied two additional methods that identify cell types associated with complex traits, through regression analysis of genome-wide associations beyond known GWAS loci: stratified LD score regression (S-LDSC) and MAGMA (Fig. 1e; Methods). The primary enriched cell types for POAG and IOP found with ECLIPSER, including ciliary and TM fibroblasts, ONH fibroblasts, and retinal macroglial cells, were significant with S-LDSC (Supplementary Table 48; Supplementary Fig. 22) and more restrictively with MAGMA (Supplementary Table 49; Supplementary Fig. 22); additional enrichment was found in vascular types. Enrichment of POAG loci in ONH and ON oligodendrocytes was only found with ECLIPSER suggesting that the enrichment is primarily driven by genes with strong genetic effects. The cell type enrichment significance of ECLIPSER was reasonably correlated with that of S-LDSC and MAGMA (Average Pearson’s r=0.53, range: 0.18-0.86; Supplementary Table 50). We further used conditional analysis implemented in MAGMA to test whether the different cell type enrichment signals for POAG or IOP were independent of each other in each tissue (Supplementary Table 51). In the anterior segment, the enrichment of POAG associations in ciliary fibroblasts was independent of TM fibroblasts, but not vice versa (Conditional P=0.04). In retina, POAG associations were significantly enriched in astrocytes (P<6E-8) and Müller glia cells (P<0.002), but only astrocytes remained significant after conditional analysis (Conditional P<4E-6), suggesting that astrocytes may play a more important role in glaucoma pathogenicity than Müller Glia cells. In the ONH, the POAG and IOP enrichment in fibroblasts, astrocytes, vascular endothelium, and mural cells were all independent of each other (Supplementary Table 51).

Finally, to augment the POAG and IOP enrichment analysis, we tested for cell type enrichment of genes mapped to GWAS loci of additional glaucoma associated traits (listed in Supplementary Table 34), including vertical-cup-to-disc ratio (VCDR), cornea hysteresis, and central cornea thickness (Methods) in all ocular tissue regions (Supplementary Table 38, Fig. 6a and Fig. 7a, b). In the anterior segment, genes mapped to central corneal thickness and corneal hysteresis were most significantly enriched in corneal fibroblasts (FDR<0.007; Fig. 6a and Supplementary Fig. 23a, b), highlighting the specificity of the POAG and IOP gene enrichment in the outflow pathway fibroblasts. The VCDR GWAS loci from a well-powered GWAS that used deep learning (ML) to score the characteristics of fundus images from 65,680 European individuals^12^ showed significant enrichment in the TM fibroblasts from the conventional outflow pathway (FDR=0.02; Fig. 6a and Supplementary Fig. 23c). TM fibroblasts were also the top nominally enriched cell type for a smaller VCDR GWAS, where 23,899 fundus images were manually scored by ophthalmologists^13^ (P=0.0076, FDR=0.3; Fig. 6a and Supplementary Fig. 23d). In the retina, the ML-based VCDR loci displayed significant enrichment in GABAergic amacrine cells, cone photoreceptors and Müller glia cells (FDR<0.085; Fig. 7a and Supplementary Fig. 23e) and nominal enrichment in astrocytes. In the ONH, the ML-based VCDR genes were nominally enriched in fibroblasts in the PPS, vascular endothelium primarily in the choroid, and astrocytes in the ONH and ON, similarly to POAG loci (Fig. 7b and Supplementary Fig. 23g, h; Supplementary Table 38).

In summary, our cell type enrichment analysis has revealed roles for both known and less well-studied cell types in POAG pathogenicity, such as fibroblasts in the unconventional and conventional outflow pathways, astrocytes in retina and ONH, OPCs in the ON and ONH, and Schwann cells and fibroblasts in the PPS and choroid. It also suggests known and new causal genes for POAG and related eye traits that may be affecting glaucoma susceptibility through specific cell types in the anterior and posterior parts of the eye in IOP-dependent and independent manners (Supplementary Tables 38 and 41).

## Discussion

We report results of a systematic investigation of the underlying causal mechanisms, genes and cell types of over 130 cross-ancestry or European loci associated with POAG^8,9^ and over 110 loci associated with its major risk factor, elevated IOP^9^. Our analysis integrated a variety of datasets, including expression and splicing QTLs from 49 GTEx tissues^26^ and from retina^26, 27^, genome topology data from retina^40^, single-nucleus expression data from a whole-eye cell atlas that includes key structures of both the anterior^36^ and posterior segments^33, 37^, and the largest to date GWAS meta-analyses for these traits^8,9^. Our finding that eQTLs and sQTLs in GTEx tissues and retina are enriched for hundreds of known and more modest POAG and IOP associations, suggests a primary role for transcriptional regulation in POAG susceptibility, as observed for other diseases^24–27^, and implies that GTEx tissues can be used to uncover causal mechanisms for glaucoma. The GTEx e/sQTLs likely capture shared genetic regulation with the actual pathogenic tissues for glaucoma, such as fibroblasts and vascular endothelial cells, as well as shared regulation across cell types and tissues^26^.

Using two QTL/GWAS colocalization methods^41, 42^, we prioritized putative causal genes for ∼60% of the POAG and IOP GWAS loci. A similar fraction of GWAS loci with significant colocalization results has been found for other complex diseases and traits^26, 48^. For a quarter (80) of the POAG and IOP loci, a single gene was proposed, ten of which are noncoding genes (lincRNA and antisense), suggesting that transcriptional and post-transcriptional gene regulation contribute to glaucoma susceptibility. We provided additional support for three quarters of the colocalizing e/sGenes using Mendelian randomization^49–51^, which tests for horizontal pleiotropy, not accounted for by Bayesian colocalization analysis^52, 53^. For about one third of the GWAS loci, none of the proposed causal gene/s were the nearest gene to the lead GWAS variant, similar to that observed for other complex traits^24, 48, 54^. These results emphasize the value of using e/sQTLs or other functional assays that link regulatory regions to distal target genes^24, 55–57^ to prioritize causal genes underlying common variant associations.

Integrating e/sQTLs with POAG and IOP GWAS loci proposed both previously suggested^8^ and new biological processes for these traits. The POAG colocalizing genes, which included several known mendelian, early-onset glaucoma genes (*EFEMP1*^45, 58^ and *LTBP2*^59^), were most strongly enriched in extracellular matrix organization and elastic fiber formation, as previously reported^8^, followed by TGF receptor signaling pathway. Structural changes of the extracellular matrix induced by TGF-beta2 in both the trabecular meshwork in the outflow pathway and the optic nerve head have been associated with POAG^60^, and have been suggested to cause impairment of optic nerve axonal transport and neurotrophic supply that could influence RGC degeneration^60^. Regulation by the homeodomain transcription factor, VENTX, was the most significantly enriched gene set for IOP genes, which has not yet been associated with glaucoma. VENTX is proposed to play important roles during embryonic patterning (by homology), including in neural crest development^61^, as well as hematopoiesis, leukemogenesis, cellular senescence and macrophage differentiation^62^. Its strongest expression in our single cell data was in lymphocytes in the anterior segment, and macrophages in the optic nerve head, proposing a novel link between immune-related processes and IOP levels. Reduced circulating endothelial progenitor cells has been reported in POAG patients^63^, which could explain impaired flow-mediated vasodilation in POAG^64^. Additional processes suggested to affect IOP regulation, aside from previously suggested^9^ vascular development, are regulation of cytoskeleton organization, and adherens junction, a cell-cell junction whose cytoplasmic face is linked to the actin cytoskeleton, and that allows cells to respond to biomechanical forces and structural changes in the tissue microenvironment^65^. Inhibition of adherens junction regulation in trabecular meshwork has been shown to modestly influence IOP levels in rabbits^66^. We also found modest enrichment of POAG genes in neuronal-related processes, including genes affecting retinal morphology, sensory neuron innervation pattern, and regulation of axon guidance. These genes may represent IOP-independent mechanisms, which will need to be corroborated in future GWAS with larger numbers of normal tension glaucoma cases.

In addition to prioritizing causal genes for POAG and IOP, e/sQTLs suggest the direction of effect of gene expression changes or alternative splicing on disease risk that could inform drug design. In this study, we provide various hypotheses of putative causal genes and regulatory mechanisms that may affect POAG susceptibility in an IOP-dependent or independent manner, for experimental follow up. For example, an increase in expression or alternative splicing of *TMCO1*, a gene that regulates the balance of calcium ions inside the endoplasmic reticulum, or a decrease in expression of *LMX1B*, LIM homeobox transcription factor 1 beta, that is essential for several developmental processes including the anterior segment of the eye^67^, were proposed to reduce POAG risk and IOP levels. The lead IOP GWAS variant (rs116089225) in *TMCO1* that colocalized with *TMCO1* e/sQTLs has been recently associated with variable number tandem repeat (VNTR) length in the UK biobank study^68^. However, the VNTR did not display allelic series association with *TMCO1* expression levels in GTEx^68^, suggesting that the GWAS variant might be tagging more than one causal mechanism. As for IOP-independent mechanisms, an sQTL acting on *CDKN2B-AS1*, which leads to skipping of exons 2 and 3 that overlap the *CDKN2B* gene on the opposite strand, is proposed as a potential mechanism of action for the protective signal found in this gene^69^.

Skipping of these exons might render the *CDKN2B* antisense less efficient in forming a complex with the *CDKN2B* RNA. Retinal Hi-C and epigenetic data further support potential roles for e/sQTL effects on POAG in the retina, such as increased expression of *RERE* (arginine-glutamic acid dipeptide repeats) proposed to increase POAG risk. Of note, *RERE* has also been associated with VCDR^13^. Overexpression of the *RERE* protein that co-localizes with a nuclear transcription factor triggers apoptosis, and its deficiency in mice causes retinal and optic nerve atrophy^70^. This study also suggested a potential secondary causal gene, *PIGC* for the strongest POAG association, a nonsense mutation in *MYOC*, which will need to be replicated in a larger independent POAG GWAS. The *MYOC* mutation causes aggregation of misfolded myocilin proteins in the trabecular meshwork, which may lead to elevated IOP levels^71^. Conversely, the enrichment of *PIGC* (that encodes an endoplasmic reticulum-associated protein) expression, along with other POAG-colocalizing genes, in oligodendrocytes in the optic nerve head (ONH), suggests a secondary causal role for this locus in RGC support, in the posterior part of the eye.

There are several reasons why we may not have found colocalizing e/sQTLs for 40% of the loci. First, some of the causal genes or regulatory effects may be specific to regions in the eye or rare cell types for which we do not yet have representative e/sQTLs. Second, some genes may affect POAG or IOP by perturbing processes only active during development or under specific conditions or stimuli, not captured in adult tissues. Third, the causal variant may be another type of molecular QTL not tested in this study, such as e/sQTL acting in *trans* or protein QTLs. Fourth, some of the genetic associations may be tagging deleterious protein-coding variants^8^. Finally, there are several limitations to Bayesian colocalization methodologies^52, 53^, as described in the Supplementary Note.

By applying a novel method (ECLIPSER^38, 39^) to the colocalizing e/sGenes and single nucleus expression data from glaucoma-relevant eye tissues, we provided support for previously implicated cell types affecting POAG development, and shed light on less well-established or novel pathogenic cell types for POAG and IOP. One of the unique features of ECLIPSER, compared to other single cell enrichment methods^72–76^ is that it identifies cell types that are specific to a given disease or trait, compared to a range of unrelated complex diseases and traits. Our results found that gene expression variation in the ciliary and iris fibroblasts in the unconventional outflow pathway in the anterior segment, in addition to the TM cells and Schlemm’s canal cells in the conventional outflow pathway, may both be key contributors to local IOP regulation and POAG risk. The expression profile of the ciliary fibroblasts that was most strongly enriched for POAG genes, is most similar to the ‘beam cell A’ defined in the single cell RNA-seq atlas of the outflow pathway in van Zyl *et al*.^34^ (Fig. S6B in ^34^), which populates the ciliary muscle and uveal base of the TM; the iris fibroblasts are most similar to ‘beam cell B’, and the TM fibroblasts to the JCT (juxtacanalicular trabecular meshwork) cell type that resides adjacent to the Schlemm’s canal. Since the role of the unconventional outflow pathway in IOP homeostasis remains relatively understudied compared to that of the conventional pathway^77^, these findings may encourage further avenues of investigation. Furthermore, the enrichment of IOP genes in pericytes, many of which are not currently associated with POAG, extends our understanding of how genetic variation may be affecting IOP in the anterior segment. Pericytes are mural cells that wrap around the endothelial cells that line capillary blood vessels. A recent study has found reduced capillary diameter and impaired blood flow at pericyte locations in mouse eyes with high IOP^78^.

In the peripheral and macular retina, we found significant enrichment of POAG colocalizing genes in astrocyte and Müller glia cells. Astrocyte and Müller glia are two types of macroglia cells that interact with RGCs and blood vessels, and play an important role in retinal homeostasis, including metabolic supply and structural support, maintaining the extracellular environment of the neurons, and neurotransmitter transmission. Müller glia, the most common glial cell in the retina, span the entire retinal layer, while astrocytes are present only in the innermost layer of the retina. Several studies in animal models and patients with glaucoma^79–81^ have found that astrocytes and Müller glia cells become reactive at early stages of glaucomatous conditions when RGCs are intact, suggesting a role for macroglia in the initiation and progression of glaucoma. POAG genes, but not IOP genes, were also enriched in astrocyte types residing primarily in the ONH that contains the lamina cribrosa (LC)^82^, a mesh-like structure where unmyelinated RGCs pass through the sclera to exit the eye, and another in the ONH and optic nerve (ON). Notably, astrocytes have been found to be one of the major cell types isolated from human ONH^82^ and in LC dissected from human ONH^83^, and make up ∼20% of the cells in our ONH snRNA-seq dataset.

In the ONH and surrounding posterior tissues, POAG genes were most strongly enriched in fibroblasts abundant in the peripapillary sclera (PPS), which surrounds the ONH. Pressure on the ONH and PPS that is a continuum of the LC can cause astrocyte reactivity and compression of RGC axons that can lead to RGC death^84, 85^. Genes mapped to POAG, but not IOP loci were also enriched in oligodendrocytes that form a myelin sheath around the axons of RGCs, and oligodendrocyte precursor cells found in the ON, suggesting new IOP-independent mechanisms that can affect optic nerve degeneration. The strongest enrichment for IOP was in vascular endothelial (also enriched for POAG genes) and fibroblast cells primarily residing in the choroid, but also in the ONH and PPS, suggesting that vascular structural abnormalities or functional dysregulation of blood flow to the optic nerve and retina may be an important contributor to POAG^86^. It is also possible that the enrichment in the ONH vascular endothelial cells is capturing causal mechanisms acting in the vascular endothelial cells in the anterior segment that were enriched for most of the same IOP genes as in ONH. The IOP genes were also enriched in vascular smooth muscle cells (VSMC) in the PPS and sclera. These VSMC-specific genes were enriched in lipid binding and negative regulation of cell substrate adhesion processes, suggesting a role in cytoskeleton-associated cell-cell adhesion and cell-extracellular matrix adhesion. These muscle cells may be part of the LC, as LC cells isolated from human ONH were found to stain for alpha-smooth muscle actin^82, 83^. Cells in the LC produce extracellular matrix proteins to support the LC structure^83^, and biomechanical strain on the LC, such as from elevated IOP, is thought to be one of the causes of RGC degeneration^87, 88^. Further investigation will be needed to determine whether the muscle cells found in the ONH single nucleus dataset^37^ reside in the LC or in other structures such as blood vessels.

Notably, we did not find significant enrichment of cell type-specific expression of POAG or IOP genes in RGCs, whose cell death is the key characteristic of glaucoma, but rather in neuronal support cells. This highlights the importance of targeting the support cells in new therapy design. It should be noted that multiple colocalizing POAG genes are expressed in RGCs (see Supplementary Fig. 20 and Monavarfeshani *et al*.^37^), however they are also expressed in other retinal cell types. Hence, the potential effect of these genes on POAG via RGCs merits further investigation. Furthermore, while several studies have suggested that microglia, specialized macrophage-like cells, may affect RGC survival^89^, we did not find support for a causal role of microglia or immune cells in POAG susceptibility. A less expected result was the enrichment of POAG genes in RPE cells, which was also shown in the posterior ocular cell atlas^37^. In all, our findings in retina, ONH and the surrounding tissues propose cell types and biological processes that may be viable targets for neuroprotective therapies.

A potential limitation of our GWAS-cell type enrichment method, ECLIPSER is that it only considers genes that map to genome-wide significant loci and not subthreshold associations. We thus provided further support for our cell type enrichment results using two additional methods, stratified LD score regression and MAGMA, that analyze multiple modest associations genome-wide in additional to known GWAS loci. Furthermore, ECLIPSER primarily considers genes whose expression is specific to one or few cell types within a tissue, as the cell type specificity scoring metric was found to be successful in identifying known pathogenic cell types for a range of complex diseases and traits in a cross-tissue single nucleus expression atlas using GTEx samples^38^. We note though that genes expressed at similar levels across most or all cell types may also contribute to disease risk or trait variation and would be missed with this approach.

In conclusion, our work has generated new insights into POAG mechanisms, which could inform the development of novel therapies targeting IOP reduction and neuroprotection. By integrating genetic regulation and single cell expression in glaucoma-relevant ocular tissues with GWAS summary statistics we have identified known and new causal genes and biological processes; proposed key ocular cell types that may be pathogenic for glaucoma; and provided evidence for the existence of hundreds of novel genetic associations of regulatory effects for glaucoma. In the future, detection of e/sQTLs in relevant eye tissues and at the cellular level^90, 91^ is expected to provide a more complete picture of the causal molecular and cellular mechanisms of POAG risk and IOP variation.

## Methodss

### GWAS datasets

We applied colocalization and fine-mapping analysis to 127 GWAS loci identified in the cross-ancestry POAG GWAS meta-analysis of 34,179 cases and 349,321 controls from European, African, and East Asian populations^8^, 68 GWAS loci from the GWAS meta-analysis of the European subset of 16,677 POAG cases and 199,580 controls^8^, and 133 LD-independent GWAS variants in 112 loci from the IOP GWAS meta-analysis of 139,555 primarily UK Biobank (European) samples^9^. The GWAS meta-analysis summary statistics, which included p-value, effect size and standard error, were obtained from the corresponding studies. Chromosome positions were lifted over from genome build 37 (hg19) to hg38. Association results on chromosome X were only available for the POAG GWAS meta-analyses (cross-ancestry and European subset).

### GTEx and EyeGEx QTL datasets

*cis*-eQTLs and *cis*-sQTLs from 49 tissues from GTEx release v8^26^ and *cis*-eQTLs from peripheral retina^27^ were used in this study. Summary statistics of all variant-gene e/sQTL pairs tested in each of the 50 tissues, the significant e/sGenes and e/sVariants at FDR<0.05, and the gene expression levels and LeafCutter^92^ values are available for download from the GTEx portal (URLs). The summary statistics of all variant-gene pairs tested per gene and tissue was used as input to the colocalization analysis, and the LocusZoom^93^ and LocusCompare plots (URLs). Plots of exon and exon junction read counts were taken from the visualizations on the GTEx portal (URLs). GENCODE versions 26 and 25 were used for the GTEx v8 and EyeGEx studies, respectively.

### Enrichment of POAG and IOP associations among e/sQTLs using *QTLEnrich*

To test whether genome-wide significant and nominal POAG and IOP trait associations are enriched among eQTLs and sQTLs, and to assess the contribution of e/sQTLs to these traits, we applied *QTLEnrich*^24, 26^ to the POAG and IOP GWAS meta-analyses summary statistics, using eQTLs and sQTLs from the 49 GTEx tissues^26, 27^ and eQTLs from peripheral retina (EyeGEx^27^). *QTLEnrich* is a rank and permutation-based method that evaluates the fold-enrichment significance of trait associations among a set of e/sQTLs in a given tissue, correcting for three confounding factors: minor allele frequency (MAF), distance to the target gene’s transcription start site, and local LD^24^ (for more details see Supplementary Note). Only protein-coding and lincRNA genes were considered in this analysis. Significant tissues were determined based on an Enrichment P-value that passed Bonferroni correction, correcting for 50 tissues and two QTL types tested (P<5×10^-4^). The adjusted fold-enrichment was used to rank the significantly enriched tissues, as this statistic is not correlated with tissue sample size or number of significant e/sQTLs per tissue^24^, as observed with the colocalization analysis (Supplementary Fig. 4). For the significant trait-tissue pairs, the fraction and number of e/sVariants proposed to be associated with POAG or IOP were estimated using an empirically derived, true positive rate (Adj. !_!_) approach that we implemented in the latest version of *QTLEnrich* (URLs), based on Storey’s analytical !_!_^94, 95^ and an empirical FDR method^94, 95^ (see Supplementary Note).

### Colocalization analysis

To identify a high confidence set of genes and regulatory mechanisms (e/sQTLs) that may be mediating the functional mechanisms underlying known common variant associations with POAG and IOP, we applied two Bayesian-based colocalization methods: eCAVIAR^41^ and *enloc*^42^. These methods assess the probability that co-occurring GWAS and e/sQTL signals are tagging the same causal variant or haplotype, accounting for local LD and allelic heterogeneity, using slightly different fine-mapping and colocalization approaches. They are applied to GWAS and QTL summary-level statistics enabling the analysis of large, well-powered GWAS meta-analyses, for which genotype data are not available. For the *enloc* analysis, DAP-G^42^ was used to perform fine-mapping of GWAS and e/sQTL loci to estimate the posterior probabilities of each variant in each locus being the causal variant, while eCAVIAR has the fine-mapping feature built in. We applied the two colocalization methods to 127 POAG cross-ancestry GWAS, 68 POAG GWAS loci from the European subset meta-analysis, and 133 independent IOP variants (112 loci) from a primarily European study (described above). Z-scores from the GWAS and GTEx e/sQTL studies, computed as the effect size (beta) divided by the standard error of the effect size for each variant, were used as input into eCAVIAR and DAP-G. For the retina eQTLs, we computed z-scores from the variant association p-values assuming a chi-square distribution with 1 degree of freedom.

All GWAS loci were tested for colocalization with all eQTLs and sQTLs from 49 GTEx tissues^26^ and peripheral retina eQTLs^27^ that had at least 5 e/sVariants (FDR<0.05) within the GWAS locus LD interval. An LD window around each lead GWAS variant was defined as the chromosome positions on either side containing variants within r^2^ > 0.1, determined using 1000 Genomes Project Phase 3^96^ as the reference panel, and extending an additional 50kb on either side. For the IOP and POAG EUR loci, LD was computed using only the European samples in 1000 Genomes Project, while for the cross-ancestry POAG loci, LD was computed using the European, African, and East Asian samples in 1000 Genomes. If a GWAS variant was not found in 1000 Genomes, an LD proxy variant (r^2^ > 0.8) was searched for in GTEx, and if not found, the nearest variant was used. The interval boundaries and number of variants tested are reported in Supplementary Tables 7-13. eCAVIAR and *enloc* analyses were applied to all common variants (MAF>1%) that fell within the GWAS LD intervals and were present in both the GWAS and e/sQTL studies. The effect allele of the variants in each GWAS was aligned relative to the alternative (ALT) allele that was used as the effect allele in GTEx and EyeGEx. Colocalization analysis of the retina eQTLs was only performed using eCAVIAR. GWAS-e/sQTL-tissue combinations with a colocalization posterior probability (CLPP) above 0.01 were considered significant with eCAVIAR and/or with an RCP above 0.1 were considered significant with *enloc* based on the methods’ recommendations^41, 42, 52^. To remove potential false positives, we filtered out variant, gene, tissue, trait combinations where the e/sVariant with a significant colocalization result had a GWAS p-value above 1×10^-5^ or whose e/sQTL p-values was above 1×10^-4^ and/or did not pass FDR<0.05 (FALSE in column ‘Pass_QC_QTL_FDR05_P1E04_GWAS_P1E05’ in Supplementary Tables 7-12). Further details on the eCAVIAR and *enloc* analyses and quality control can be found in Supplementary Note.

### Mendelian randomization (MR)

Mendelian randomization (MR)^51^ was used to provide additional genetic support for a causal relationship between colocalizing e/sQTLs and POAG and/or IOP loci. Significant e/sVariants were used as the instrumental variable (IV) in MR to facilitate causal inference^97^ (See Supplementary Note). Two sample MR was applied to the summary statistics of the e/sQTLs (exposure) and POAG or IOP GWAS (outcome) for all significant colocalizing loci (Supplementary Tables 7-12), using the *TwoSampleMR* and *MendelianRandomization* packages in R (version 4.1.2)^98^. To avoid confounding by ancestry, MR was conducted using the European ancestry subset of the POAG GWAS and the IOP GWAS, which primarily contains European individuals. MR estimates were generated by calculating the Wald ratio, i.e., the variant-outcome association beta divided by the variant-exposure association beta^99^. Where multiple variants constituted the instrument for the candidate gene, the inverse-variance weighted (IVW) method was used as the primary method for pooling variant-specific estimates^100^. Given that the IVW approach assumes no horizontal pleiotropy, methods robust to violation of the exclusion-restriction assumption were used as sensitivity analyses. The simple-median^101^, weighted-median^101^, MR-Egger^102^, and MR-PRESSO^103^ methods were applied. Horizontal pleiotropy was tested using the Egger-intercept test and the MR-PRESSO global heterogeneity test, for which *P* < 0.05 indicated the presence of horizontal pleiotropy. MR associations with Benjamini-Hochberg (BH) FDR < 0.05 for the primary IVW/Wald ratio test were considered statistically significant. In cases where horizontal pleiotropy was found based only on the MR-PRESSO global heterogeneity test, an MR PRESSO outlier-corrected p-value < 0.05 was considered a significant result.

### Integration of retina Hi-C and epigenetic data with colocalizing POAG loci and e/sQTLs

To identify retina eQTLs or GTEx e/sQTLs that colocalized with POAG GWAS loci that may be exerting their causal effect on POAG in the retina, we inspected all POAG loci in the context of chromatin loops, *cis* regulatory elements (CREs) and super-enhancers (SEs) that were previously detected in retina from 5 postmortem non-diseased human donor eyes^40^. The loops were calculated from Hi-C (3D chromosome conformation capture) data, and the CREs and SEs from epigenetic data, as described in Marchal *et al*.^40^. The lead POAG GWAS variants and their LD proxy variants (r^2^>0.8), the colocalizing e/sQTLs and LD proxy variants, which are also significant e/sVariants (FDR<0.05), and the e/sQTL target genes were inspected for overlap or closest overlapping gene with the Hi-C loops, CREs, and SEs, using the closestBed command from bedtools (v2.27.1)^104^. For retina eQTLs and GTEx e/sQTLs GENCODE versions 25 and 26 were used, respectively, to overlap genes and TSS hg38 coordinates. Colocalizing e/sQTLs were proposed as putative causal genes to POAG, if the e/sVariant overlapped one foot of the loop and the second foot overlapped the gene body or TSS of the target gene. CRE and SE target genes were defined if the e/sVariant and gene body or TSS of the gene overlapped the same CRE or SE. The closest target genes identified using chromatin loops for the POAG cross-ancestry GWAS loci was taken from our recently published Hi-C study (Supplemental Data 4 in Marchal *et al*.^40^).

### Single nucleus RNA-seq datasets and differential gene expression

We analyzed gene expression values (log(TPK+1)) from four single-nucleus (sn) RNA-seq data sets from the following glaucoma-relevant regions of the eye: anterior segment^36^, retina^33, 36^, macula^36^, and optic nerve head and surrounding posterior tissues^37^. All tissue samples were dissected from non-diseased eye globes from post-mortem donors with no record of eye disease, and were de-identified. The number of cells per cell types in each of the tissues can be found in Supplementary Table 38. Differential gene expression was applied to genes expressed in at least 5% of cells in any cell type cluster in each of the datasets. Here is a brief description of the four datasets:

### Anterior segment

Six tissues in the anterior segment, including central cornea, corneoscleral wedge (CSW), trabecular meshwork (TM), iris, ciliary body (CB) and lens, were dissected from six donors within 6 hours from death, as described in^36^. To be able to compare across cell types between tissues in the anterior segment, the snRNA-seq data from cornea, CSW, CB, iris, and TM were pooled, downsampled to 1000 cells per type in each tissue, and reclustered yielding 34 clusters^36^. Five clusters were identified for the lens. Differential gene expression analysis between each cell type and all other cell types was performed using the regression model in MAST^105^ that corrects for the proportion of genes expressed per cell.

### Retina

Retina samples from the fovea (4mm punch), macula (6 mm punch) and/or periphery were collected from six donors within 6 hours from death from the Utah Lions Eye Bank, flash frozen and processed as described in^33^ (more details in Supplementary Note). The number of cells from the three retinal regions from each donor is given in Supplementary Table 35. snRNA-seq data from RGCs from a few additional donors were added to the data set, given the relevance of RGCs to glaucoma, though RGCs still only comprised about 1/250 of the total data set. Differential gene expression for each cell type in the ml_class level used in this study was computed using the Wilcoxon rank sum test.

### Macula

Macular samples were dissected with 8mm punches from five donors within 4 hours of death at the University of Utah (Supplementary Table 36). For three of the samples, RGCs were enriched by staining the nuclei with NEUN antibody (Millipore Sigma, #FCMAB317PE) followed by FACS sorting. The macular samples were processed similarly to the optic nerve head samples below. Differential gene expression for each cell type compared to all other cell types was computed using the MAST method^105^.

### Optic nerve head and posterior tissues

The optic nerve head, including peripapillary tissues, was dissected with 4 mm punches from 13 donors, the optic nerve was dissected from 7 donors, peripapillary sclera from 4 donors, sclera from 3 donors, and choroid from 5 donors, within a median of 6 hours from death at either the University of Utah or Massachusetts General Hospital (Supplementary Table 37). Further description of the tissues’ dissection, single-nuclei isolation, and snRNA-sequencing can be found in Supplementary Note and Monavarfeshani *et al.*^37^. snRNA-seq data processing and analyses were performed similarly to the pipeline used for the anterior segment in van Zyl *et al.*^36^. Thirty-six cell type clusters were identified across the five tissues. Differential gene expression (DGE) was computed using the MAST method^105^, comparing the cells from each cell type to all other cells, excluding cells from the same cell class similar to the cell type of interest, aside for the given cell type (e.g., excluding all fibroblast cell types when computing DGE for cell type, 5-Fibro).

### Cell type-specific enrichment of genes that map to GWAS loci for a given complex trait using ECLIPSER

To identify ocular cell types that are enriched for cell type-specific expression of genes mapped to GWAS loci of POAG, IOP and related traits, we extended a method we recently developed called ECLIPSER (Enrichment of Causal Loci and Identification of Pathogenic cells in Single Cell Expression and Regulation data)^38, 39^, to target genes of colocalizing e/sQTLs. ECLIPSER assesses whether genes mapped to a set of GWAS loci for a given complex disease or trait are enriched for cell type-specific expression compared to the cell type specificity of genes mapped to a background (null) set of GWAS loci associated with hundreds of unrelated traits. The underlying assumption of ECLIPSER is that multiple (though not necessarily all) trait-associated genes will be more highly expressed in a given pathogenic cell type compared to non-pathogenic cell types in a tissue of action, more so than unrelated traits. The analysis consisted of the following main steps: **(i) Mapping genes to GWAS loci.** For the POAG and IOP traits, e/sQTL colocalization analysis was used to prioritize genes in GWAS loci. For the cornea-related, VCDR and negative control traits, genes were mapped to GWAS loci if they were target genes of a GTEx or retina e/sQTL that was in LD (r^2^>0.8) with the GWAS locus (since colocalization analysis for these traits was beyond the scope of this paper). The genome-wide significant variants associated with the cornea traits and negative control traits were taken from Open Targets Genetics^106^, and for physician and machine learning-based VCDR measures from the corresponding published GWAS meta-analyses^12, 13^. **(ii) Null set of GWAS loci.** We compiled a null set of GWAS loci, by selecting all genome-wide significant associations for a range of complex traits in Open Targets Genetics^106^ that were taken from the NHGRI-EBI GWAS catalog and UK Biobank GWAS studies. We excluded from the null set variants associated with any ocular trait. **(iii) LD clumping of loci.** We collapsed GWAS variants that were in LD with each other (r^2^>0.8) or that shared a mapped gene into a single locus for the set of GWAS loci for each ocular or negative control trait and for the null set, separately, to avoid inflating the cell type enrichment results due to LD^39^. **(iv) Cell type specificity locus score.** We scored each GWAS locus for the ocular traits and the null set as the fraction of genes mapped to the locus, that demonstrated cell type specificity (defined here as fold-change > 1.3 and FDR < 0.1). Only genes expressed in at least 5% of cells in any cell type cluster were included in the analysis. **(v) Assessing cell type-specificity of GWAS locus set.** We estimated a cell type specificity fold-enrichment and p-value per trait (GWAS locus set), tissue and cell type combination, compared to the null GWAS locus set, using a Bayesian Fisher’s exact test and the 95th percentile of the null locus scores for the cell type specificity cutoff. The Bayesian approach enables estimating 95% confidence intervals of the fold-enrichment, including for traits that have few or no loci that fall above the enrichment cutoff^39^. **(vi) Cell type specific disease-contributing genes.** Cell type-specific genes mapped to GWAS loci whose score was equal to or above the 95th percentile enrichment cutoff in significantly enriched cell types (‘leading edge loci’) were proposed to influence the given complex trait in the given cell type (‘leading edge genes’), though it is possible that some of these genes are affecting the given trait through other cell types. Cell types with a tissue-wide Benjamini-Hochberg FDR equal to or below 0.1, correcting for the number of cell types tested per tissue, were considered significantly enriched for genes associated with a given trait. To test the specificity of ECLIPSER, we applied the method to eight negative control traits listed in Supplementary Table 45. To assess the robustness of the cell type enrichment results with ECLIPSER, we ran two additional cell type enrichment methods of GWAS data that consider genome-wide genetic associations beyond genome-wide significant loci: stratified LD score regression^25^ and MAGMA^107^ (see below).

### Cell type specific heritability enrichment of disease associations using stratified LD score regression

We applied stratified LD score regression (S-LDSC)^25^ (v1.0.1; URLs) to the GWAS summary statistics of the POAG cross-ancestry meta-analysis, POAG European subset meta-analysis and IOP meta-analysis, and the four single-nucleus differential gene expression datasets described above, to evaluate the contribution of genetic variation in cell type-specific genes to trait heritability. Common variants (MAF>1%) within or near genes specifically expressed in the different cell types (fold-change > 1.1 and FDR<0.1) in each of the four single-nucleus eye tissue datasets described above, were considered in the S-LDSC analysis. A 100 kb windows on either side of each gene was used. The European samples in 1000 Genomes Project Phase 3^96^ were used as the reference panel for computing the LD scores for all three GWAS meta-analyses. Heritability enrichment per cell type was considered significant at Benjamini-Hochberg FDR below 0.1.

### MAGMA gene-association correlation with cell type gene expression

We applied the regression-based model MAGMA (v1.10)^107^ to the POAG cross-ancestry, POAG European subset, and IOP GWAS meta-analyses and the four single-nucleus ocular expression datasets described above, which tests for association between gene association z-scores and average gene expression per cell type, controlling for average gene expression across all cell types per tissue. Gene-based association z-scores were computed for each GWAS based on the most significant variant (SNP-wise=top) within 100kb around each gene, as described in de Leeuw *et al.*^108^. The European samples in 1000 Genomes Project Phase 3^96^ were used as the reference panel for the POAG EUR and IOP GWAS, while all five populations (EUR, AFR, AMR, EAS, SAS) were used for the POAG cross-ancestry GWAS. Significance was determined at Benjamini-Hochberg FDR below 0.1. We applied conditional analysis to all pairwise combinations of nominally significant (P<0.05) cell types within a given tissue, to identify cell types whose trait association signals are independent of the other significant cell type^107^. A proportional significance (PS) of the conditional P-value of a cell type relative to its marginal P-value was computed for each cell type in each cell type pair. Two cell types in a given pair with PS≥0.8 were considered independently associated cell types, and a pair of cell types with PS≥0.5 were considered partial-joint associations. In the case where one cell type had PS≥0.5 and the second cell type a conditional P-value ≥0.05, the first cell type was retained and the second cell type was considered completely dependent on the association of the first cell type. For more details see: https://fuma.ctglab.nl/tutorial#celltype.

### Gene set enrichment analysis of POAG and IOP associated genes

We used *GeneEnrich*^24, 26^ to test whether genes proposed to affect POAG risk or IOP variation cluster in specific biological processes or mouse phenotype ontologies. *GeneEnrich* assesses enrichment of a set of genes of interest in biological pathways or other types of biologically meaningful gene sets, using a hypergeometric distribution and permutation analysis. To account for biases that could arise from the set of genes expressed in a given tissue, an empirical gene set enrichment P-value was computed as the fraction of 1,000 to 100,000 *k* randomly sampled genes (*k* = number of significant genes, e.g., colocalizing e/sGenes) from all genes expressed in the given tissue (background set) that have a hypergeometric probability equal to or higher than that of the significant list of genes. Given the high LD in the HLA region on chromosome 6 (chr6:28510120-33480577) we removed all genes in this region from the gene set enrichment analysis, unless noted otherwise.

We applied *GeneEnrich* to three groups of POAG and IOP associated genes: (i) All 228, 118, and 279 unique target genes of eQTLs and sQTLs that colocalized with POAG cross-ancestry, POAG EUR, and IOP GWAS loci, respectively. Given that the colocalizing e/sQTLs were derived from the different GTEx tissues and retina, we used all genes expressed in any of the 49 GTEx tissues and retina as the background set of genes, and did not correct for expression levels given the differences in expression levels between the tissues. (ii) Sets of POAG and IOP colocalizing genes that were enriched in specific cell types in the eye tissues based on ECLIPSER analysis (tissue-wide FDR ≤ 0.1). For the background sets of genes, we chose all genes expressed in the GTEx or retina tissue that was most relevant for the enriched cell type (e.g., Brain for Optic nerve head; full list in Supplementary Table 39). Given that the expression levels in a tissue may not fully reflect the expression levels in the particular cell type, we did not correct for expression levels in the gene set enrichment analysis of the cell type-specific gene sets. (iii) Target genes of e/sQTLs (FDR<0.05) with top ranked POAG or IOP GWAS P-values (P < 0.05) in tissues whose e/sQTLs were enriched for trait associations based on *QTLEnrich*. Given that e/sQTLs in most tissues displayed significant enrichment, a selected set of QTL/tissue-trait pairs was chosen for gene set enrichment analysis based on the tissue having a top ranked adjusted fold-enrichment and consisting of cell types that may be relevant to glaucoma pathophysiology, such as cells cultured fibroblasts, brain, and artery (Supplementary Tables 4-5). The background sets of genes were defined as all genes expressed in the given tissue excluding the target genes of e/sQTLs with GWAS P<0.05. The expression levels of the randomly sampled genes from the background set in the permutation analysis were matched on the expression levels of the significant set of genes.

We applied *GeneEnrich* to over 11,000 gene sets from four databases downloaded from MSigDB (URLs): Gene Ontology (GO) with three domains: biological processes, molecular function, and cellular components; Reactome; Kyoto Encyclopedia of Genes and Genomes (KEGG); and mouse phenotype ontology gene sets from the Mouse Genome Informatics (MGI). Only gene sets with 10 to 1000 genes were tested, and only genes that were found in the given database were included in the analysis. Statistical significance was determined using a Benjamini Hochberg FDR below 0.1 per database, given extensive gene set overlap between databases. Gene sets with empirical gene set enrichment below 0.05 were considered nominally significant.

### Conditional analysis of *MYOC* POAG locus

Given our finding of significant colocalization of a *PIGC* sQTL with the POAG cross-ancestry association signal in the GWAS locus rs74315329, whose lead variant is a nonsense mutation in the *MYOC* gene, we tested whether there was a secondary independent signal in this locus that might colocalize with the *PIGC* sQTL. We performed association testing on all variants on chromosome 1 conditioning on rs74315329, the lead POAG GWAS variant in the locus by applying the tool COJO (URLs) to the POAG cross-ancestry GWAS meta-analysis summary statistics on chromosome 1. To maintain the *MYOC* lead variant in the initial association testing we filtered out variants with MAF<0.0001. The effective sample size of the POAG cross-ancestry GWAS was computed based on the equation: 4/[(1/Ncases)+(1/Ncontrols)]^109^, which yielded N=124,531 for the POAG GWAS cross-ancestry meta-analysis^8^. For the variant allele frequencies required as input to COJO, we used the European, African and East Asian samples in 1000 Genomes Project^96^, as the POAG GWAS meta-analysis is comprised of these three ancestral groups. eCAVIAR and *enloc* were applied to the residual statistics in the *MOYC* locus from the conditional analysis and all overlapping e/sQTLs from the GTEx tissues and retina. To remove potential false positives, we filtered out variant, gene, tissue, trait combinations if the e/sVariant with a significant colocalization result had a GWAS p-value above 2×10^-5^ or an e/sQTL p-value above 1×10^-4^ and/or an FDR above 0.05 (FALSE in column ‘Pass_QC_QTL_FDR05_P1E04_GWAS_P2E05’ in Supplementary Tables 25 and 27). We used a slightly more lenient GWAS p-value cutoff for the conditional analysis (P<2×10^-5^ compared to P<1×10^-5^ used for the original GWAS summary statistics) given the reduced association power of conditional analysis.

## Data availability

All GTEx protected data are available through the database of Genotypes and Phenotypes (dbGaP) (accession no. phs000424.v8). The GTEx eQTL and sQTL and EyeGEx retina eQTL summary statistics are available on the GTEx portal (https://gtexportal.org/home/datasets). The snRNA-seq data for the anterior segment and macula are available in Gene Expression Omnibus (GEO) accession number GSE199013, for the optic nerve head and posterior tissues in GSE236566, and for the retina in GSE226108. The processed data of the anterior and posterior segments can be visualized in the Broad Institute’s Single Cell Portal at https://singlecell.broadinstitute.org/single_cell/study/SCP1841 and https://singlecell.broadinstitute.org/single_cell/study/SCP2298. The retina Hi-C data is accessible in GEO accession number GSE202471. The GWAS summary statistics for the POAG cross-ancestry GWAS meta-analysis and European subset meta-analysis are accessible in GEO under accession numbers GCST90011770 and GCST90011766, respectively, and for IOP are available from the corresponding publication (Khawaja *et al.,* Nature Genetics 2018). The GWAS loci for complex traits analyzed in this study were downloaded from Open Targets Genetics (https://genetics.opentargets.org/). The gene sets taken from MSigDB were downloaded from: http://www.gsea-msigdb.org/gsea/msigdb/collections.jsp, and the mouse phenotype ontology gene sets from the Mouse Genome Informatics (MGI) website (http://www.informatics.jax.org/).

All data produced in the current study are either contained in the manuscript, deposited in online databases as stated in the manuscript, or can be obtained upon request to the authors. All GTEx protected data are available through the database of Genotypes and Phenotypes (dbGaP) (accession no. phs000424.v8). The single nucleus RNA-sequencing data of the anterior and posterior eye tissues are available in Gene Expression Omnibus (GEO) accession numbers GSE199013, GSE236566, and GSE226108. The retina Hi-C data is accessible in GEO accession number GSE202471. The GWAS summary statistics for the POAG cross-ancestry GWAS meta-analysis and European subset meta-analysis are accessible in GEO accession numbers GCST90011770 and GCST90011766, respectively, and for IOP GWAS are available from the corresponding publication (Khawaja et al., Nature Genetics 2018).

## Code Availability

The code of all tools used for analyses in this paper are publicly available and are listed in the URLs below. Custom code used to generate some of the plots are available upon request.

## URLs

GTEx: https://gtexportal.org/home/datasets EyeGEx: https://gtexportal.org/home/datasets

QTLEnrich v2: https://github.com/segrelabgenomics/QTLEnrich

GeneEnrich v2: https://github.com/segrelabgenomics/GeneEnrich

MSigDB: http://www.gsea-msigdb.org/gsea/msigdb/collections.jsp

PLINK: https://www.cog-genomics.org/plink/

eCAVIAR: https://github.com/fhormoz/caviar

fastEnloc: https://github.com/xqwen/fastenloc

DAP-G: https://github.com/xqwen/dap/tree/master/dap_src

ECLIPSER: https://github.com/segrelabgenomics/ECLIPSER

MAGMA v1.10: https://ctg.cncr.nl/software/magma, https://fuma.ctglab.nl/tutorial#celltype

S-LDSC v1.0.1: https://github.com/bulik/ldsc

genomAD: https://gnomad.broadinstitute.org/

QMplot: https://github.com/ShujiaHuang/qmplot

LocusCompare: https://github.com/boxiangliu/locuscomparer

## Acknowledgements

We thank William Wen for helpful discussions on the interpretation of the colocalization results. We thank members of the Segrè lab for valuable comments and feedback. This work was funded by NIH/NEI R01 EY031424-01 (AVS, ARH, JR, PAM), NIH/NEI P30 EY014104 (JLW, AVS), NIH/NEI EY032559-01 (JLW, AVS), and the Chan Zuckerberg Initiative (CZI) Seed Network for the Human Cell Atlas awards CZF2019-002459 (JRS, AVS, WY, AM) and CZF2019-002425 (QL, RC). XJ is supported by the University of Edinburgh and University of Helsinki joint PhD studentship program in Human Genomics. VV is supported by an MRC University Unit Programme grant (MC_UU_00007/10) (QTL in Health and Disease). APK is supported by a UK Research and Innovation Future Leaders Fellowship, an Alcon Research Institute Young Investigator Award and a Lister Institute for Preventive Medicine Award. This research was supported by the NIHR Biomedical Research Centre at Moorfields Eye Hospital and the UCL Institute of Ophthalmology.

## References

1. Tham, Y.-C. et al. Global prevalence of glaucoma and projections of glaucoma burden through 2040: a systematic review and meta-analysis. Ophthalmology 121, 2081–2090 (2014).

2. Weinreb, R. N. et al. Primary open-angle glaucoma. Nat Rev Dis Primers 2, 16067 (2016).

3. Leske, M. C. et al. Predictors of long-term progression in the early manifest glaucoma trial. Ophthalmology 114, 1965–1972 (2007).

4. Kwon, Y. H., Fingert, J. H., Kuehn, M. H. & Alward, W. L. M. Primary open-angle glaucoma. N. Engl. J. Med. 360, 1113–1124 (2009).

5. Kim, J. et al. Impaired angiopoietin/Tie2 signaling compromises Schlemm’s canal integrity and induces glaucoma. J. Clin. Invest. 127, 3877–3896 (2017).

6. Costagliola, C. et al. How many aqueous humor outflow pathways are there? Surv. Ophthalmol. 65, 144–170 (2020).

7. Anderson, D. R., Drance, S. M., Schulzer, M. & Collaborative Normal-Tension Glaucoma Study Group. Natural history of normal-tension glaucoma. Ophthalmology 108, 247–253 (2001).

8. Gharahkhani, P. et al. Genome-wide meta-analysis identifies 127 open-angle glaucoma loci with consistent effect across ancestries. Nat. Commun. 12, 1258 (2021).

9. Khawaja, A. P. et al. Genome-wide analyses identify 68 new loci associated with intraocular pressure and improve risk prediction for primary open-angle glaucoma. Nature Genetics vol. 50 778–782 Preprint at https://doi.org/10.1038/s41588-018-0126-8 (2018).

10. MacGregor, S. et al. Genome-wide association study of intraocular pressure uncovers new pathways to glaucoma. Nature Genetics vol. 50 1067–1071 Preprint at https://doi.org/10.1038/s41588-018-0176-y (2018).

11. Gao, X. R. et al. Genome-wide association analyses identify new loci influencing intraocular pressure. Human Molecular Genetics vol. 27 2205–2213 Preprint at https://doi.org/10.1093/hmg/ddy111 (2018).

12. Alipanahi, B. et al. Large-scale machine-learning-based phenotyping significantly improves genomic discovery for optic nerve head morphology. Am. J. Hum. Genet. 108, 1217–1230 (2021).

13. Springelkamp, H. et al. New insights into the genetics of primary open-angle glaucoma based on meta-analyses of intraocular pressure and optic disc characteristics. Hum. Mol. Genet. 26, 438–453 (2017).

14. Lu, Y. et al. Common genetic variants near the Brittle Cornea Syndrome locus ZNF469 influence the blinding disease risk factor central corneal thickness. PLoS Genet. 6, e1000947 (2010).

15. Vitart, V. et al. New loci associated with central cornea thickness include COL5A1, AKAP13 and AVGR8. Hum. Mol. Genet. 19, 4304–4311 (2010).

16. Hoehn, R. et al. Population-based meta-analysis in Caucasians confirms association with COL5A1 and ZNF469 but not COL8A2 with central corneal thickness. Hum. Genet. 131, 1783–1793 (2012).

17. Gao, X. et al. A genome-wide association study of central corneal thickness in Latinos. Invest. Ophthalmol. Vis. Sci. 54, 2435–2443 (2013).

18. Iglesias, A. I. et al. Cross-ancestry genome-wide association analysis of corneal thickness strengthens link between complex and Mendelian eye diseases. Nat. Commun. 9, 1864 (2018).

19. Gao, X. et al. Genome-wide association study identifies WNT7B as a novel locus for central corneal thickness in Latinos. Hum. Mol. Genet. 25, 5035–5045 (2016).

20. Fan, B. J. et al. Family-Based Genome-Wide Association Study of South Indian Pedigrees Supports WNT7B as a Central Corneal Thickness Locus. Invest. Ophthalmol. Vis. Sci. 59, 2495–2502 (2018).

21. Ivarsdottir, E. V. et al. Sequence variation at ANAPC1 accounts for 24% of the variability in corneal endothelial cell density. Nat. Commun. 10, 1284 (2019).

22. Simcoe, M. J., Khawaja, A. P., Hysi, P. G., Hammond, C. J. & UK Biobank Eye and Vision Consortium. Genome-wide association study of corneal biomechanical properties identifies over 200 loci providing insight into the genetic etiology of ocular diseases. Hum. Mol. Genet. 29, 3154–3164 (2020).

23. Han, X. et al. Automated AI labeling of optic nerve head enables insights into cross-ancestry glaucoma risk and genetic discovery in >280,000 images from UKB and CLSA. Am. J. Hum. Genet. 108, 1204–1216 (2021).

24. Gamazon, E. R. et al. Using an atlas of gene regulation across 44 human tissues to inform complex disease- and trait-associated variation. Nat. Genet. 50, 956–967 (2018).

25. Finucane, H. K. et al. Heritability enrichment of specifically expressed genes identifies disease-relevant tissues and cell types. Nat. Genet. 50, 621–629 (2018).

26. GTEx Consortium. The GTEx Consortium atlas of genetic regulatory effects across human tissues. Science 369, 1318–1330 (2020).

27. Ratnapriya, R. et al. Retinal transcriptome and eQTL analyses identify genes associated with age-related macular degeneration. Nat. Genet. 51, 606–610 (2019).

28. Strunz, T. et al. A mega-analysis of expression quantitative trait loci in retinal tissue. PLoS Genet. 16, e1008934 (2020).

29. Orozco, L. D. et al. Integration of eQTL and a single-cell atlas in the human eye identifies causal genes for age-related macular degeneration. Cell Rep. 30, 1246–1259.e6 (2020).

30. Liu, B. et al. Genetic analyses of human fetal retinal pigment epithelium gene expression suggest ocular disease mechanisms. *Commun*. Biol. 2, 186 (2019).

31. Yan, W. et al. Cell Atlas of The Human Fovea and Peripheral Retina. Sci. Rep. 10, 9802 (2020).

32. Dharmat, R., Kim, S., Li, Y. & Chen, R. Single-Cell Capture, RNA-seq, and Transcriptome Analysis from the Neural Retina. Methods Mol. Biol. 2092, 159–186 (2020).

33. Liang, Q. et al. A multi-omics atlas of the human retina at single-cell resolution. Cell Genom. 3, 100298 (2023).

34. van Zyl, T. et al. Cell atlas of aqueous humor outflow pathways in eyes of humans and four model species provides insight into glaucoma pathogenesis. Proc. Natl. Acad. Sci. U. S. A. 117, 10339–10349 (2020).

35. Patel, G. et al. Molecular taxonomy of human ocular outflow tissues defined by single-cell transcriptomics. Proc. Natl. Acad. Sci. U. S. A. 117, 12856–12867 (2020).

36. van Zyl, T. et al. Cell atlas of the human ocular anterior segment: Tissue-specific and shared cell types. Proc. Natl. Acad. Sci. U. S. A. 119, e2200914119 (2022).

37. Monavarfeshani, A., et al. Transcriptomic Analysis of the Ocular Posterior Segment Completes a Cell Atlas of the Human Eye. bioRxiv (2023) doi:10.1101/2023.04.26.538447.

38. Eraslan, G. et al. Single-nucleus cross-tissue molecular reference maps toward understanding disease gene function. Science 376, eabl4290 (2022).

39. Rouhana, J., J. Wang, G. Eraslan, S. Anand, A. Hamel, B. Cole, A. Regev, F. Aguet, K. Ardlie, and A. V. Segrè. ECLIPSER: identifying causal cell types and genes for complex traits through single cell enrichment of e/sQTL-mapped genes in GWAS loci. BioRxiv (2021) doi:10.1101/2021.11.24.469720.

40. Marchal, C. et al. High-resolution genome topology of human retina uncovers super enhancer-promoter interactions at tissue-specific and multifactorial disease loci. Nat. Commun. 13, 5827 (2022).

41. Hormozdiari, F. et al. Colocalization of GWAS and eQTL Signals Detects Target Genes. Am. J. Hum. Genet. 99, 1245–1260 (2016).

42. Wen, X., Pique-Regi, R. & Luca, F. Integrating molecular QTL data into genome-wide genetic association analysis: Probabilistic assessment of enrichment and colocalization. PLoS Genet. 13, e1006646 (2017).

43. Thomson, B. R. et al. Angiopoietin-1 is required for Schlemm’s canal development in mice and humans. J. Clin. Invest. 127, 4421–4436 (2017).

44. Wiggs, J. L. et al. Common variants at 9p21 and 8q22 are associated with increased susceptibility to optic nerve degeneration in glaucoma. PLoS Genet. 8, e1002654 (2012).

45. Collantes, E. R. A. et al. EFEMP1 rare variants cause familial juvenile-onset open-angle glaucoma. Hum. Mutat. 43, 240–252 (2022).

46. Wiggs, J. L. & Pasquale, L. R. Genetics of glaucoma. Hum. Mol. Genet. 26, R21–R27 (2017).

47. Lewczuk, K., Jabłońska, J., Konopińska, J., Mariak, Z. & Rękas, M. Schlemm’s canal: the outflow “vessel.” Acta Ophthalmol. (2021) doi:10.1111/aos.15027.

48. Barbeira, A. N. et al. Exploiting the GTEx resources to decipher the mechanisms at GWAS loci. Genome Biol. 22, 49 (2021).

49. Zhu, Z. et al. Integration of summary data from GWAS and eQTL studies predicts complex trait gene targets. Nat. Genet. 48, 481–487 (2016).

50. Hemani, G., Bowden, J. & Davey Smith, G. Evaluating the potential role of pleiotropy in Mendelian randomization studies. Hum. Mol. Genet. 27, R195–R208 (2018).

51. Zuber, V. et al. Combining evidence from Mendelian randomization and colocalization: Review and comparison of approaches. Am. J. Hum. Genet. 109, 767–782 (2022).

52. Hukku, A. et al. Probabilistic colocalization of genetic variants from complex and molecular traits: promise and limitations. Am. J. Hum. Genet. 108, 25–35 (2021).

53. Wallace, C. Eliciting priors and relaxing the single causal variant assumption in colocalisation analyses. PLoS Genet. 16, e1008720 (2020).

54. GTEx Consortium et al. Genetic effects on gene expression across human tissues. Nature 550, 204–213 (2017).

55. Claussnitzer, M. et al. FTO Obesity Variant Circuitry and Adipocyte Browning in Humans. N. Engl. J. Med. 373, 895–907 (2015).

56. Nasser, J. et al. Genome-wide enhancer maps link risk variants to disease genes. Nature 593, 238–243 (2021).

57. Cao, J. et al. Joint profiling of chromatin accessibility and gene expression in thousands of single cells. Science 361, 1380–1385 (2018).

58. Mackay, D. S., Bennett, T. M. & Shiels, A. Exome Sequencing Identifies a Missense Variant in EFEMP1 Co-Segregating in a Family with Autosomal Dominant Primary Open-Angle Glaucoma. PLoS One 10, e0132529 (2015).

59. Lim, S.-H. et al. CYP1B1, MYOC, and LTBP2 mutations in primary congenital glaucoma patients in the United States. Am. J. Ophthalmol. 155, 508–517.e5 (2013).

60. Fuchshofer, R. & Tamm, E. R. The role of TGF-β in the pathogenesis of primary open-angle glaucoma. Cell Tissue Res. 347, 279–290 (2012).

61. Scerbo, P. & Monsoro-Burq, A. H. The vertebrate-specific VENTX/NANOG gene empowers neural crest with ectomesenchyme potential. Sci Adv 6, eaaz1469 (2020).

62. Kumar, S., Kumar, V., Li, W. & Kim, J. Ventx Family and Its Functional Similarities with Nanog: Involvement in Embryonic Development and Cancer Progression. Int. J. Mol. Sci. 23, (2022).

63. Fadini, G. P. et al. Reduced endothelial progenitor cells and brachial artery flow-mediated dilation as evidence of endothelial dysfunction in ocular hypertension and primary open-angle glaucoma. Acta Ophthalmol. 88, 135–141 (2010).

64. Su, W.-W. et al. Glaucoma is associated with peripheral vascular endothelial dysfunction. Ophthalmology 115, 1173–1178.e1 (2008).

65. Green, K. J., Getsios, S., Troyanovsky, S. & Godsel, L. M. Intercellular junction assembly, dynamics, and homeostasis. Cold Spring Harb. Perspect. Biol. 2, a000125 (2010).

66. Pattabiraman, P. P., Epstein, D. L. & Rao, P. V. Regulation of Adherens Junctions in Trabecular Meshwork Cells by Rac GTPase and their influence on Intraocular Pressure. J. Ocul. Biol. Dis. Infor. 1, (2013).

67. Gould, D. B., Smith, R. S. & John, S. W. M. Anterior segment development relevant to glaucoma. Int. J. Dev. Biol. 48, 1015–1029 (2004).

68. Ronen E. Mukamel, Robert E. Handsaker, Maxwell A. Sherman, Alison R. Barton, Margaux L. A. Hujoel, Steven A. McCarroll, Po-Ru Loh. Repeat polymorphisms in non-coding DNA underlie top genetic risk loci for glaucoma and colorectal cancer. bioRxiv (2022) doi:10.1101/2022.10.11.22280955.

69. Pasquale, L. R. et al. CDKN2B-AS1 genotype-glaucoma feature correlations in primary open-angle glaucoma patients from the United States. Am. J. Ophthalmol. 155, 342–353.e5 (2013).

70. Kim, B. J. & Scott, D. A. RERE deficiency causes retinal and optic nerve atrophy through degeneration of retinal cells. Dev. Dyn. 250, 1398–1409 (2021).

71. Wang, H. et al. Physiological function of myocilin and its role in the pathogenesis of glaucoma in the trabecular meshwork (Review). Int. J. Mol. Med. 43, 671–681 (2019).

72. Finucane, H. K. et al. Partitioning heritability by functional annotation using genome-wide association summary statistics. Nat. Genet. 47, 1228–1235 (2015).

73. Watanabe, K., Taskesen, E., van Bochoven, A. & Posthuma, D. Functional mapping and annotation of genetic associations with FUMA. Nat. Commun. 8, 1826 (2017).

74. Jagadeesh, K. A., et al. Identifying disease-critical cell types and cellular processes across the human body by integration of single-cell profiles and human genetics. *bioRxiv* 2021.03.19.436212 (2021) doi:10.1101/2021.03.19.436212.

75. Slowikowski, K., Hu, X. & Raychaudhuri, S. SNPsea: an algorithm to identify cell types, tissues and pathways affected by risk loci. Bioinformatics 30, 2496–2497 (2014).

76. Calderon, D. et al. Inferring Relevant Cell Types for Complex Traits by Using Single-Cell Gene Expression. Am. J. Hum. Genet. 101, 686–699 (2017).

77. Stamer, W. D. & Acott, T. S. Current understanding of conventional outflow dysfunction in glaucoma. Curr. Opin. Ophthalmol. 23, 135–143 (2012).

78. Alarcon-Martinez, L. et al. Pericyte dysfunction and loss of interpericyte tunneling nanotubes promote neurovascular deficits in glaucoma. Proc. Natl. Acad. Sci. U. S. A. 119, (2022).

79. Wang, R., Seifert, P. & Jakobs, T. C. Astrocytes in the Optic Nerve Head of Glaucomatous Mice Display a Characteristic Reactive Phenotype. Invest. Ophthalmol. Vis. Sci. 58, 924– 932 (2017).

80. Zhao, X., Sun, R., Luo, X., Wang, F. & Sun, X. The Interaction Between Microglia and Macroglia in Glaucoma. Front. Neurosci. 15, 610788 (2021).

81. Shinozaki, Y. & Koizumi, S. Potential roles of astrocytes and Müller cells in the pathogenesis of glaucoma. J. Pharmacol. Sci. 145, 262–267 (2021).

82. Tovar-Vidales, T., Wordinger, R. J. & Clark, A. F. Identification and localization of lamina cribrosa cells in the human optic nerve head. Exp. Eye Res. 147, 94–97 (2016).

83. Lopez, N. N., Clark, A. F. & Tovar-Vidales, T. Isolation and characterization of human optic nerve head astrocytes and lamina cribrosa cells. Exp. Eye Res. 197, 108103 (2020).

84. Strickland, R. G., Garner, M. A., Gross, A. K. & Girkin, C. A. Remodeling of the lamina cribrosa: Mechanisms and potential therapeutic approaches for glaucoma. Int. J. Mol. Sci. 23, 8068 (2022).

85. Calkins, D. J. Critical pathogenic events underlying progression of neurodegeneration in glaucoma. Prog. Retin. Eye Res. 31, 702–719 (2012).

86. Venkataraman, S. T., Flanagan, J. G. & Hudson, C. Vascular reactivity of optic nerve head and retinal blood vessels in glaucoma--a review. Microcirculation 17, 568–581 (2010).

87. Tamm, E. R., Ethier, C. R. & Lasker/IRRF Initiative on Astrocytes and Glaucomatous Neurodegeneration Participants. Biological aspects of axonal damage in glaucoma: A brief review. Exp. Eye Res. 157, 5–12 (2017).

88. Paula, J. S., O’Brien, C. & Stamer, W. D. Life under pressure: The role of ocular cribriform cells in preventing glaucoma. Exp. Eye Res. 151, 150–159 (2016).

89. Zeng, H.-L. & Shi, J.-M. The role of microglia in the progression of glaucomatous neurodegeneration-a review. Int. J. Ophthalmol. 11, 143–149 (2018).

90. Kim-Hellmuth, S., Aguet, F. & Oliva, M. Cell type–specific genetic regulation of gene expression across human tissues. (2020).

91. van der Wijst, M. et al. The single-cell eQTLGen consortium. Elife 9, (2020).

92. Li, Y. I. et al. Annotation-free quantification of RNA splicing using LeafCutter. Nat. Genet. 50, 151–158 (2018).

93. Pruim, R. J. et al. LocusZoom: regional visualization of genome-wide association scan results. Bioinformatics 26, 2336–2337 (2010).

94. Storey, J. D. & Tibshirani, R. Statistical significance for genomewide studies. Proc. Natl. Acad. Sci. U. S. A. 100, 9440–9445 (2003).

95. Gamazon, E. R., Huang, R. S., Dolan, M. E., Cox, N. J. & Im, H. K. Integrative genomics: quantifying significance of phenotype-genotype relationships from multiple sources of high-throughput data. Front. Genet. 3, 202 (2012).

96. Zheng-Bradley, X. et al. Alignment of 1000 Genomes Project reads to reference assembly GRCh38. Gigascience 6, 1–8 (2017).

97. Smith, G. D. & Ebrahim, S. “Mendelian randomization”: can genetic epidemiology contribute to understanding environmental determinants of disease? Int. J. Epidemiol. 32, 1–22 (2003).

98. Yavorska, O. O. & Burgess, S. MendelianRandomization: an R package for performing Mendelian randomization analyses using summarized data. Int. J. Epidemiol. 46, 1734–1739 (2017).

99. Burgess, S., Small, D. S. & Thompson, S. G. A review of instrumental variable estimators for Mendelian randomization. Stat. Methods Med. Res. 26, 2333–2355 (2017).

100. Burgess, S., Butterworth, A. & Thompson, S. G. Mendelian randomization analysis with multiple genetic variants using summarized data. Genet. Epidemiol. 37, 658–665 (2013).

101. Bowden, J., Davey Smith, G., Haycock, P. C. & Burgess, S. Consistent Estimation in Mendelian Randomization with Some Invalid Instruments Using a Weighted Median Estimator. Genet. Epidemiol. 40, 304–314 (2016).

102. Bowden, J., Davey Smith, G. & Burgess, S. Mendelian randomization with invalid instruments: effect estimation and bias detection through Egger regression. Int. J. Epidemiol. 44, 512–525 (2015).

103. Verbanck, M., Chen, C.-Y., Neale, B. & Do, R. Detection of widespread horizontal pleiotropy in causal relationships inferred from Mendelian randomization between complex traits and diseases. Nat. Genet. 50, 693–698 (2018).

104. Quinlan, A. R. BEDTools: The Swiss-army tool for genome feature analysis. Curr. Protoc. Bioinformatics 47, 11.12.1-34 (2014).

105. Finak, G. et al. MAST: a flexible statistical framework for assessing transcriptional changes and characterizing heterogeneity in single-cell RNA sequencing data. Genome Biol. 16, 278 (2015).

106. Ghoussaini, M. et al. Open Targets Genetics: systematic identification of trait-associated genes using large-scale genetics and functional genomics. Nucleic Acids Res. 49, D1311– D1320 (2021).

107. Watanabe, K., Umićević Mirkov, M., de Leeuw, C. A., van den Heuvel, M. P. & Posthuma, D. Genetic mapping of cell type specificity for complex traits. Nat. Commun. 10, 3222 (2019).

108. de Leeuw, C. A., Mooij, J. M., Heskes, T. & Posthuma, D. MAGMA: generalized gene-set analysis of GWAS data. PLoS Comput. Biol. 11, e1004219 (2015).

109. Pollack, S. et al. Multiethnic Genome-Wide Association Study of Diabetic Retinopathy Using Liability Threshold Modeling of Duration of Diabetes and Glycemic Control. Diabetes 68, 441–456 (2019).

110. Li, B. & Dewey, C. N. RSEM: accurate transcript quantification from RNA-Seq data with or without a reference genome. BMC Bioinformatics 12, 323 (2011).

