## Supplementary Notes for "Integrating genetic regulation and single-cell expression with GWAS prioritizes causal genes and cell types for glaucoma"

**Supplementary Note**

Hamel *et al.,* medRxiv 2023, **doi:** https://doi.org/10.1101/2022.05.14.22275022

**Overview of approach and methods.** We applied the following analytical steps to known GWAS loci and to genome-wide association summary statistics of POAG and its major risk factor, IOP, to identify regulatory mechanisms, genes, pathways, and cell types that may significantly contribute to POAG risk. First, to test the relevance of *cis*-eQTLs and *cis*-sQTLs in non-ocular GTEx tissues to POAG and IOP, we tested whether POAG and IOP genetic associations beyond genome-wide significance were enriched among eQTLs and sQTLs in GTEx tissues and retina, using *QTLEnrich*[1,2] that controls for confounding factors. A lower bound number of e/sQTLs proposed to contribute to POAG risk or IOP variation in significant tissues was estimated using an empirical true positive rate estimation approach implemented in *QTLEnrich*[2] (Fig. 1a and Methods). In cases where significant enrichment of GWAS associations among e/sQTLs was found, the target genes of e/sQTLs with top ranked POAG or IOP GWAS p-values ($P<0.05$) were tested for enrichment in biological processes using *GeneEnrich*[1], to identify new genetic associations and associated pathways (Fig. 1c). Next, to propose putative causal genes that may underlie genome-wide significant POAG and IOP GWAS loci, we applied two colocalization methods, eCAVIAR[3] and *enloc*[4], to the summary statistics of each known GWAS locus and all overlapping *cis*-eQTLs and *cis*-sQTLs in 49 GTEx tissues (v8)[2] and *cis*-eQTLs in peripheral retina[5] (Fig. 1b and Methods). We applied Mendelian randomization (MR) to the significant colocalization results to further identify a high confidence set of e/sQTLs that may be causal to POAG and/or IOP (Fig. 1b and Methods). We also integrated retina Hi-C and epigenetic data with colocalizing e/sQTLs and POAG loci to provide supportive evidence for potential causal regulatory effects on POAG in retina (Fig. 1b and Methods). We applied gene set enrichment analysis (*GeneEnrich*) to the proposed causal genes based on colocalization analysis and known signaling and metabolic pathways, gene ontologies, and mouse phenotype ontologies (Fig. 1c and Methods). Finally, to identify key pathogenic cell types through which the colocalizing genes may be mediating their effect on POAG and IOP, we applied a method we recently developed, ECLIPSER[6,7] to the GWAS-e/sQTL colocalizing genes (Fig. 1d and Methods) and single-nucleus expression data from glaucoma-relevant ocular tissues: six tissues from the anterior segment[8], including the outflow pathways, retina[8,9], and the optic nerve head, optic nerve, and surrounding posterior tissues[10]. ECLIPSER tests whether the expression of genes mapped to GWAS loci is enriched in specific cell types compared to a null distribution of GWAS loci associated with thousands of traits unrelated to the tissue of interest (Methods). To corroborate the cell type enrichment results, we applied two regression-based methods that assess cell type-specificity for complex traits considering all genetic associations genome-wide: stratified-LD score regression (LDSC)[11] and MAGMA[12] (Fig. 1e and Methods). All software and tools developed for this analysis were released on GitHub (see URLs).

**Enrichment of POAG and IOP associations among e/sQTLs.** As the GWAS loci discovered to date for POAG and IOP explain only a small proportion of the traits' heritability, we tested whether eQTLs and sQTLs can help identify new trait associations with modest effects beyond genome-wide significance, not yet detected due to insufficient GWAS sample sizes (Fig. 1a). To this end, we tested whether *cis*-eQTLs and *cis*-sQTLs from 49 GTEx tissues (v8)[2] and peripheral retina eQTLs[5] were enriched for multiple POAG or IOP associations (GWAS $P<0.05$) using *QTLEnrich*[1,2], which adjusts for confounding factors and tissue sample size (Methods). In cases where enrichment was found, we estimated the number of e/sQTLs that may be true associations with POAG or IOP over noise based on an empirical estimation of the true positive rate ($\pi_1$), which we implemented in *QTLEnrich*[2] (see Methods). We found significant enrichment of POAG and IOP associations among both eQTLs and sQTLs in most of the 49 GTEx tissues and retina (Bonferroni-corrected $P<5x10^{-4}$) with stronger enrichment found for IOP compared to POAG (Supplementary Tables 1-3; One-sided Wilcoxon rank sum test $P<1.6x10^{-8}$). For POAG cross-ancestry, the strongest enrichment among eQTLs was found in brain, lymphocyte, cells cultured fibroblasts, esophagus, and artery (Fig. 2a), and among sQTLs in kidney cortex, heart atrial appendage, brain, glands, and adipose (Supplementary Figs. 1-2 and Supplementary Tables 2-3). For IOP, the strongest enrichment among eQTLs was found in brain, adipose, skin, fibroblast, and artery (Fig. 2b), and among sQTLs in brain, glands, and heart atrial appendage (Supplementary Fig. 2 and Supplementary Tables 2-3). eQTLs in retina were also enriched for POAG and IOP associations ($P<1x10^{-5}$), though was not among the top-ranking tissues. While the GTEx tissues are non-ocular tissues, many of the most strongly enriched tissues share cell types that may be pathogenic to glaucoma based on prior knowledge and our GWAS cell type enrichment analysis (Figs. 6-7 and Supplementary Tables 38, 48-49). For example, macroglial cells in brain may share e/sQTLs also present in astrocytes or Müller glia cells in the retina, vasculature cells in artery may be a proxy for the lymphatic vessels in the aqueous outflow pathway, and cultured fibroblasts may share genetic programs with the fibroblast cells in the ocular outflow pathways. In all, this analysis suggests that over a thousand common variants that affect gene expression or alternative splicing are likely to contribute to POAG risk and IOP variation.

**Pathway analysis of e/sQTL target genes of top ranked POAG and IOP associations.** To identify true positive POAG and IOP associations beyond genome-wide significance, over noise, amongst the enriched e/sQTLs, and to propose potentially new causal genes and pathways, we searched for target genes of e/sQTLs with POAG or IOP GWAS P-values below 0.05 that were enriched in biological pathways, gene ontologies, or mouse phenotype ontologies. Our underlying hypothesis was that genes that contribute to complex traits will cluster in a limited set of biological processes. We applied *GeneEnrich*[1] (Methods) to a selected set of tissues per trait that displayed the highest adjusted fold-enrichment, controlling for gene expression levels in the given tissue (Supplementary Tables 2-3). We found significant enrichment of target genes of brain hippocampus sQTLs with top ranked POAG cross-ancestry associations (GWAS P<0.05) in mitochondrial respiratory complex I (FDR<0.05), including NADH dehydrogenase complex assembly and mitochondrial respiratory chain complex assembly, which included newly proposed genes for POAG (e.g., *NDUFB2, NDUFB4, NDUFAF1, NDUFAF2, NDUFAF6,* and *ATP5SL*), and the citric acid TCA cycle (FDR=0.09) (Supplementary Table 4). Target genes of gastroesophageal junction eQTLs with top ranked POAG associations were enriched (FDR<0.1) in metabolic processes, including glycosaminoglycan biosynthesis keratan sulfate and limonene and pinene degradation. Target genes of brain spinal cord eQTLs enriched for IOP associations were enriched in cellular amino acid catabolic metabolism, in particular, histidine metabolism (FDR=0.03), and skin eQTLs with top ranked IOP associations were enriched in dynactin binding that is involved in moving membrane vesicles along microtubules (FDR=0.07) (Supplementary Table 5). This analysis identified novel genes that may influence POAG risk or IOP via regulatory effects (listed in Supplementary Tables 4-5), functioning primarily in metabolic processes. Examining other types of functional relationships, such as co-regulation or protein-protein interactions, might help identify additional glaucoma associations and genes.

**Noncoding genes proposed as contributors to POAG risk.** While the majority of colocalizing e/sGenes are protein coding (60-72% per trait), noncoding RNA genes make up 18-20% of the proposed causal genes for POAG and IOP loci (Fig. 2f and Supplementary Table 17), half of which are lincRNAs and half antisense genes. Noncoding genes were proposed as a sole potential causal gene for 10 POAG or IOP loci, including the lincRNA *LINC01948* in POAG cross-ancestry locus rs112142644 on chromosome 5, and *RP11-217B7.2*, a lnc-ABCA1-1:1 lincRNA, in the shared POAG and IOP locus on chromosome 10 (rs2472494 and rs2472493, respectively) (Supplementary Table 13). We further found that e/sQTLs acting on 14 pairs of protein coding genes and their corresponding antisense gene colocalized in the same GWAS locus, including *HLA-F* and *HLA-F-AS1* with POAG risk, *LPP* and *LPP-AS2* with IOP levels, and *MAPT* and *MAPT-AS1* with both POAG and IOP (Supplementary Table 18).

**Secondary signal found in Myocilin locus for POAG**. The POAG cross-ancestry GWAS association with the largest odds ratio (OR=5.47) rs74315329 (chr1:171636338:G:A), a nonsense mutation (p.Gln368Ter) in *MYOC*, is European-specific. Its minor allele frequency in non-Finnish European in gnomAD is 0.00126, in African American is 0.00039, and in East and South Asian populations is less than 0.0001 based on gnomAD (URLs). We tested all e/sQTL gene-tissue pairs that overlapped this locus targeting 24 genes identified an sQTL (chr1:172398537:A:G) acting on *PIGC,* phosphatidylinositol glycan anchor biosynthesis class C, in spleen with eCAVIAR (CLPP=0.12) or artery tissue with *enloc* (artery tibial, RCP=0.34; heart atrial appendage RPC=0.26) that significantly colocalized with the POAG cross-ancestry locus (splicing event: chr1:172370311-172376554; Supplementary Table 13 and Supplementary Fig. 13). Since the nonsense variant in *MYOC* is likely the primary causal variant in the locus, we tested whether a secondary haplotype may exist in the locus and, if so, if it colocalizes with the *PIGC* sQTL. We performed association testing of all variants in the GWAS locus LD interval conditioning on the lead POAG variant rs74315329 using COJO (URLs; Methods), and found two low frequency, intronic variants that passed genome-wide significance (chr1:171531082:A:G EUR MAF=0.0036 and chr1:170716994:A:C EUR MAF=0.002) and another LD-independent signal (chr1:172398537:A:G, POAG GWAS conditional $P=1.08\times10^{-6}$, EUR MAF=0.075 in gnomAD) that passed Bonferroni correction for the number of variants tested in the locus ($P<5.87\times10^{-6}$) (Supplementary Table 24). We next tested whether the residuals of this secondary signal, which are independent of the locus' lead variant rs74315329, colocalized with any of the e/sQTL-tissue pairs in the locus using eCAVIAR and *enloc*. We found suggestive significance (CLPP=0.009) for the POAG LD-independent signal chr1:172398537:A:G and the same *PIGC* sQTL in spleen as above (chr1:172370311-172376554), as well as for a *PRRX1* eQTL (CLPP=0.01) with eCAVIAR (Supplementary Tables 25-26), and significant colocalization for the same *PIGC* sQTL in several tissues with *enloc* (maximum RCP=0.13) (Supplementary Tables 27-28). *Enloc* also found significant POAG colocalization for another sQTL acting on *PIGC* (max RCP=0.22, chr1:172376636-17237813) and for *PIGC* eQTLs (RCP=0.14) in several tissues (Supplementary Tables 27-28). These results suggest that decreased exon 2 skipping in *PIGC* (Supplementary Fig. 13) or increased *PIGC* expression may lead to increased POAG risk.

**Enrichment of POAG genes in retinal macroglial cells is IOP-independent.** While the expression of POAG cross-ancestry colocalizing e/sGenes were significantly enriched in astrocytes and Müller glia cells in retina (FDR<0.04; Supplementary Table 38, Fig. 7a and Supplementary Fig. 16a), IOP genes were only nominally enriched in astrocytes (P=0.032; Supplementary Fig. 16c). We assessed the extent to which the enrichment of POAG genes in retinal astrocytes was driven by IOP-independent associations. Only one third of the 12 POAG genes driving astrocyte enrichment in retina were common with IOP (*DGKG, FMNL2, GAS7, LPP*) (Supplementary Fig. 16d, f). Furthermore, when applying ECLIPSER to POAG-only or IOP-only loci or shared loci, significant enrichment (FDR<0.08) was found in astrocytes and Müller glia cells in retina and macula solely for POAG-only and shared loci, but not for IOP-only loci (Supplementary Table 41 and Supplementary Fig. 17), suggesting an IOP-independent effect of astrocytes and Müller glia cells on glaucoma. The POAG genes driving the enrichment in astrocytes were nominally enriched in diacylglycerol metabolic process ($P=1.8x10^{-4}$), negative regulation of fat cell differentiation ($P=7.7x10^{-4}$) and regulation of hematopoietic stem cell differentiation ($P=1.6x10^{-3}$) (Supplementary Table 40).

**Cell type enrichment analysis of negative control traits for glaucoma.** To rule out the possibility that some of the cell type enrichment signals are due to unaccounted confounding factors, we applied ECLIPSER to ocular and non-ocular diseases and traits unrelated to glaucoma (listed in Supplementary Table 45). The negative control traits showed enrichment in relevant cell types, such as pigmented cells for eye color, lens in the anterior segment for cataract, retinal pigment epithelium (RPE) in the macula and vascular cells in the ONH for AMD, immune cells for asthma and atopic dermatitis, and melanocytes for melanoma (Supplementary Table 46, Fig. 6a and Supplementary Fig. 21). Since macroglial and microglial cells have been suggested to be transcriptionally reactive to single cell dissociation protocols, more so than to single nuclei dissociation protocols[13–15], we evaluated the enrichment in these cell types among the negative control traits. No significant enrichment was found in retinal astrocytes or Müller Glia cells in any of the eight traits. AMD and atopic dermatitis showed nominal or FDR<0.1 enrichment, respectively, in astrocytes only in the macula single-nucleus dataset, but not in the retina dataset. Furthermore, we tested whether cell abundance may have an effect on cell type enrichment results. The ECLIPSER cell type enrichment significance (p-value) for the POAG cross-ancestry, POAG European, IOP, VCDR, machine learning-based VCDR, central corneal thickness, and corneal hysteresis GWAS did not correlate with cell count per cell type in any of the four single-nucleus datasets (Pearson $R^2<0.2$, P>0.12; Supplementary Table 47).

**Extended Discussion points:**

**Limitations to Bayesian colocalization methodologies.** There are several limitations to Bayesian-based colocalization analysis[16,17]. Under-estimation of enrichment of e/sQTLs in a given tissue in GWAS hits, used as a prior in the colocalization analysis, or GWAS loci with modest effect sizes may lead to loss of power[16]. Furthermore, the choice of genotype reference panel used to compute LD for fine-mapping of the GWAS and QTL loci may have an effect on colocalization sensitivity. We used GTEx as our reference panel, whose European to African sample ratio is comparable to that of the POAG cross-ancestry GWAS, but has substantially fewer East Asian samples[2]. We do not expect this to have a significant impact on our colocalization results as most of the POAG loci were found to be shared across populations[18], and the fraction of loci with significant colocalization results was similar between the POAG cross-ancestry and European subset GWAS meta-analyses. While colocalization analysis identifies GWAS and e/sQTL signals tagging the same causal variant/haplotype, which suggests that change in expression or splicing of the QTL target gene may be causal to the GWAS trait (vertical pleiotropy), we cannot rule out the possibility that the causal effect on POAG or IOP may be through another mechanism other than change in gene expression or alternative splicing (horizontal pleiotropy). We thus applied Mendelian randomization[19,20,21] to all significantly colocalizing e/sQTLs and GWAS loci to test for horizontal pleiotropy and provide additional support for a causal relationship between the colocalizing e/sQTLs and corresponding GWAS loci. Of note, some of the non-significant MR results for POAG may be due to a limitation of two sample MR that requires the e/sQTL and GWAS studies to be from similar population backgrounds, and hence could not be run on the larger and better-powered POAG cross-ancestry GWAS.

**Supplementary Methods**

**Enrichment of POAG and IOP associations among e/sQTLs using *QTLEnrich*.** To test whether genome-wide significant and nominal POAG and IOP trait associations are enriched among eQTLs and sQTLs, and to assess the contribution of e/sQTLs to these traits, we applied *QTLEnrich*[2] to the POAG and IOP GWAS meta-analyses summary statistics, and eQTLs and sQTLs from the 49 GTEx tissues[2,5] and eQTLs from peripheral retina (EyeGEx[5]). *QTLEnrich* is a rank and permutation-based method that evaluates the fold-enrichment significance of trait associations among a set of e/sQTLs in a given tissue, compared to a tissue-specific null distribution of variants matched on three confounding factors: minor allele frequency (MAF), distance to the target gene's transcription start site, and local LD[1]. Briefly, fold-enrichment was computed for each trait by e/sQTL-tissue combination as the fraction of e/sVariants (FDR<0.05) with a GWAS P-value<0.05 compared to expectation, assuming a uniform null distribution. Fold-enrichment was also computed for $<10^5$ randomly sampled sets of *k* null variants (non-significant e/sVariants; *k* = number of significant e/sVariants in given tissue) matched on the three confounding factors relative to each significant e/sVariant. Random sampling was applied to decile bins of the confounding factors with replacement. The null background of variants was defined as all common variants (MAF>1%) tested for e/sQTL in GTEx or retina excluding the significant e/sVariants (FDR<0.05). The enrichment p-value was computed as the fraction of up to $10^5$ null variant sets whose fold-enrichment was equal to or higher than that of the significant set of e/sVariants for a given tissue. To adjust for the potential enrichment of trait associations amongst null variants, an adjusted fold-enrichment was computed per trait-e/sQTL-tissue combination as the observed fold-enrichment divided by the median fold-enrichment of 1000 confounder-matched null variant sets. Local LD was estimated based on the number of LD proxy variants ($R^2$ > 0.5) per variant, using GTEx WGS v8 VCF as the reference panel. For the enrichment analysis only the most significant e/sVariant per e/sGene (FDR<0.05) in the given tissue was used to prevent inflation of trait association enrichment due to LD. In cases where the e/sQTL was the most significant e/sVariant for more than one e/sGene, the smallest distance to TSS was used. Only protein coding and lincRNA genes were considered in this analysis. Significant tissues were determined based on an Enrichment P-value that passed Bonferroni correction, correcting for 50 tissues and two QTL types tested ($P<5x10^{-4}$). The adjusted fold-enrichment was used to rank the significantly enriched tissues, as this statistic is not correlated with tissue sample size or number of significant e/sQTLs per tissue[1], in contrary to that observed with the colocalization analysis (Supplementary Fig. 4).

For the significant trait-tissue pairs, the fraction and number of e/sVariants proposed to be associated with POAG or IOP were estimated using an empirically derived, true positive rate (Adj. $\pi_1$) approach implemented in the latest version of *QTLEnrich* (URLs), based on Storey's analytical $\pi_1$[22,23] and an empirical FDR method[22,23]. The adj. $\pi_1$ adjusts for enrichment of trait associations amongst non-significant e/sQTL variants matched on confounding factors. A lambda of 0.8 was used to estimate the adjusted true negative rate. The estimated number of trait associations amongst a set of e/sVariants in a given tissue was computed as the adj. $\pi_1$ times the number of e/sQTLs analyzed for that tissue. In few cases, the adj. $\pi_1$ can be zero for e/sQTLs that display significant enrichment of trait associations. This may be caused by deflation of the GWAS p-values of the e/sQTL set at the higher end of p-values that are used to estimate the true negative rate. *QTLEnrich* thus also estimates a lower bound number of trait associations among e/sQTLs with GWAS P<0.05 that is not affected by this issue (Supplementary Tables 2-3).

**Colocalization analysis:**

**eCAVIAR analysis.** eCAVIAR[3] is a probabilistic model that estimates the posterior probability that the same variant is causal in overlapping GWAS and e/sQTL signals, accounting for the uncertainty of LD and allelic heterogeneity. It simultaneously performs statistical fine-mapping on the GWAS and e/sQTL signals to optimize integration. For the eCAVIAR analysis, we assumed at most two independent causal variants per locus[2]. Only variants present in both the GWAS and QTL studies were analyzed. LD matrices with $R^2$ values between all pairwise variant comparisons per locus were computed for each GWAS locus LD interval (defined above), using the GTEx whole genome sequencing (WGS) data from release v8 as the reference panel, and were used for fine-mapping of all variants in the GWAS and e/sQTL loci. The genotypes from all 838 GTEx donors in v8[2], consisting of European (84%), African (12.3%), and East Asian (1.4%) ancestries, were used for the LD calculations for the cross-ancestry POAG loci, and the European subset (715 donors) was used for the European POAG and IOP loci. Due to computational limitations, for GWAS LD intervals with more than 1000 variants, variants with e/sQTL p-values below 0.05 were filtered out. GWAS-e/sQTL-tissue combinations with a colocalization posterior probability (CLPP) above 0.01 were considered significant in this study, based on previous simulations that suggested a high true positive rate and low false positive rate at CLPP > 0.01[3]. Colocalization analysis of the retina eQTLs was only performed using eCAVIAR.

***enloc* analysis.** *enloc* (Enrichment estimation aided colocalization analysis)[4] is a Bayesian hierarchical model used to determine the probability of colocalization of molecular QTLs with GWAS signals, that consists of three steps: estimation of enrichment levels of QTLs among the GWAS hits, fine-mapping using an empirical Bayes method (DAP-G)[4], and colocalization analysis that estimates a regional colocalization probability (RCP) that is the sum of SNP-level colocalization probabilities (SCPs) of all variants per GWAS locus. *enloc* does not limit the potential number of independent causal variants. To estimate the posterior probability of causality (posterior inclusion probabilities (PIPs)) of variants in the GWAS loci we applied DAP-G (URLs) to all variants in the LD intervals defined above for the POAG cross-ancestry, POAG European, and IOP loci. For several loci, we restricted the maximum model size parameter (-msize) to 5, which represents the number of independent association clusters, due to memory limitations. DAP-G outputs were post-processed, selecting all fine-mapped variants and their associated cluster number and PIP for each GWAS locus which were inputted into *enloc*. For the GTEx e/sQTLs, we used similar DAP-G fine-mapping results published for all e/sQTLs in 49 GTEx tissues that were computed using the European subset of GTEx samples[24]. LD matrices were computed per GWAS locus using all GTEx WGS samples for the POAG cross-ancestry loci and the subset of European individuals in GTEx for the POAG EUR and IOP loci, as the reference panel. We used an LD-threshold of 0.75 (-ld_control) that determines the strength of genotype correlation between variants that belong to a given fine-mapped signal cluster, as used in the GTEx European subset e/sQTL analysis[24]. Finally, we applied *fastEnloc*[25] (URLs), a faster implementation of *enloc*, to the fine-mapped results of the GWAS loci and overlapping GTEx e/sQTLs. Only e/sVariants included in a 25% or greater credible set of causal variants and that overlapped with the GWAS variants were analyzed in the colocalization analysis. The colocalization analysis outputted RCP values above $1x10^{-4}$ for each tested GWAS locus, trait, tissue, and gene or gene-intron excision cluster combination. An RCP above 0.1 was considered significant in this study, based on the method's recommendation[4,16]. In Supplementary Tables 10-12 we listed the set of variants included in the 95% credible set for each independent GWAS locus, e/sQTL, gene and tissue colocalization signal (cluster). In cases where a significantly colocalizing sQTL that regulates an intron cluster mapped to more than one gene, all target genes were reported as significant.

**Quality control and summary across the two colocalization methods.** In inspecting the significant colocalization results and plotting the $-\log_{10}$(P-value) of the GWAS signal versus the $-\log_{10}$(P-value) of the e/sQTL signal, we noticed multiple cases where the GWAS P-values and/or the e/sQTL P-values of the variants with significant colocalization posterior probabilities (CLPP>0.01 and/or RCP>0.1) were not significant (e.g., association P>0.05) or did not pass multiple hypothesis correction (see examples in Supplementary Fig. 3). Thus, to remove potential false positives, we filtered out variant, gene, tissue, trait combinations where the e/sVariant with a significant colocalization result had a GWAS p-value above $1x10^{-5}$ or whose e/sQTL p-values was above $1x10^{-4}$ and/or did not pass FDR<0.05 (FALSE in column 'Pass_QC_QTL_FDR05_P1E04_GWAS_P1E05' in Supplementary Tables 7-12). We chose a GWAS $P<10^{-5}$ cutoff, as it has been suggested that in many cases the lead GWAS variant is not the causal variant, but instead tagging the causal variant that may have not reached genome-wide significance[26]. We defined two sets of putative causal genes and regulatory mechanisms for POAG and IOP: a "comprehensive set" based on significant colocalization with at least one of the colocalization methods (CLPP > 0.01 and/or RCP > 0.1) (Supplementary Table 13), and a "high confidence set" based on significance with both methods (CLPP > 0.01 and RCP > 0.1; Table 1 and Supplementary Table 15). Since the number of significantly colocalizing e/sGenes correlated with the tissue sample size (coefficient of determination $R^2$=0.72, P=1x$10^{-14}$, Supplementary Fig. 4), we disregarded the tissue and only considered the QTL type and e/sGene when generating the list of prioritized causal genes per GWAS locus.

**Mendelian randomization (MR) of significantly colocalizing genes.** We used Mendelian randomization (MR)[21] to identify a high confidence set of e/sQTLs that may be causal to POAG and/or IOP of the significant e/sQTL-GWAS colocalization results. MR comprises the use of genetic variants within an instrumental variable (IV) framework to facilitate causal inference[27]. For the variants to be valid, three assumptions must be met: (1) Relevance: the variants associate robustly with the exposure, (2) Independence: the variants are independent of confounders of the exposure and outcome, and (3) Exclusion-restriction: the genetic variants influence the outcome only via the exposure, i.e., there is no horizontal pleiotropy. To instrument putative causal genes, for each e/sQTL, we selected a set of LD-independent variants (pairwise LD threshold of $r^2 < 0.1$) that associated with gene expression at $P < 5 \times 10^{-6}$. If there were no significant e/sVariants at $P < 5 \times 10^{-6}$ for a given e/sGene and tissue, FDR < 0.05 was used. LD pruning was performed using 1000 Genomes Project Phase 3 version 5[28] as the reference panel and PLINK v2.0[29]. Two-sample MR was applied to the summary statistics of the e/sQTLs (exposure) and POAG or IOP GWAS (outcome) for all colocalizing e/sQTL-GWAS locus pairs. This included 1,866 eQTLs targeting 408 genes and 1,068 sQTLs targeting 147 genes in a range of tissues that colocalized with POAG and/or IOP GWAS loci based on eCAVIAR and/or *enloc* (Supplementary Tables 7-13). Effect alleles were aligned between the exposure and outcome datasets to harmonize genetic associations between the two studies. The Wald ratio, i.e., the variant-outcome association beta divided by the variant-exposure association beta[30], was used for exposures where only one variant constituted the genetic instrument. Where multiple variants constituted the instrument for the targeted e/sGene, the inverse-variance weighted (IVW) method was used as the primary method for pooling variant-specific estimates[31]. We applied four additional methods to test the robustness of the results, given that the IVW approach assumes no horizontal pleiotropy: the simple-median[32], weighted-median[32], MR-Egger[33], and MR-PRESSO[34] methods. Horizontal pleiotropy was tested using the Egger-intercept test and the MR-PRESSO global heterogeneity test; $P < 0.05$ indicated the presence of horizontal pleiotropy. If horizontal pleiotropy was found only based on the MR-PRESSO global heterogeneity test, an MR PRESSO outlier-corrected p-value < 0.05 was considered a significant result. Due to potential differences in allele frequency between populations, MR was applied only to the European ancestry subset of the POAG GWAS meta-analysis and the IOP GWAS meta-analysis that is primarily comprised of European individuals, to avoid confounding by ancestry. Due to this limitation, some of the non-significant MR results for POAG may be due to not having applied MR on the larger, better-powered POAG cross-ancestry GWAS, where in many cases the colocalization was observed. All MR results are summarized in Supplementary Table 29.

**Single nucleus RNA-seq datasets and differential gene expression.**

**Retina**: Retina samples from the fovea, macula and/or periphery were collected from six donors within 6 hours from death from the Utah Lions Eye Bank (Supplementary Table 35), flash-frozen, and processed, as described in Liang *et al*.[9]. The number of cells from the three retinal regions from each donor is given in Supplementary Table 35. The fovea (with central macula) and macula samples were collected separately, using a 4 mm and 6 mm disposable biopsy punch, respectively, and were flash-frozen in liquid nitrogen. snRNA-seq data from RGCs from a few additional donors were added to the data set, given the relevance of RGCs to glaucoma, though RGCs still only comprised about 1/250 of the total data set. Nuclei were isolated by pre-chilled fresh-made lysis buffer (10 mM Tris-HCl, 10 mM NaCl, 3 mM MgCl2, 0.02% NP40). The frozen tissue was triturated to break the tissue structure in lysis buffer and homogenized with a Wheaton™ Dounce Tissue Grinder. For the nuclei to be enriched, isolated nuclei were stained with mouse anti-NeuN monoclonal antibody in pre-chilled wash buffer (1% BSA in PBS, 0.2U/μl RNAse inhibitor) for 30min at 4°C. After being centrifuged at 500g, the pellet was resuspended in wash buffer and filtered with 40μm Flowmi Cell Strainer. DAPI (4′,6-diamidino-2-phenylindole, 10 μg/ml) was added before loading the nuclei for fluorescent cytometry sorting. Stained nuclei were sorted with a FACS Aria II flow sorter (Becton Dickinson, San Jose, CA) (70μm nozzle). Sorting gates were based on the strengths of DAPI signal and FITC signal. Samples were sorted at a rate of 50 events per second. Fluorescence detection used a 450-nm/40-nm-band pass barrier filter for DAPI, and a 530-nm/30-nm-band pass filter for FITC. All single-nuclei RNA sequencing of the retina samples was performed at the Single Cell Genomics Core at Baylor College of Medicine. Single-nuclei cDNA library preparation and sequencing were performed following the manufacturer's protocols (https://www.10xgenomics.com). Single-nuclei suspension was loaded on a Chromium controller to obtain single-cell GEMS (Gel Beads-In-Emulsions) for the reaction. The snRNA-seq library was prepared with Chromium Next GEM Single Cell 3' Kit v3.1 (10x Genomics). The library was then sequenced on Illumina NovaSeq 6000 (https://www.illumina.com). Reads from snRNA-seq were demultiplexed and then aligned to the 'Ref_cellranger.hg19.premrna' human reference (from 10x Genomics) using Cell Ranger (version 3.0.2, 10x Genomics). The gene expression matrices were generated by the Cell Ranger pipeline.

For each matrix, genes detected in less than 5 cells were removed; cells with total UMI counts less than 500 were removed, and the top 5% of cells with the highest mitochondrial gene expression proportion were removed. We then performed doublet removal on the remaining cells using DoubletFinder in Seurat[35]. The gene expression was then normalized by total UMI counts for each cell, scaled by 10000, and transformed with the natural logarithm. To account for batch effects originating from sample collection processes, the CarDEC framework[36] was applied with standard parameters. Both highly variable genes (top 3000) and lowly variable genes were used for building the model, and donor resources were used as the batch key. The low dimensional embeddings generated were then used to perform leiden clustering, and major cell types were annotated based on known marker genes (GNAT1, PDE6A for rod; ARR3, PDE6C for cone; RLBP1, GLUL for MG; C1QA, C1QB for microglia; GFAP for astrocyte; RPE65, LRAT for RPE). Differential gene expression for each cell type in the ml_class level used in this study was computed using the Wilcoxon rank sum test, applied to genes expressed in at least 5% of cells per cell type.

**Optic nerve head and posterior tissues:** The optic nerve head, including peripapillary tissues, was dissected with 4 mm punches from six donors within 4 hours of death at the University of Utah and Massachusetts General Hospital (Supplementary Table 37), as described in Monavarfeshani *et al.*[10] Tissues were frozen and kept at -80C. For single-nuclei isolation, frozen tissues were homogenized in a dounce homogenizer with 0.1% NP-40 lysis buffer and passed through a 40-μm cell strainer. The nuclei were then pelleted at 500 rcf over 5 min, resuspended in 2% BSA, and stained with DAPI. ~40K DAPI+ nuclei were sorted and pelleted at 500 rcf over 5 min. Finally, nuclei were resuspended in 0.04% non-acetylated BSA/PBS solution and adjusted to a concentration of 1000 nuclei/μL. The integrity of the nuclear membrane, protrusion of nuclear content, and presence of non-nuclear material were assessed under a brightfield microscope. Nuclei were then loaded into a 10X Chromium Single Cell Chip with a targeted recovery of 8000 nuclei. Single nuclei libraries were generated with Chromium 3’ V3.1 platform (10X Genomics, Pleasanton, CA) following the manufacturer’s protocol. Briefly, single nuclei were partitioned into Gel-beads-in-EMulsion (GEMs) where nuclear lysis and barcoded reverse transcription of RNA would take place to yield full-length cDNA; this was followed by amplification, enzymatic fragmentation, and 5’ adaptor and sample index attachment to yield the final libraries. The single nucleus RNA sequencing of the optic nerve head samples was performed on NovaSeq machines at a Harvard University facility. snRNA data processing and analyses were performed similarly to the pipeline used for the anterior segment in van Zyl *et al.*[8].

### URLs

GTEx: https://gtexportal.org/home/datasets
EyeGEx: https://gtexportal.org/home/datasets
QTLEnrich: https://github.com/segrelabgenomics/QTLEnrich
GeneEnrich: https://github.com/segrelabgenomics/GeneEnrich
MSigDB: http://www.gsea-msigdb.org/gsea/msigdb/collections.jsp
PLINK: https://www.cog-genomics.org/plink/
eCAVIAR: https://github.com/fhormoz/caviar
fastEnloc: https://github.com/xqwen/fastenloc
DAP-G: https://github.com/xqwen/dap/tree/master/dap_src
COJO: https://yanglab.westlake.edu.cn/software/gcta/
ECLIPSER: https://github.com/segrelabgenomics/ECLIPSER
genomAD: https://gnomad.broadinstitute.org/
QMplot: https://github.com/ShujiaHuang/qmplot
LocusCompare: https://github.com/boxiangliu/locuscomparer

### References


1. Gamazon, E. R. *et al.* Using an atlas of gene regulation across 44 human tissues to inform complex disease- and trait-associated variation. *Nat. Genet.* **50**, 956–967 (2018).
2. GTEx Consortium. The GTEx Consortium atlas of genetic regulatory effects across human tissues. *Science* **369**, 1318–1330 (2020).
3. Hormozdiari, F. *et al.* Colocalization of GWAS and eQTL Signals Detects Target Genes. *Am. J. Hum. Genet.* **99**, 1245–1260 (2016).
4. Wen, X., Pique-Regi, R. & Luca, F. Integrating molecular QTL data into genome-wide genetic association analysis: Probabilistic assessment of enrichment and colocalization. *PLoS Genet.* **13**, e1006646 (2017).
5. Ratnapriya, R. *et al.* Retinal transcriptome and eQTL analyses identify genes associated with age-related macular degeneration. *Nat. Genet.* **51**, 606–610 (2019).
6. Rouhana, J., J. Wang, G. Eraslan, S. Anand, A. Hamel, B. Cole, A. Regev, F. Aguet, K. Ardlie, and A. V. Segrè. ECLIPSER: identifying causal cell types and genes for complex traits through single cell enrichment of e/sQTL-mapped genes in GWAS loci. *BioRxiv* (2021) doi:10.1101/2021.11.24.469720.
7. Eraslan, G. *et al.* Single-nucleus cross-tissue molecular reference maps to decipher disease gene function. *bioRxiv* 2021.07.19.452954 (2021) doi:10.1101/2021.07.19.452954.

8. van Zyl, T. *et al.* Cell atlas of the human ocular anterior segment: Tissue-specific and shared cell types. *Proc. Natl. Acad. Sci. U. S. A.* **119**, e2200914119 (2022).
9. Liang, Q. *et al.* A multi-omics atlas of the human retina at single-cell resolution. *Cell Genom.* **3**, 100298 (2023).
10. Monavarfeshani, A. *et al.* Transcriptomic Analysis of the Ocular Posterior Segment Completes a Cell Atlas of the Human Eye. *bioRxiv* (2023) doi:10.1101/2023.04.26.538447.
11. Finucane, H. K. *et al.* Heritability enrichment of specifically expressed genes identifies disease-relevant tissues and cell types. *Nat. Genet.* **50**, 621–629 (2018).
12. Watanabe, K., Umićević Mirkov, M., de Leeuw, C. A., van den Heuvel, M. P. & Posthuma, D. Genetic mapping of cell type specificity for complex traits. *Nat. Commun.* **10**, 3222 (2019).
13. Todd, L. *et al.* Reactive microglia and IL1β/IL-1R1-signaling mediate neuroprotection in excitotoxin-damaged mouse retina. *J. Neuroinflammation* **16**, 118 (2019).
14. Denisenko, E. *et al.* Systematic assessment of tissue dissociation and storage biases in single-cell and single-nucleus RNA-seq workflows. *Genome Biol.* **21**, 130 (2020).
15. Eraslan, G. *et al.* Single-nucleus cross-tissue molecular reference maps toward understanding disease gene function. *Science* **376**, eabl4290 (2022).
16. Hukku, A. *et al.* Probabilistic colocalization of genetic variants from complex and molecular traits: promise and limitations. *Am. J. Hum. Genet.* **108**, 25–35 (2021).
17. Wallace, C. Eliciting priors and relaxing the single causal variant assumption in colocalisation analyses. *PLoS Genet.* **16**, e1008720 (2020).
18. Gharahkhani, P. *et al.* Genome-wide meta-analysis identifies 127 open-angle glaucoma loci with consistent effect across ancestries. *Nat. Commun.* **12**, 1258 (2021).
19. Zhu, Z. *et al.* Integration of summary data from GWAS and eQTL studies predicts complex trait gene targets. *Nat. Genet.* **48**, 481–487 (2016).
20. Hemani, G., Bowden, J. & Davey Smith, G. Evaluating the potential role of pleiotropy in Mendelian randomization studies. *Hum. Mol. Genet.* **27**, R195–R208 (2018).
21. Zuber, V. *et al.* Combining evidence from Mendelian randomization and colocalization: Review and comparison of approaches. *Am. J. Hum. Genet.* **109**, 767–782 (2022).
22. Storey, J. D. & Tibshirani, R. Statistical significance for genomewide studies. *Proc. Natl. Acad. Sci. U. S. A.* **100**, 9440–9445 (2003).
23. Gamazon, E. R., Huang, R. S., Dolan, M. E., Cox, N. J. & Im, H. K. Integrative genomics: quantifying significance of phenotype-genotype relationships from multiple sources of high-throughput data. *Front. Genet.* **3**, 202 (2012).

24. Barbeira, A. N. *et al.* Exploiting the GTEx resources to decipher the mechanisms at GWAS loci. *Genome Biol.* **22**, 49 (2021).
25. Pividori, M. *et al.* PhenomeXcan: Mapping the genome to the phenome through the transcriptome. *Sci. Adv.* **6**, eaba2083 (2020).
26. Brown, A. A. *et al.* Predicting causal variants affecting expression by using whole-genome sequencing and RNA-seq from multiple human tissues. *Nat. Genet.* **49**, 1747–1751 (2017).
27. Smith, G. D. & Ebrahim, S. "Mendelian randomization": can genetic epidemiology contribute to understanding environmental determinants of disease? *Int. J. Epidemiol.* **32**, 1–22 (2003).
28. 1000 Genomes Project Consortium *et al.* A global reference for human genetic variation. *Nature* **526**, 68–74 (2015).
29. Purcell, S. *et al.* PLINK: a tool set for whole-genome association and population-based linkage analyses. *Am. J. Hum. Genet.* **81**, 559–575 (2007).
30. Burgess, S., Small, D. S. & Thompson, S. G. A review of instrumental variable estimators for Mendelian randomization. *Stat. Methods Med. Res.* **26**, 2333–2355 (2017).
31. Burgess, S., Butterworth, A. & Thompson, S. G. Mendelian randomization analysis with multiple genetic variants using summarized data. *Genet. Epidemiol.* **37**, 658–665 (2013).
32. Bowden, J., Davey Smith, G., Haycock, P. C. & Burgess, S. Consistent Estimation in Mendelian Randomization with Some Invalid Instruments Using a Weighted Median Estimator. *Genet. Epidemiol.* **40**, 304–314 (2016).
33. Bowden, J., Davey Smith, G. & Burgess, S. Mendelian randomization with invalid instruments: effect estimation and bias detection through Egger regression. *Int. J. Epidemiol.* **44**, 512–525 (2015).
34. Verbanck, M., Chen, C.-Y., Neale, B. & Do, R. Detection of widespread horizontal pleiotropy in causal relationships inferred from Mendelian randomization between complex traits and diseases. *Nat. Genet.* **50**, 693–698 (2018).
35. Hao, Y. *et al.* Dictionary learning for integrative, multimodal and scalable single-cell analysis. *Nat. Biotechnol.* (2023) doi:10.1038/s41587-023-01767-y.
36. Lakkis, J. *et al.* A joint deep learning model enables simultaneous batch effect correction, denoising, and clustering in single-cell transcriptomics. *Genome Res.* **31**, 1753–1766 (2021).
