## Supplementary Figures for "Integrating genetic regulation and single-cell expression with GWAS prioritizes causal genes and cell types for glaucoma"

**Affiliations:**
[1]Ocular Genomics Institute, Department of Ophthalmology, Massachusetts Eye and Ear, Boston, MA
[2]Department of Ophthalmology, Harvard Medical School, Boston, MA
[3]Broad Institute of Harvard and MIT, Cambridge, MA.
[4]Department of Molecular and Cellular Biology and Center for Brain Science, Harvard University, Cambridge, MA, USA
[5]MRC Human Genetics Unit, Institute of Genetics and Cancer, The University of Edinburgh, Edinburgh, UK
[6]Centre for Genomic and Experimental Medicine, Institute of Genetics and Molecular Medicine, The University of Edinburgh, Edinburgh, UK
[7]Neurobiology, Neurodegeneration and Repair Laboratory, National Eye Institute, National Institutes of Health, Bethesda, Maryland, USA
[8]Department of Molecular and Human Genetics, Baylor College of Medicine, TX
[9]Centre for Evidence-Based Medicine, University of Oxford, Oxford, UK
[10]Faculty of Medicine, Imperial College London, London, UK
[11]Harvard/MIT MD-PhD Program, Harvard Medical School, Boston, MA
[12]Department of Ophthalmology and Visual Sciences, Yale School of Medicine, New Haven, CT
[13]Department of Ophthalmology, Icahn School of Medicine at Mount Sinai, New York, NY, USA;
[14]QIMR Berghofer Medical Research Institute, Brisbane, Queensland, 4029, Australia
[15]NIHR Biomedical Research Centre, Moorfields Eye Hospital NHS Foundation Trust and UCL Institute of Ophthalmology, London, UK

* Co-second authors, these authors contributed equally

### Supplementary Figure 1

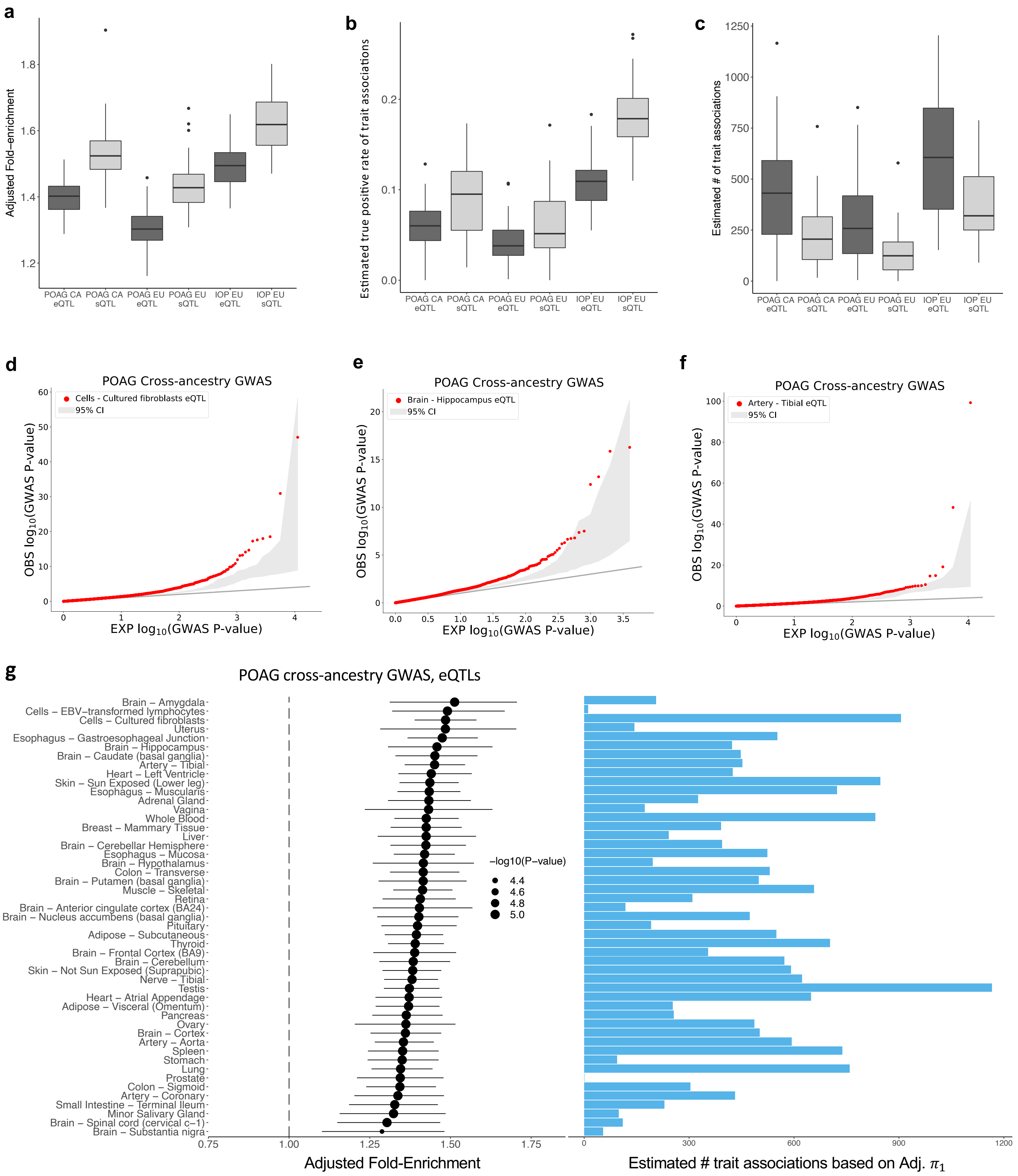

**Supplementary Figure 1. e/sQTLs enriched for hundreds of new POAG and IOP associations. a-c**, Boxplots showing the distributions of three *QTLEnrich* summary statistics for POAG cross-ancestry GWAS, POAG European (EUR) ancestry GWAS, and IOP GWAS for the significant tissue-trait pairs that passed Bonferroni correction for testing enrichment of trait associations amongst eQTLs (dark grey) and sQTLs (light grey) from 49 GTEx tissues and retina eQTLs: adjusted fold-enrichment (**a**), estimated true positive rate of trait associations among e/sQTLs (Adjusted $\boldsymbol{\pi}_1$) (**b**), and estimated number of trait associations based on adjusted $\boldsymbol{\pi}_1$ **(c)**. The center lines in the box plots depict the median value and the box edges the interquartile range. **d-f** Q-Q plots of $-\log_{10}$(P-value) of POAG cross-ancestry GWAS for the significant best eQTL per eGene set (FDR < 0.05) in top enriched tissues (red points): Cells - Cultured fibroblasts **(d),** Brain - Hippocampus **(e),** and Artery Tibial **(f)** compared to GWAS p-values of 1,000 confounder-matched null variant sets (light grey) generated by *QTLEnrich*. **g**, Forest plots (left panel) and barplots (right panel) of all tissues whose eQTLs were significantly enriched (Bonferroni correction) for POAG cross-ancestry GWAS associations based on *QTLEnrich*. Points indicate the adjusted fold-enrichment scaled by -log10(Enrichment P-value) and the lines represent 95% confidence intervals. Barplots show estimated number of e/sQTLs that are likely true trait associations per enriched tissue using an adjusted true positive rate, $\boldsymbol{\pi}_1$ approach. The lack of estimated number of POAG associations among prostate eQTLs is likely due to deflation of the GWAS p-values of the prostate eQTL set (Methods).

**Supplementary Figure 2. POAG and IOP trait associations amongst GTEx tissue and retina e/sQTLs**

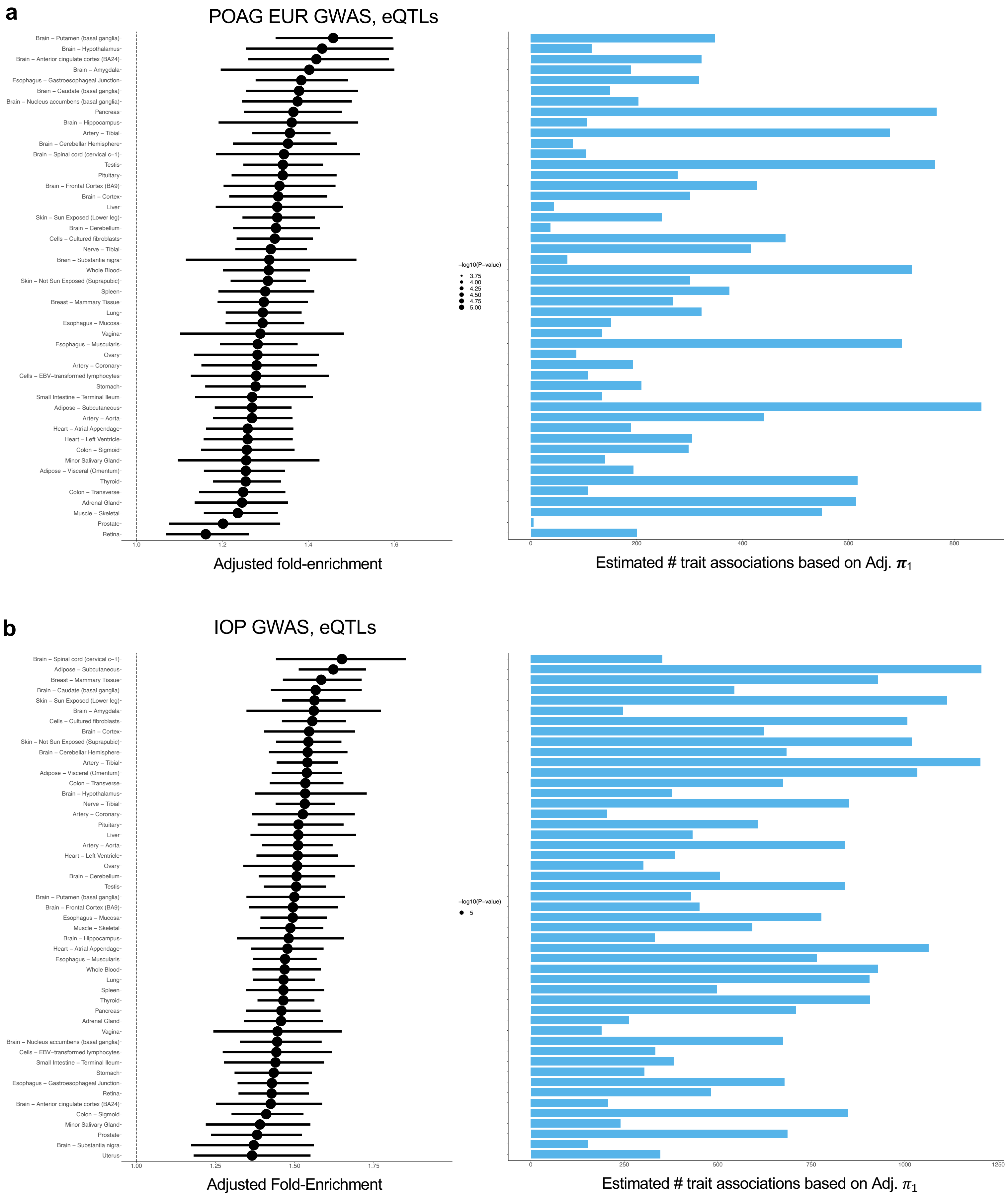

#### Supplementary Figure 2. POAG and IOP trait associations amongst GTEx tissue and retina e/sQTLs

**c**

POAG cross-ancestry GWAS, sQTLs

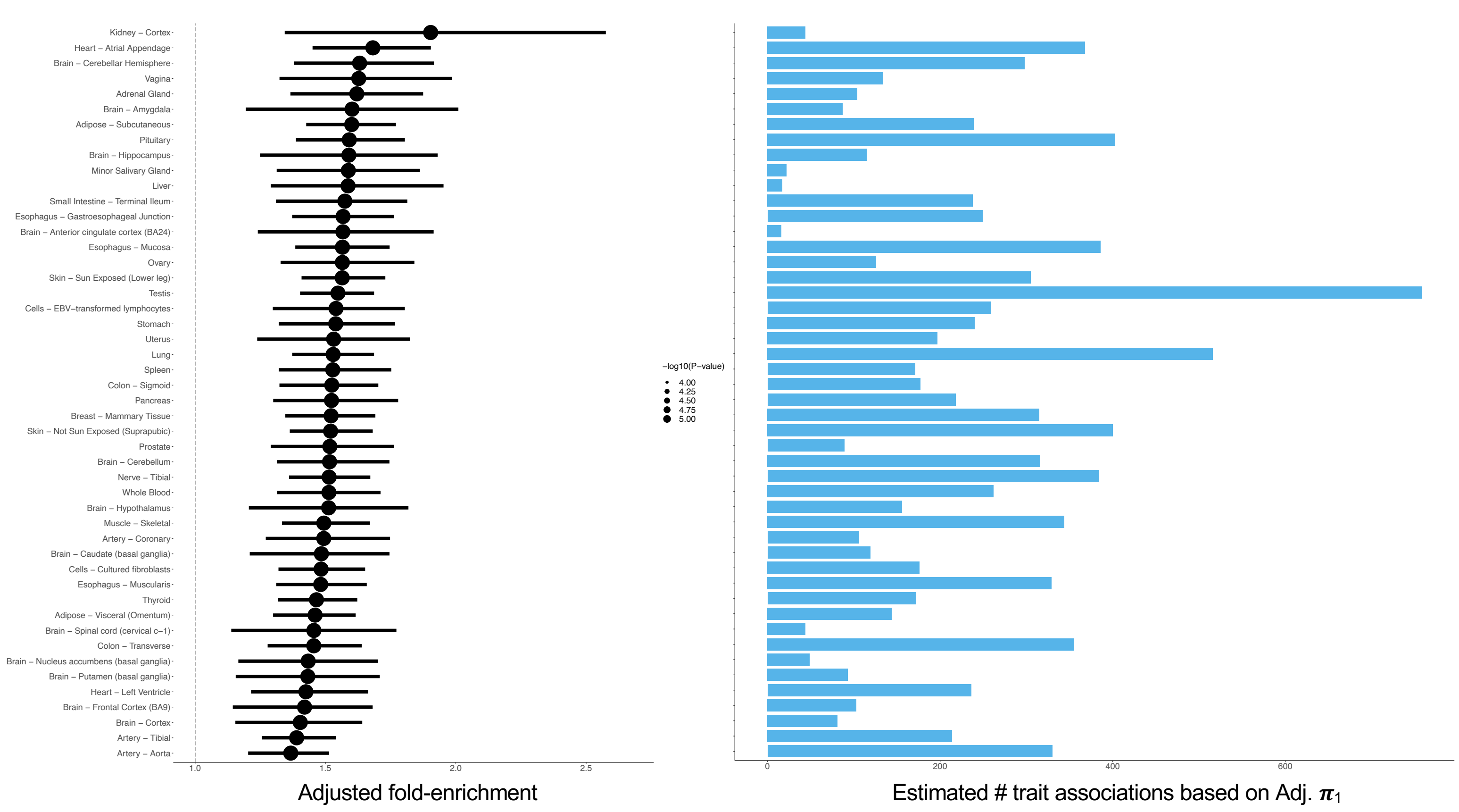


**d**

POAG EUR GWAS, sQTLs

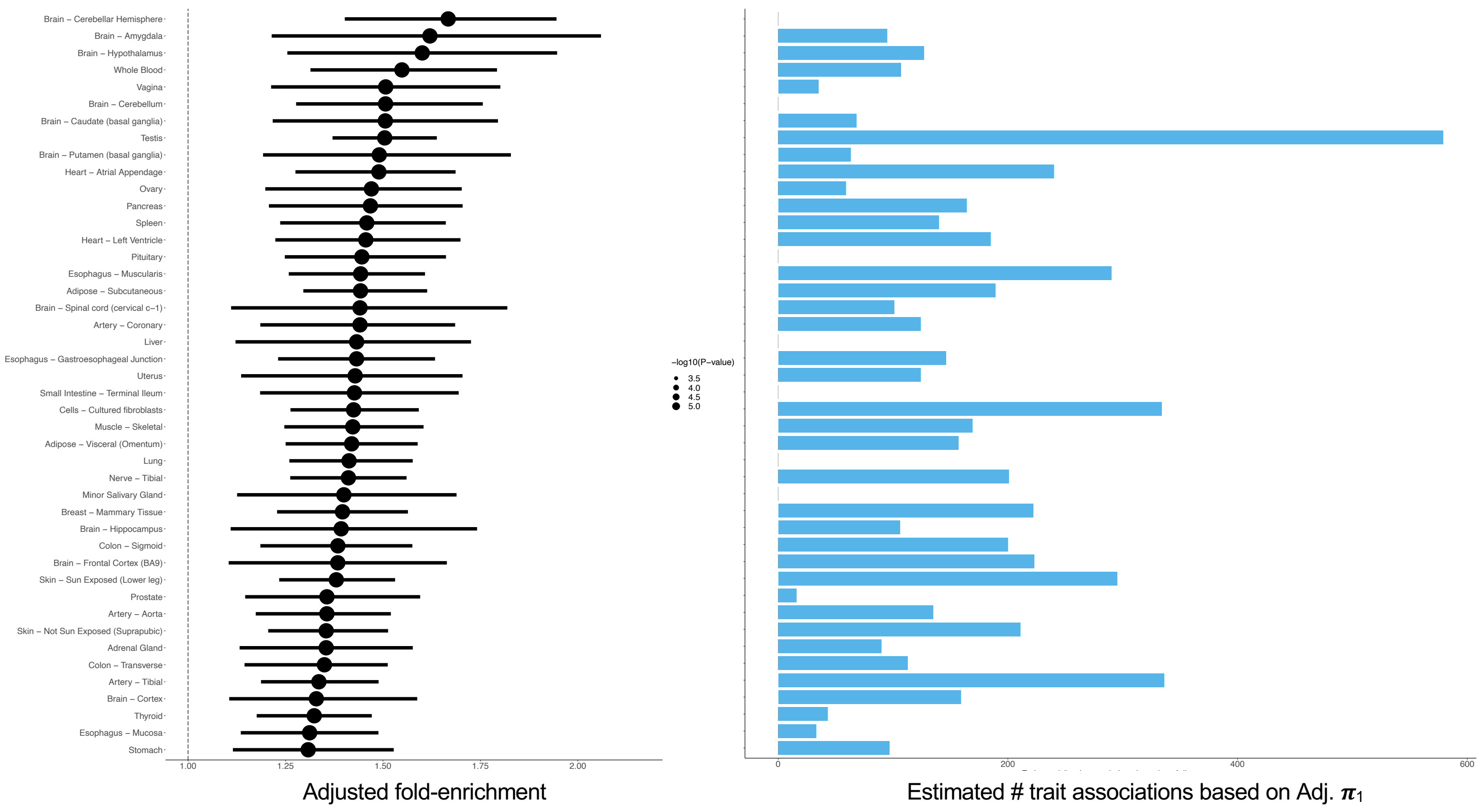

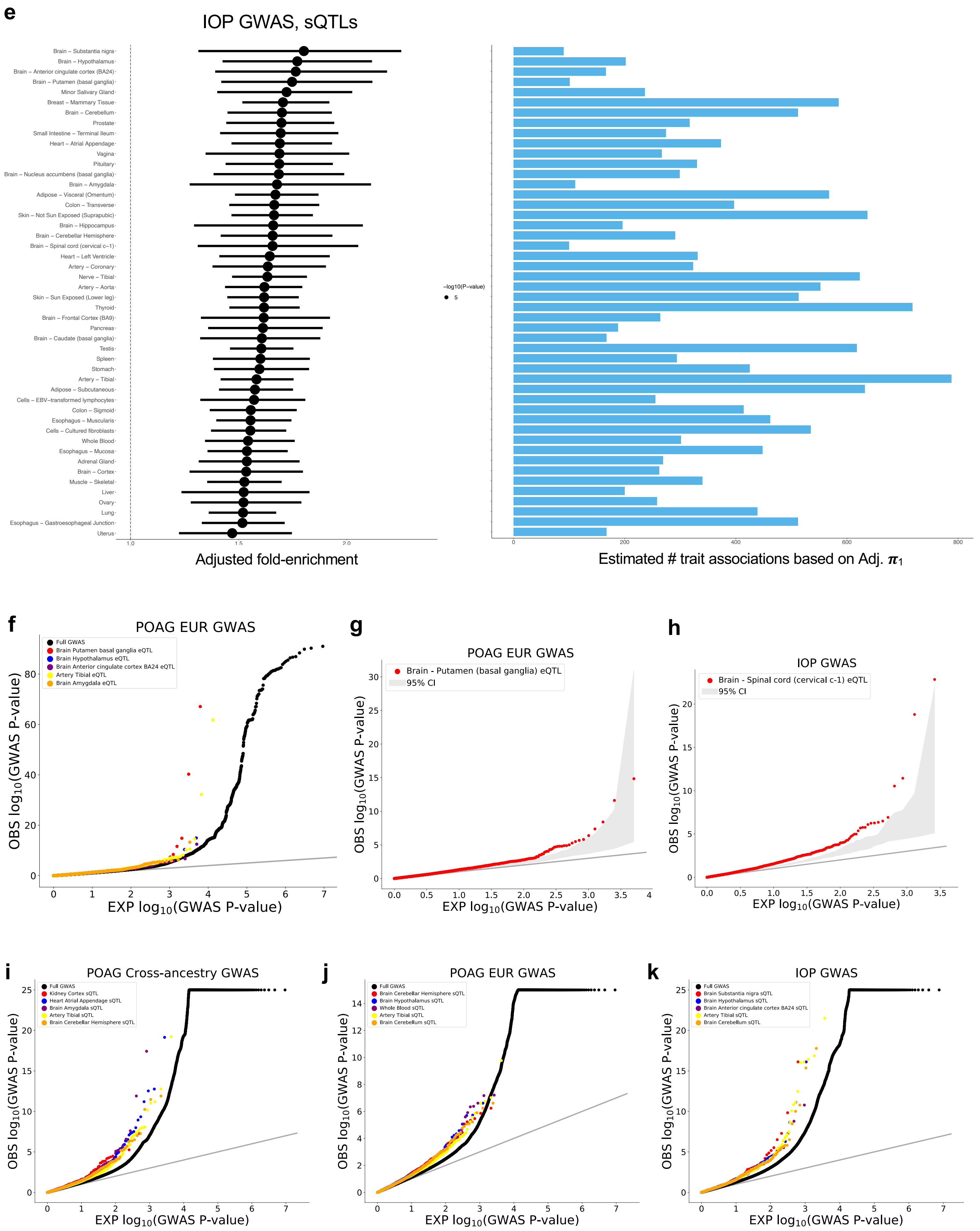
e
IOP GWAS, sQTLs
Brain – Substantia nigra
Brain – Hypothalamus
Brain – Anterior cingulate cortex (BA24)
Brain – Putamen (basal ganglia)
Minor Salivary Gland
Breast – Mammary Tissue
Brain – Cerebellum
Prostate
Small Intestine – Terminal Ileum
Heart – Atrial Appendage
Vagina
Pituitary
Brain – Nucleus accumbens (basal ganglia)
Brain – Amygdala
Adipose – Visceral (Omentum)
Colon – Transverse
Skin – Not Sun Exposed (Suprapubic)
Brain – Hippocampus
Brain – Cerebellar Hemisphere
Brain – Spinal cord (cervical c-1)
Heart – Left Ventricle
Artery – Coronary
Nerve – Tibial
Artery – Aorta
Skin – Sun Exposed (Lower leg)
Thyroid
Brain – Frontal Cortex (BA9)
Pancreas
Brain – Caudate (basal ganglia)
Testis
Spleen
Stomach
Artery – Tibial
Adipose – Subcutaneous
Cells – EBV-transformed lymphocytes
Colon – Sigmoid
Esophagus – Muscularis
Cells – Cultured fibroblasts
Whole Blood
Esophagus – Mucosa
Adrenal Gland
Brain – Cortex
Muscle – Skeletal
Liver
Ovary
Lung
Esophagus – Gastroesophageal Junction
Uterus
1.0
1.5
2.0
Adjusted fold-enrichment
−log10(P−value)
5
0
200
400
600
800
Estimated # trait associations based on Adj. $\pi_1$
f
POAG EUR GWAS
Full GWAS
Brain Putamen basal ganglia eQTL
Brain Hypothalamus eQTL
Brain Anterior cingulate cortex BA24 eQTL
Artery Tibial eQTL
Brain Amygdala eQTL
OBS $\log_{10}$(GWAS P-value)
EXP $\log_{10}$(GWAS P-value)
g
POAG EUR GWAS
Brain - Putamen (basal ganglia) eQTL
95% CI
OBS $\log_{10}$(GWAS P-value)
EXP $\log_{10}$(GWAS P-value)
h
IOP GWAS
Brain - Spinal cord (cervical c-1) eQTL
95% CI
OBS $\log_{10}$(GWAS P-value)
EXP $\log_{10}$(GWAS P-value)
i
POAG Cross-ancestry GWAS
Full GWAS
Kidney Cortex sQTL
Heart Atrial Appendage sQTL
Brain Amygdala sQTL
Artery Tibial sQTL
Brain Cerebellar Hemisphere sQTL
OBS $\log_{10}$(GWAS P-value)
EXP $\log_{10}$(GWAS P-value)
j
POAG EUR GWAS
Full GWAS
Brain Cerebellar Hemisphere sQTL
Brain Hypothalamus sQTL
Whole Blood sQTL
Artery Tibial sQTL
Brain Cerebellum sQTL
OBS $\log_{10}$(GWAS P-value)
EXP $\log_{10}$(GWAS P-value)
k
IOP GWAS
Full GWAS
Brain Substantia nigra sQTL
Brain Hypothalamus sQTL
Brain Anterior cingulate cortex BA24 sQTL
Artery Tibial sQTL
Brain Cerebellum sQTL
OBS $\log_{10}$(GWAS P-value)
EXP $\log_{10}$(GWAS P-value)

**Supplementary Figure 2. POAG and IOP trait associations amongst GTEx tissues and retina e/sQTLs. a-e**, Forest plots (left panel) and barplots (right panel) of all tissues whose eQTLs were significantly enriched (Bonferroni correction) for POAG European (**a**) or IOP (**b**) GWAS associations, or whose sQTLs were enriched for POAG cross-ancestry (**c**), POAG European (**d**), or IOP (**e**) associations based on *QTLEnrich*. Points indicate the adjusted fold-enrichment scaled by -log10(Enrichment P-value) and the lines represent 95% confidence intervals. Barplots show estimated number of e/sQTLs that are likely true trait associations per enriched tissue using an adjusted true positive rate, $\pi_1$ approach. For several tissues with significant enrichment in (**d)**, the estimated number of POAG EUR associations among sQTLs was zero (e.g., brain cerebellum or pituitary). This is likely due to deflation of the GWAS p-values of the sQTL set at the higher end of p-values, that are used to estimate the true negative rate (see Methods). When considering only the top ranked sQTLs with GWAS P<0.05 in these tissue, a lower bound of 25-88 of the sQTLs were estimated to be true POAG associations (Supplementary Table 3). **f,** Quantile-quantile (Q-Q) plot of POAG European GWAS -$\log_{10}$(P-value) compared to expectation, assuming a null uniform distribution, for the eQTLs (best eQTL per eGene set with FDR < 0.05) in the most significantly enriched tissues based on adjusted fold-enrichment (colored points), compared to all variants in the GWAS (black points). Grey line represents the diagonal. **g-h,** Q-Q plot of POAG European (**g**) or IOP (**h**) GWAS -$\log_{10}$(P-value) of the best eQTL per eGene set in the top ranked tissue per trait based on adjusted fold-enrichment (red points), compared to GWAS p-values of 1,000 confounder-matched null variant sets (light grey) generated by *QTLEnrich*. **i-k**, Q-Q plot of POAG Cross-ancestry (**i**), POAG European (**j**), and IOP (**k**) GWAS -$\log_{10}$(P-value) compared to expectation for the sQTLs (best sQTL per sGene at FDR<0.05) in the top enriched tissues per trait based on adjusted fold-enrichment compared to all variants tested in the GWAS (black).

**Supplementary Figure 3. Examples of potential false positive colocalization results that were filtered out**

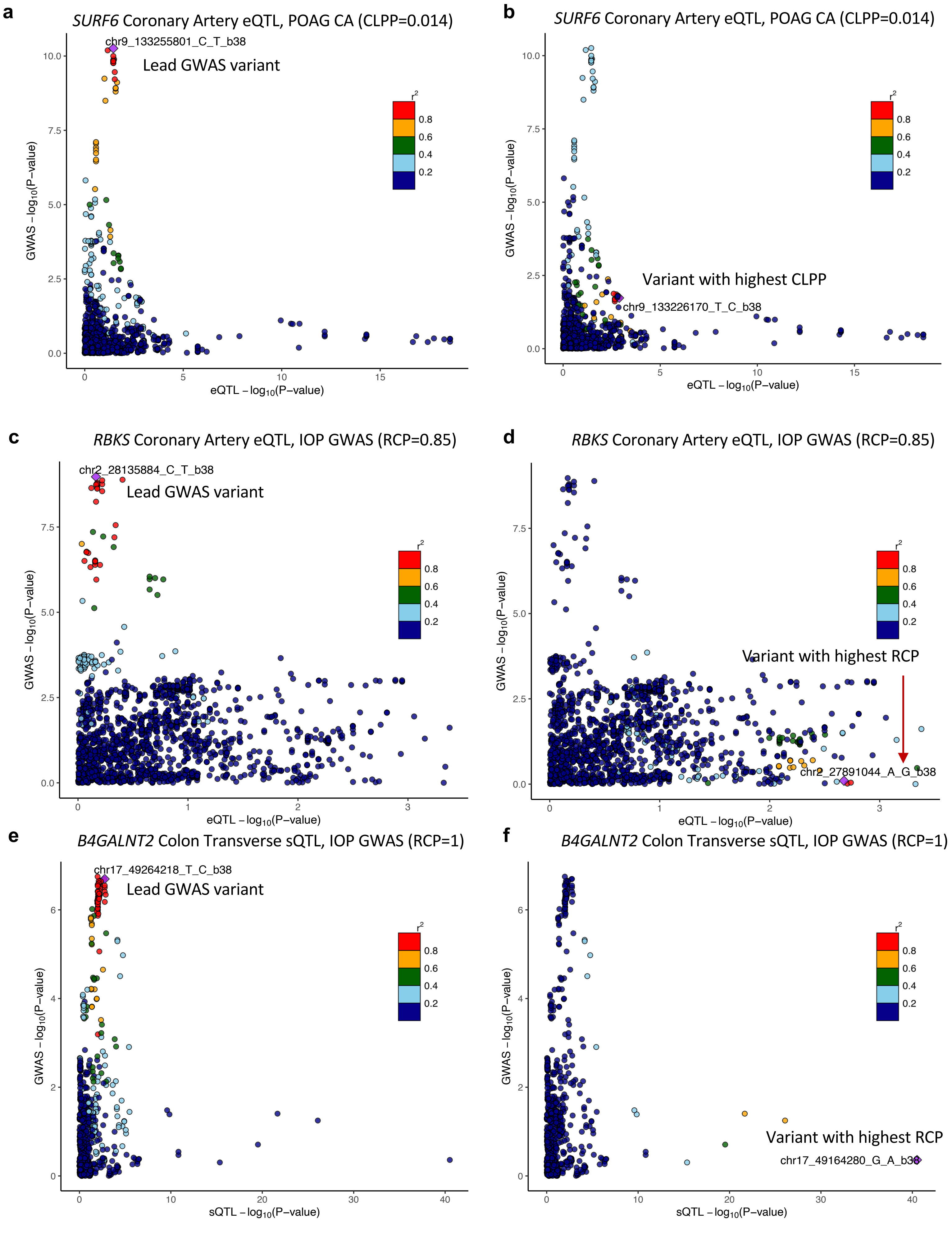

**Supplementary Figure 3. Examples of potential false positive colocalization results that were filtered out. a,b** LocusCompare plots of $-\log_{10}$ P-value of the POAG cross-ancestry (CA) GWAS as a function of the $-\log_{10}$ P-value of the *SURF6* coronary artery eQTL that displayed significant colocalization for the locus. The points representing different variants in the GWAS locus are color coded based on their LD ($r^2$) relative to the lead GWAS variant (chr9_133255801_C_T_b38) (**a**) or the eVariant (chr9_133226170_T_C_b38) with the most significant eCAVIAR colocalization posterior probability (CLPP=0.014) for the locus (**b**). It can be observed that the eQTL p-value of the lead GWAS variant is not significant, and the eQTL and GWAS p-values of the eVariant with the highest CLPP are only nominally significant. **c-f** LocusCompare plots of $-\log_{10}$ P-value of the IOP GWAS as a function of $-\log_{10}$ P-value of an *RBK4* coronary artery eQTL (**c,d**) or *B4GALNT2* colon transverse sQTL (**e,f**) which displayed significant colocalization with the corresponding locus. Points are color coded based on their LD ($r^2$) relative to the lead GWAS variant (**c,e**) or the eVariant with the highest *enloc* regional colocalization probability (RCP) (**d,f**) represented by a purple diamond. These two examples demonstrate an eQTL and sQTL that colocalize with an IOP GWAS locus with high confidence (RCP>0.85), though the variant with the highest RCP has a non-significant GWAS p-value.

### Supplementary Figure 4. Correlation of number of target genes of significantly colocalizing e/sQTLs with POAG and IOP GWAS loci with tissue sample size

**a**

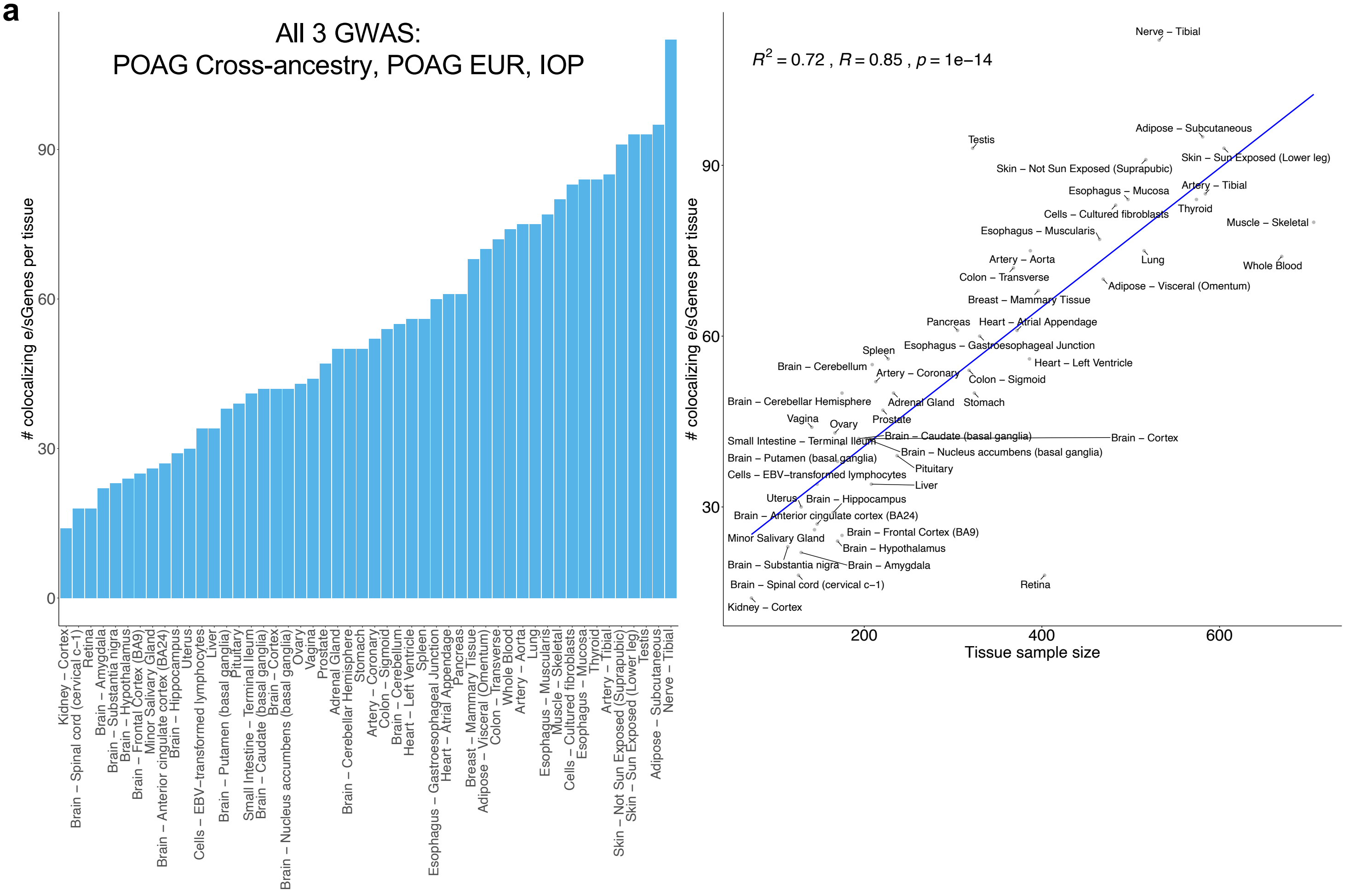


**b**

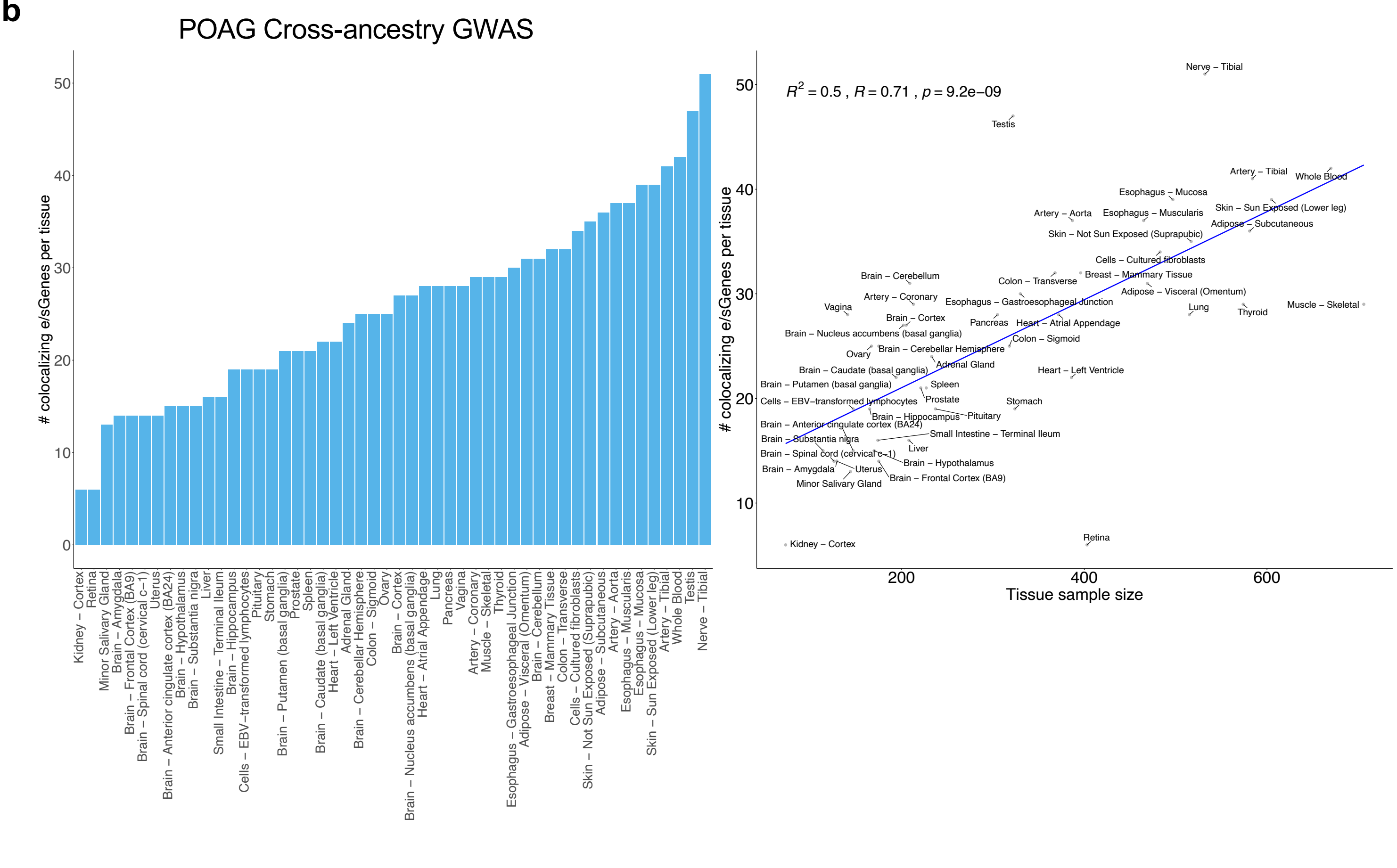

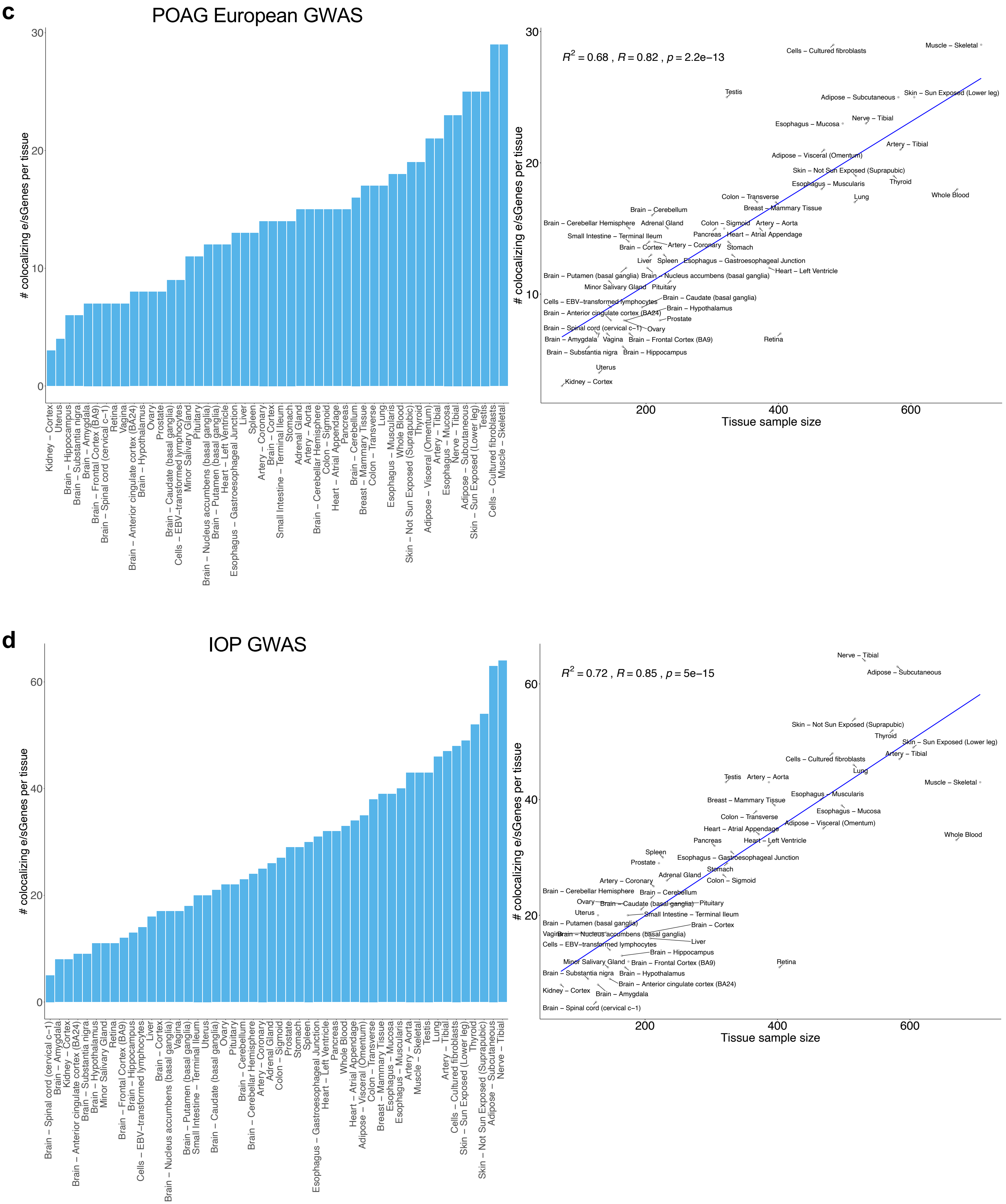


**Supplementary Figure 4: Correlation of number of target genes of significantly colocalizing e/sQTLs with POAG and IOP GWAS loci with tissue sample size.** Left panel: histogram of unique number of colocalizing e/sGenes per GTEx and retina tissue based on eCAVIAR (CLPP>0.01) and/or *enloc* (RCP>0.1). Right panel: scatter plot of number of colocalizing e/sGenes per tissue versus tissue sample size. Determination of coefficient ($R^2$), Pearson correlation coefficient (R), and p-value are shown. Blue line is a least squares fitted line. These were generated for the unique union of colocalizing e/sGenes with the POAG cross-ancestry, POAG European ancestry subset, and IOP GWAS loci (**a**), or only the POAG cross-ancestry **(b)**, POAG European subset (**c**), and IOP (**d**) GWAS loci.

#### Supplementary Figure 5. Colocalizing e/sQTLs with top POAG European and IOP GWAS loci

**a**

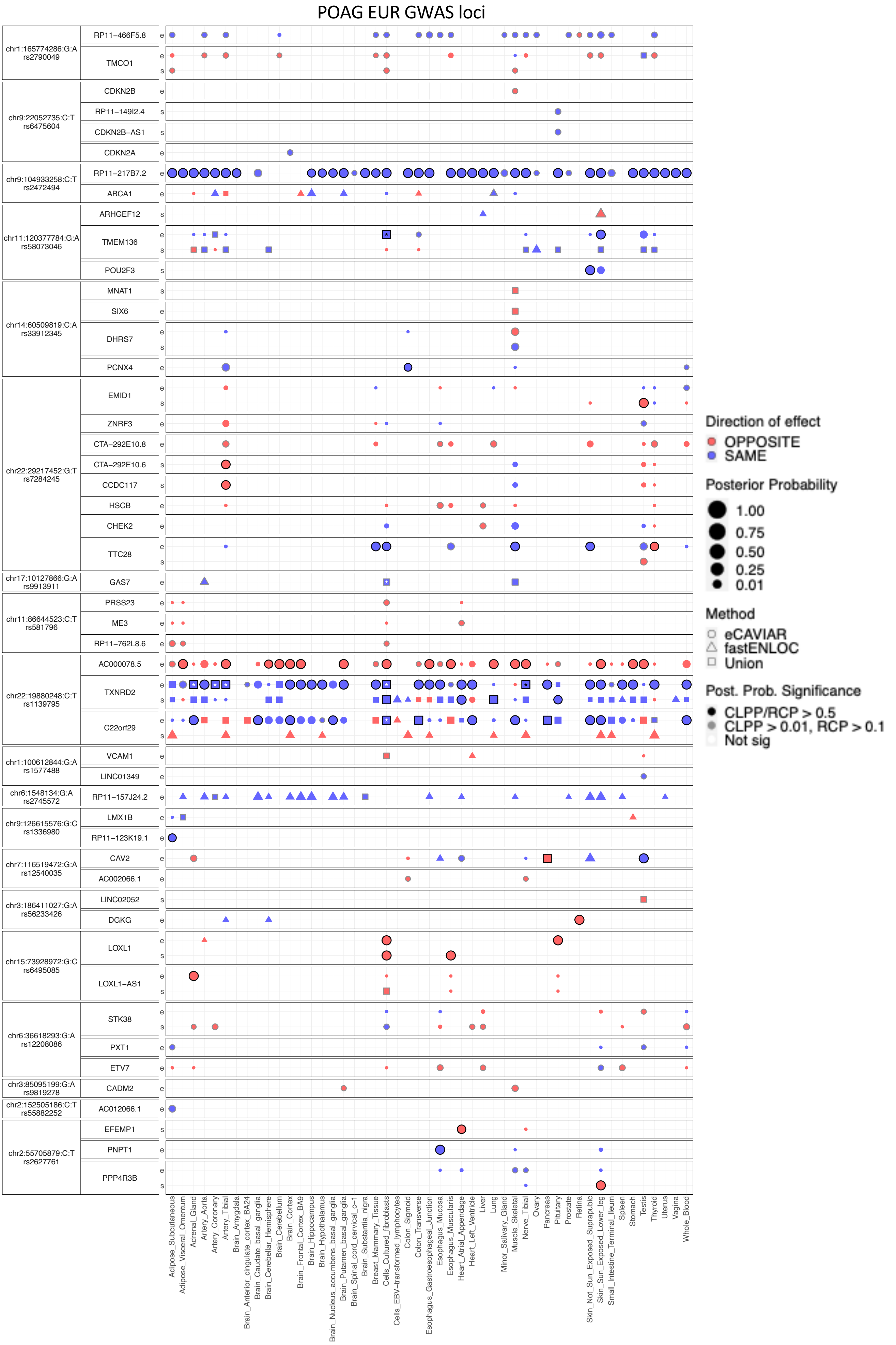

**b**

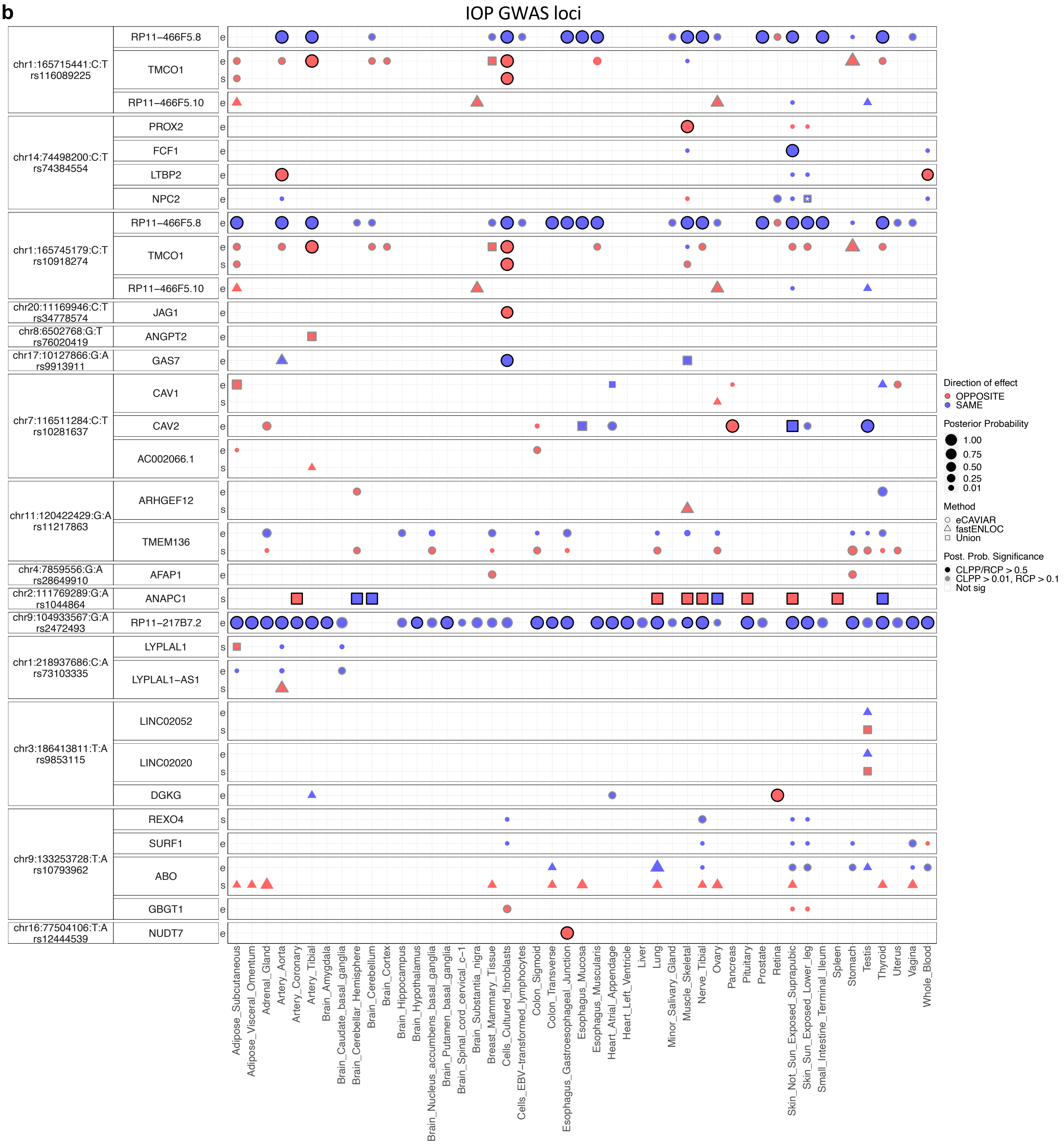


**Supplementary Figure 5. Colocalizing e/sQTLs with top POAG European GWAS and IOP GWAS loci. a-b**, Colocalization results for the top POAG EUR GWAS (**a**) and IOP EUR GWAS loci (**b**). Only genes with at least one significant colocalization result are shown for e/sQTLs tested across 49 GTEx tissues and peripheral retina per locus. GWAS loci were ordered by absolute value of their effect size in descending order, and within each locus, genes were ordered based on their chromosome position. Bubble size is proportional to the maximum colocalization posterior probability of all e/sVariants tested for the given gene, QTL type and tissue combination. Points are color-coded by direction of effect (blue if increased expression or splicing increases POAG risk or IOP levels or vice versa; red if increased expression or splicing decreases POAG risk or IOP levels or vice versa). Shape of points indicates colocalization method used: circle (eCAVIAR), triangle (*enloc*), and square (tested in both methods; results shown for method with maximum posterior probability). Grey or black border denotes variant-gene-tissue-QTL combination that passed QC filtering (Methods) and a colocalization posterior probability cutoff above 0.01/0.1 (CLPP/RCP) or 0.5 (higher confidence), respectively. White or black asterisk in the square indicates whether the second method tested passed a posterior probability cutoff 0.01/0.1 (CLPP/RCP) or 0.5, respectively.

**Supplementary Figure 6. *TMCO1* e/sQTLs colocalizing with POAG and IOP GWAS loci.**

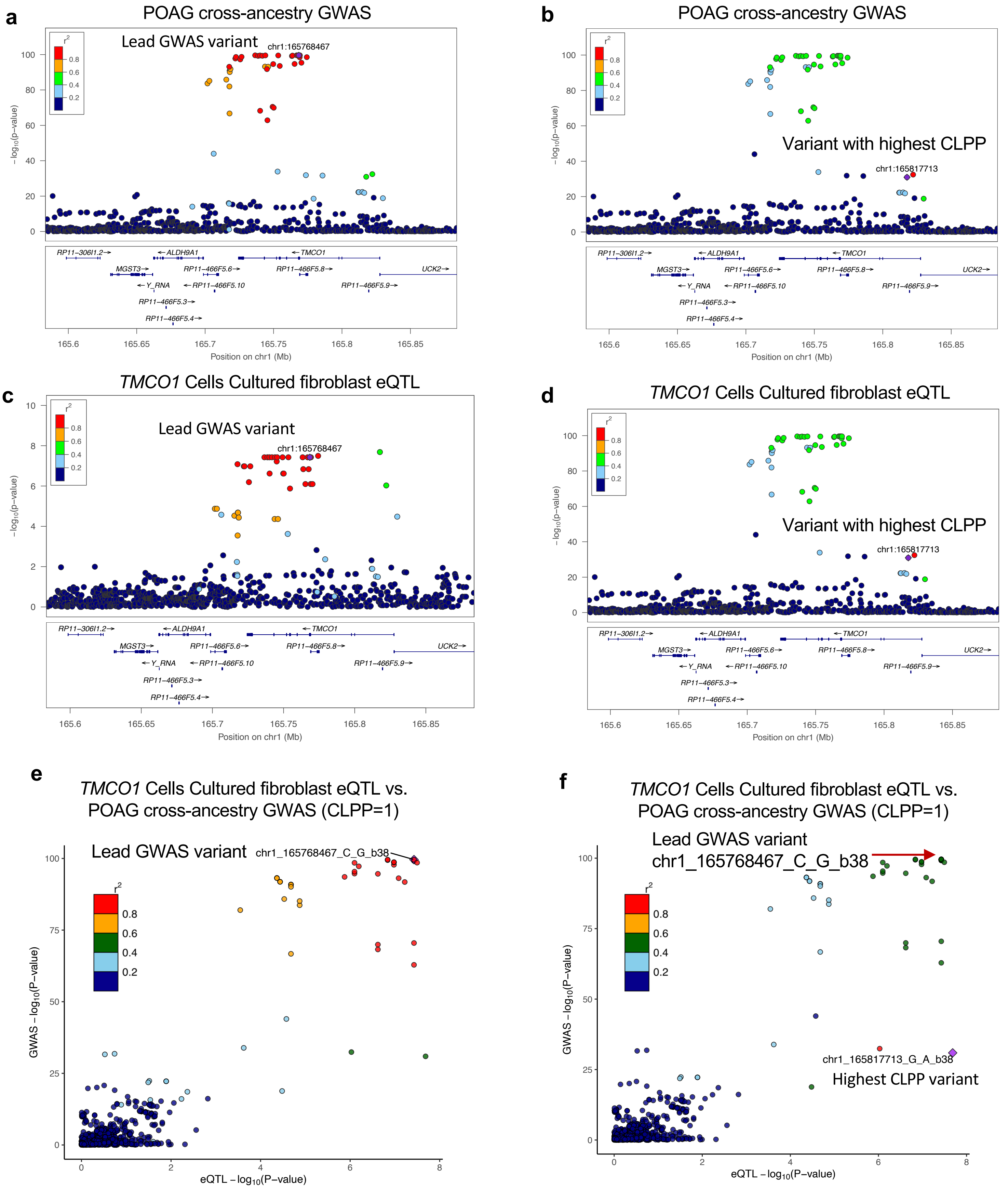

**Supplementary Figure 6. *TMCO1* e/sQTLs colocalizing with POAG and IOP GWAS loci.**

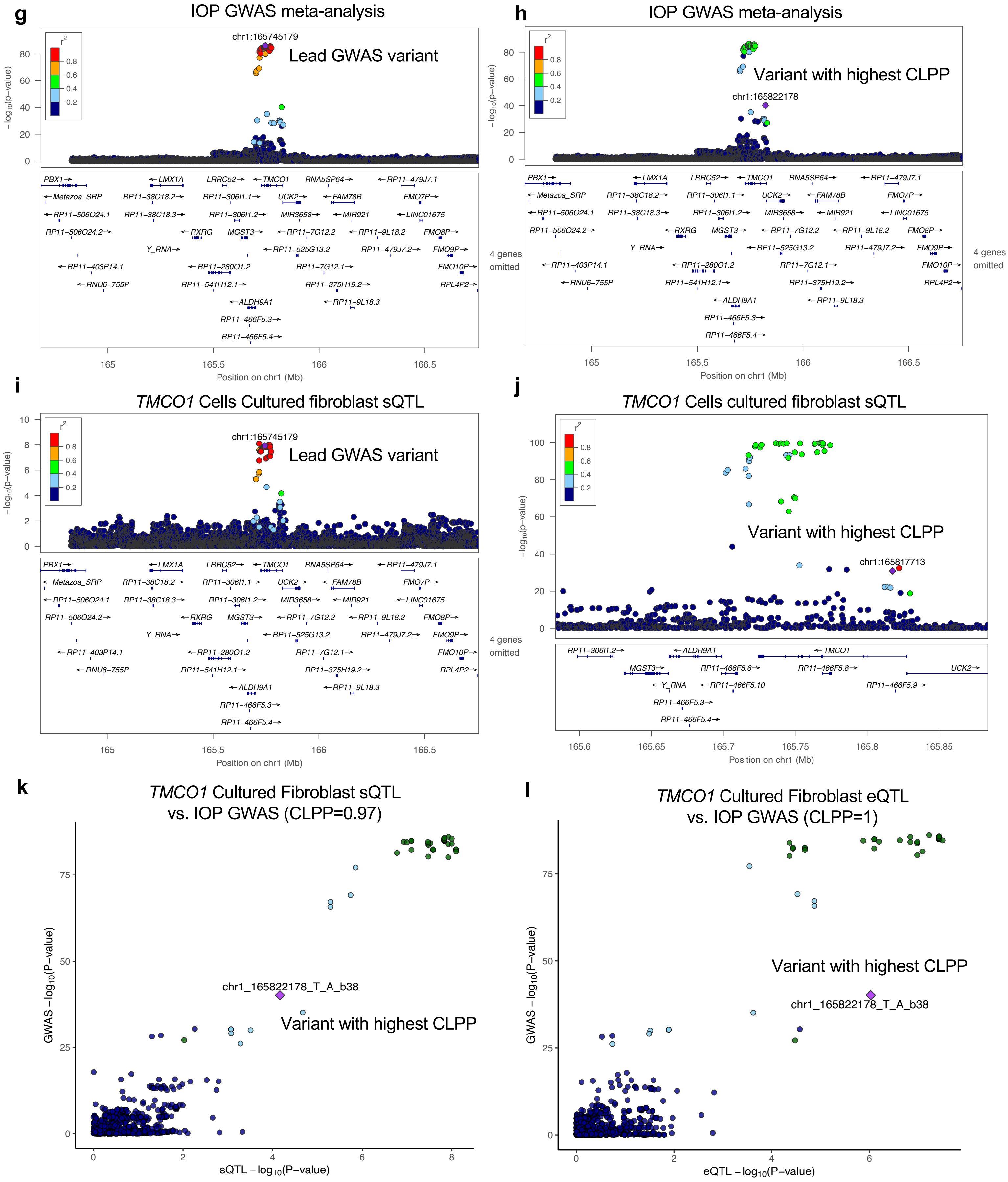

**Supplementary Figure 6. *TMCO1* e/sQTLs colocalizing with POAG and IOP GWAS loci. a-d**, LocusZoom plots for the POAG cross-ancestry GWAS p-values (**a,b**) and *TMCO1* Cell Cultured fibroblast eQTL p-values (**c,d**) on $-\log_{10}$ scale for the POAG locus rs2790053 (chr1_165768467_C_G_b38). Variants are color-coded by LD ($r^2$) relative to the lead GWAS variant (**a,c**) or the eVariant with the highest colocalization posterior probability (**b,d**). **e-f**, LocusCompare plots for *TMCO1* Cells Cultured fibroblast eQTL p-values relative to POAG cross ancestry GWAS p-values on $-\log_{10}$ scale for all variants in the GWAS locus LD interval. Variants are color-coded by LD relative to the lead GWAS variant chr1_165768467_C_G_b38 (**e**) or the variant with highest colocalization posterior probability (chr1_165817713_G_A_b38; CLPP=1) (**f**). **g-j**, LocusZoom plots for the IOP GWAS p-values (**g,h**) and *TMCO1* Cells Cultured fibroblast sQTL p-values (**i,j**) on $-\log_{10}$ scale for the IOP locus rs10918274 (chr1_165745179_T_C_b38). Variants are color-coded by LD ($r^2$) relative to the lead GWAS variant chr1_165745179_T_C_b38 (**g,i**) or the variant with the highest CLPP (chr1_165822178_T_A_b38; CLPP=0.97) (**h,j**). **k-l,** LocusCompare plots for *TMCO1* Cells Cultured fibroblast sQTL (**k**) or eQTL (**l**) p-values relative to IOP GWAS p-values on $-\log_{10}$ scale for all variants in the locus LD interval. Variants are color-coded by LD ($r^2$) relative to the variant with the highest colocalization posterior probability for the *TMCO1* (**k**) sQTL (CLPP=0.97) or (**l**) eQTL (CLPP=1) (chr1_165822178_T_A_b38).

**Supplementary Figure 7. *TMCO1* and *TMCO1-AS1* single-nucleus expression in anterior and posterior eye tissues.**

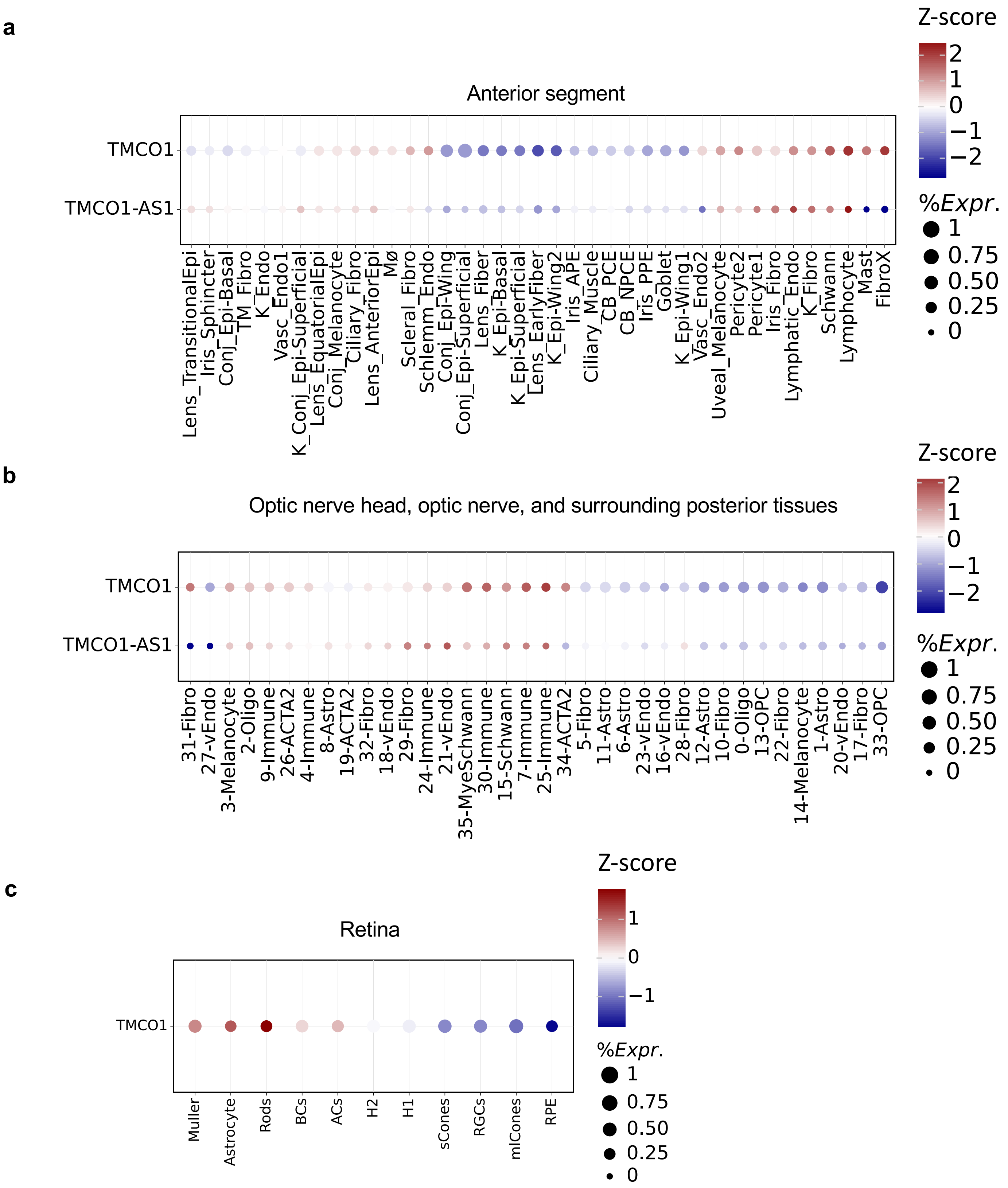


**Supplementary Figure 7. *TMCO1* and *TMCO1-AS1* single-nucleus expression in anterior and posterior eye tissues. a-d,** Bubble maps displaying the average expression of *TMCO1* and its anti-sense, *TMCO1-AS1* (*RP11-466F5.8*) across all cell types in six tissues in the anterior segment (**a**), optic nerve head and surrounding posterior tissues (**b**), and retina (**c**). *TMCO1-AS1* was not expressed in retina. The colorbar represents z-scores computed by comparing each gene's average expression in a given cell type to its per cell type average expression across all types divided by the standard deviation of all cell type expression averages. Bubble size is proportional to the percentage of cells expressing the given gene (log(TPK+1)). Cell type abbreviations are described in Supplementary Table 38.

**Supplementary Figure 8. Example of colocalizing e/sQTLs with IOP GWAS loci.**

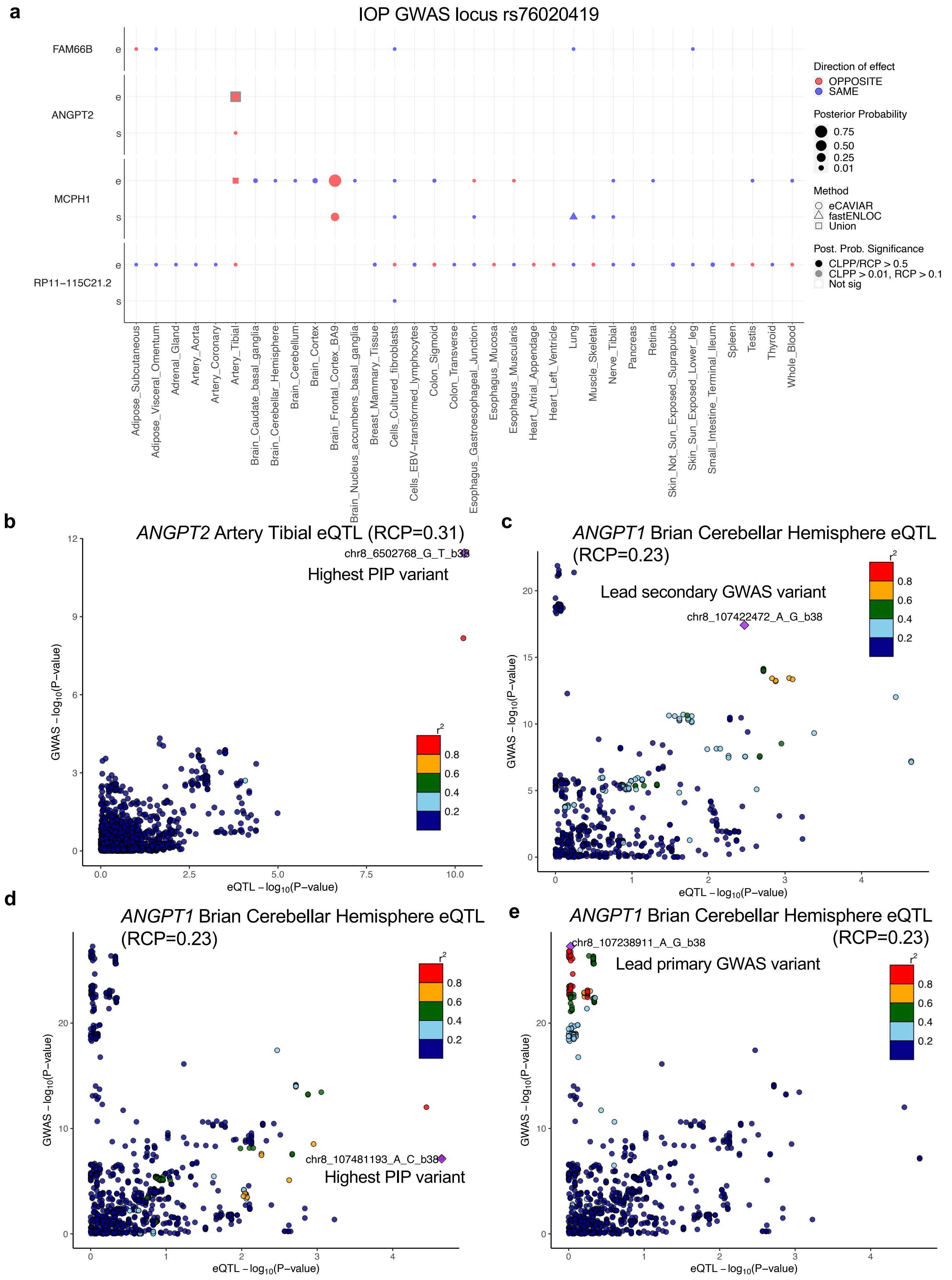

**Supplementary Figure 8. Example of colocalizing e/sQTLs with IOP GWAS loci. a,** Colocalization results for all e/sGenes tested in the IOP GWAS locus rs76020419 locus LD interval that had at least one significant eQTL or sQTL result with eCAVIAR or *enloc* across 49 GTEx tissues and peripheral retina. Genes were ordered by chromosome position. Size of points is proportional to the maximum colocalization posterior probability of all e/sVariants tested for the given gene, QTL type and tissue combination. Points are color-coded by direction of effect (blue if increased expression or splicing increases IOP levels or vice versa; red if increased expression or splicing decreases IOP levels or vice versa). Shape of points indicates colocalization method used: circle (eCAVIAR), triangle (*enloc*), and square (tested in both methods; results shown for method with maximum posterior probability). Grey or black border denote variant-gene-tissue-QTL combination that passed QC filtering (Methods) and a colocalization posterior probability cutoff above 0.01/0.1 (CLPP/RCP) or 0.5, respectively. White or black asterisk in the square indicates whether the second method tested passed a posterior probability cutoff of 0.01/0.1 (CLPP/RCP) or 0.5, respectively. **b-e,** LocusCompare plot of -$\log_{10}$(P-value) of the IOP GWAS relative to the -$\log_{10}$(P-value) of artery tibial eQTL acting on *ANGPT2* (**b**) or brian cerebellar hemisphere eQTL acting on *ANGPT1* (**c-e**). Points are color-coded based on LD ($r^2$) relative to the lead GWAS variant (**c,e**) or the variant with highest *enloc* posterior inclusion probability (PIP) (**b,d**). Interestingly, the eQTL acting on *ANGPT1 in* brain cerebellar hemisphere significantly colocalized (RCP=0.23) with the secondary independent IOP GWAS variant rs4496939 (chr8:107422472:A:G, beta = -0.118, P=$2.7 \times 10^{-18}$) (**c,d**), and not with the stronger independent IOP signal rs2022945 (chr8:107238911:G:A, beta = -0.213, P=$1.1 \times 10^{-28}$) (**e**) of the three independent signals in the locus.

**Supplementary Figure 9. *GAS7* eQTL colocalizing with POAG and IOP GWAS loci.**

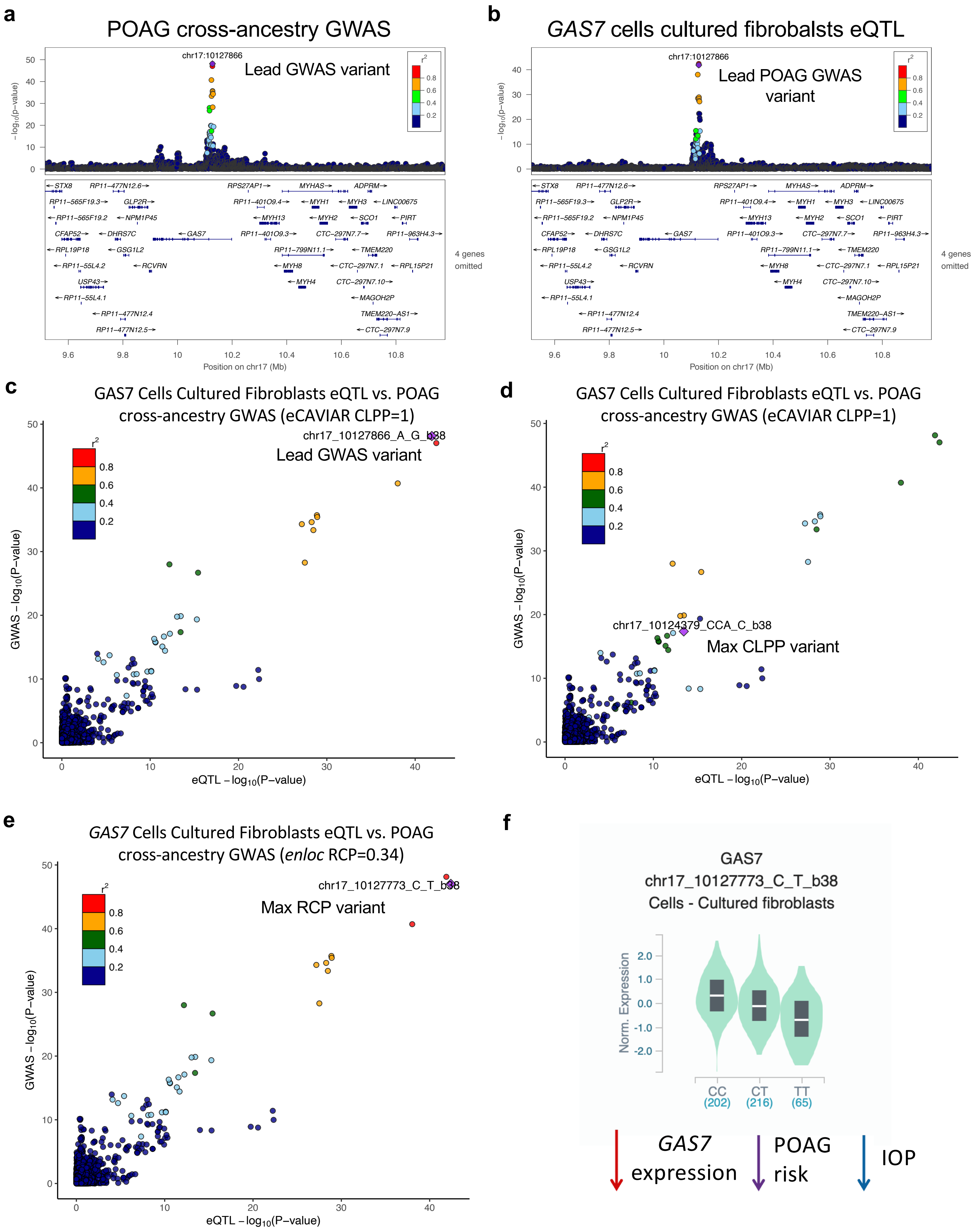

g

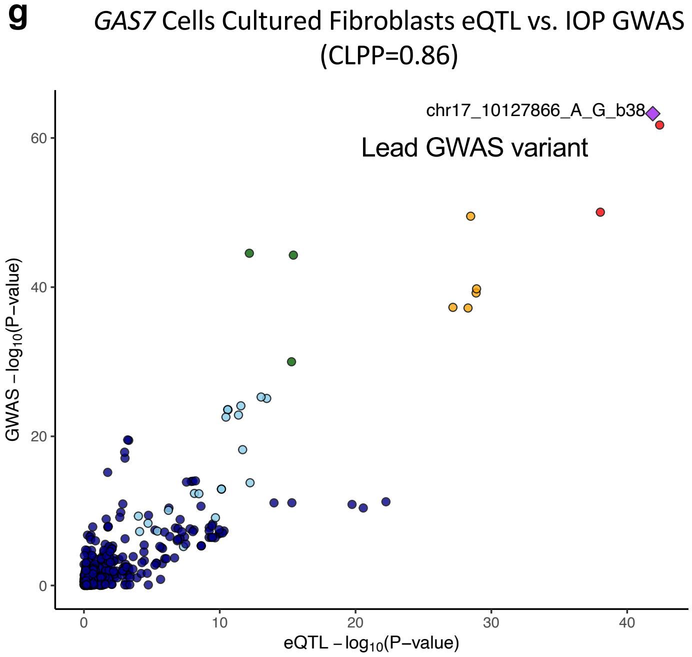


h

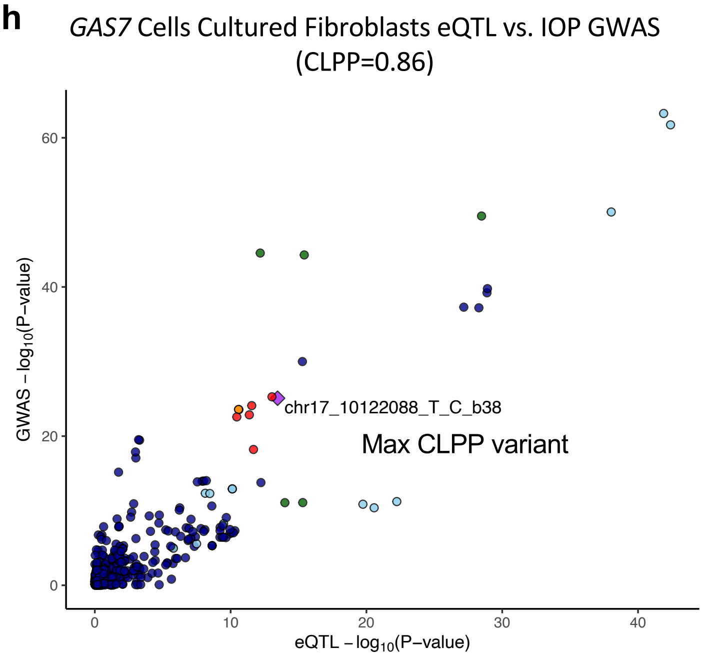


**Supplementary Figure 9. *GAS7* eQTL colocalizing with POAG and IOP GWAS loci. a-d**, LocusZoom plots for the POAG cross-ancestry GWAS p-values (**a**) and *GAS7* Cells Cultured fibroblast eQTL (**b**) -$\log_{10}$(P-values) for the POAG cross-ancestry locus with lead variant rs9913911 (chr17_10127866_A_G_b38). Variants are color-coded by LD ($r^2$) relative to the lead GWAS variant. **c-e,g**, LocusCompare plots for -$\log_{10}$(P-value) of POAG cross ancestry GWAS (**c-e**) or IOP GWAS (**g**) compared to *GAS7* Cells Cultured fibroblast eQTL -$\log_{10}$(P-value) for all variants in the GWAS locus LD interval including eVariants that significantly colocalized with POAG and IOP. Variants are color-coded by LD ($r^2$) relative to the lead GWAS variant (**c,g**) or the variant with highest colocalization posterior probability based on eCAVIAR (**d,h**) or enloc (**e**). **f,** Violin plot of normalized *GAS7* expression values in Cells Cultured fibroblasts as a function of the genotype of the eVariant chr17_10127773_C_T_b38 (rs12150284) that had the highest RCP value, taken from the GTEx portal (https://gtexportal.org). The effect size of this eVariant relative to the alternative allele (eQTL *beta* = -0.44) is in the same direction relative to POAG risk (POAG cross-ancestry GWAS beta = -0.14), suggesting that decreased *GAS7* expression is protective for POAG.

#### Supplementary Figure 10. *ABO* eQTL colocalizing with POAG and IOP GWAS loci

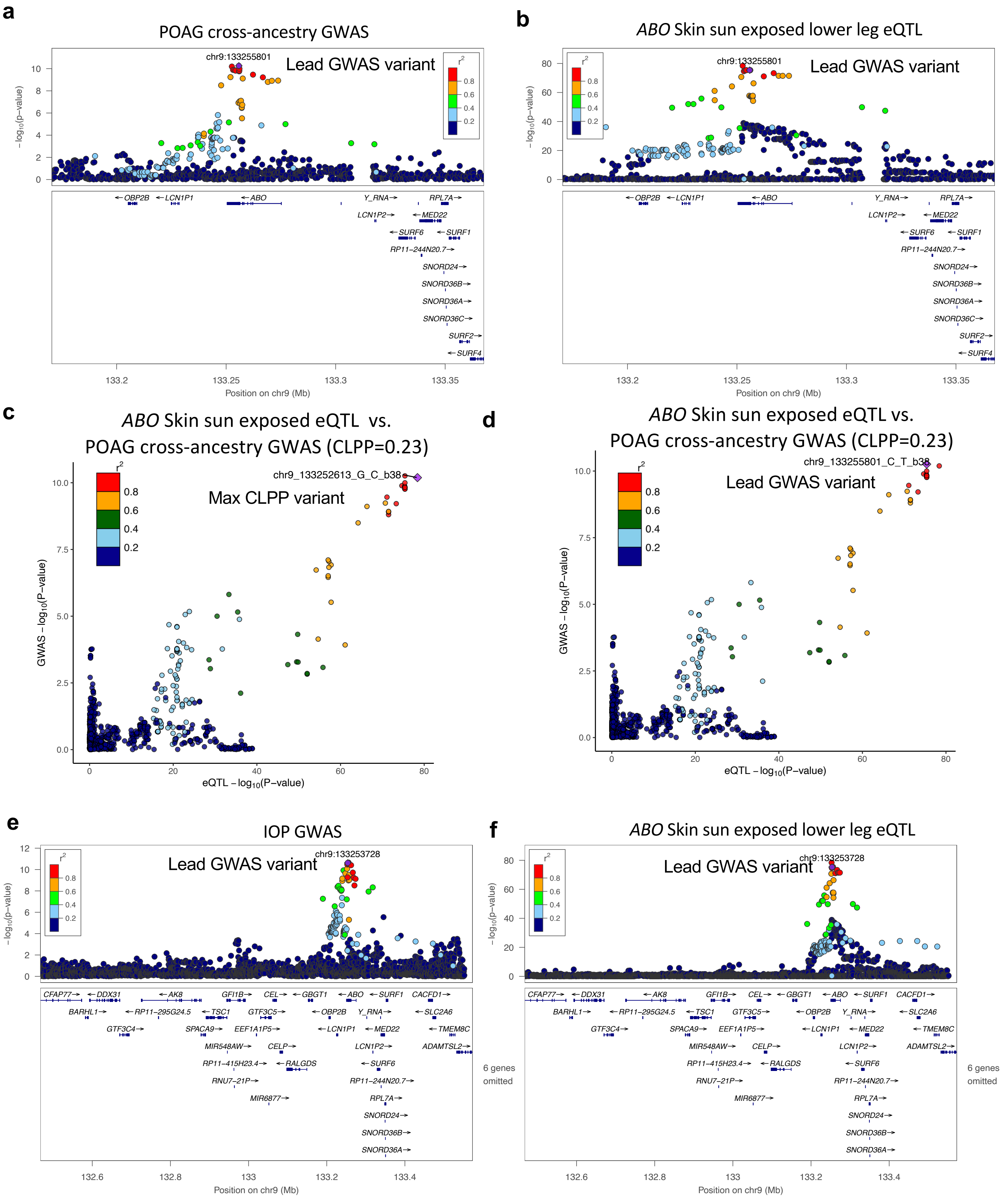

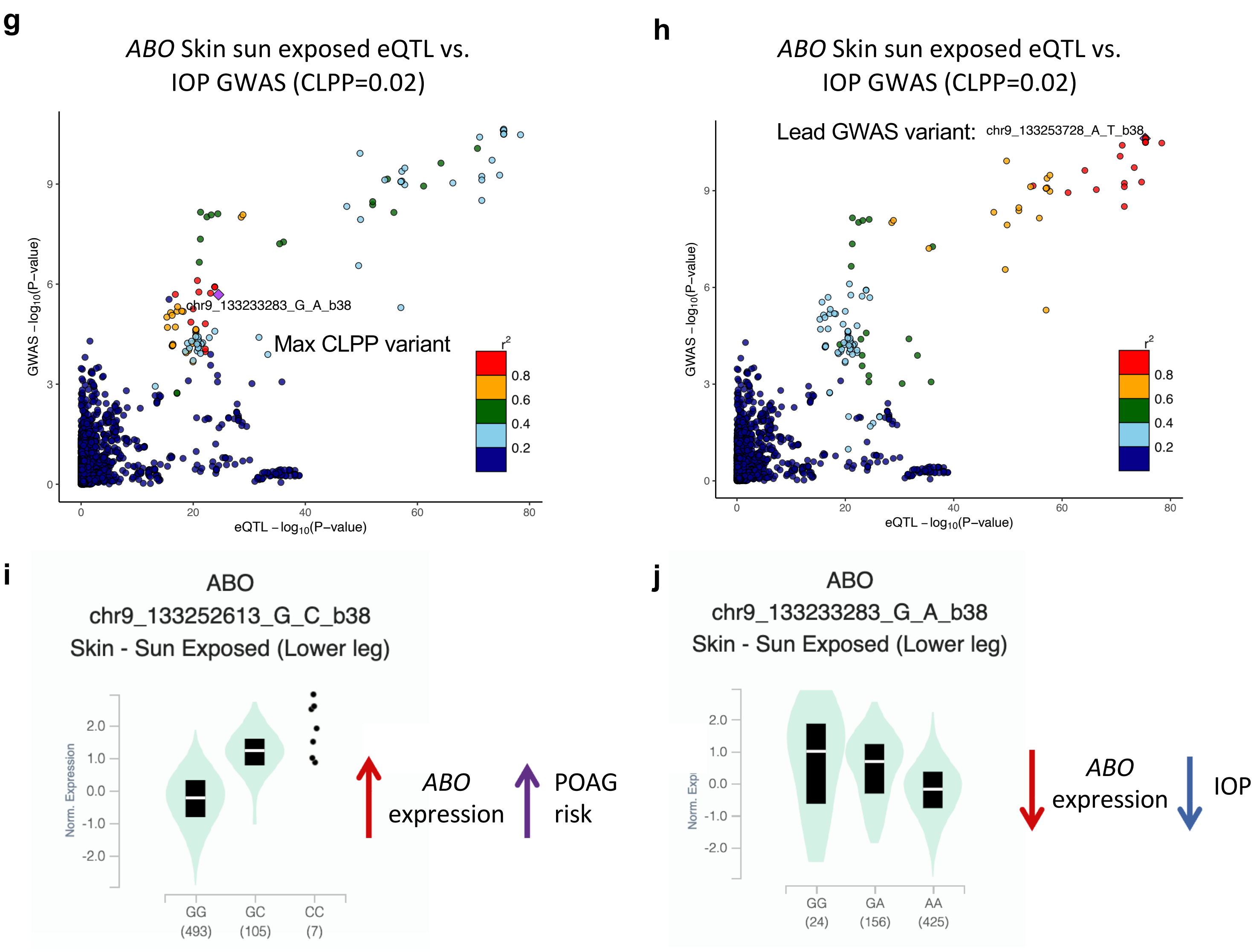


**Supplementary Figure 10. *ABO* eQTL colocalizing with POAG and IOP GWAS loci. a-b**, LocusZoom plots of -log$_{10}$(P-value) of the POAG cross-ancestry GWAS meta-analysis (**a**) and *ABO* Skin sun exposed (lower leg) eQTL (**b**) for POAG locus rs8176749 (chr9:133255801_C_T_b38). Variants are color-coded by LD (r$^2$) relative to the lead GWAS variant (chr9_133255801_C_T_b38). **c-d,** LocusCompare plots of -log$_{10}$(P-value) of the POAG cross-ancestry GWAS meta-analysis compared to the *ABO* Skin sun exposed lower leg eQTL that significantly colocalized with POAG GWAS association in locus rs8176749. Points are color-coded based on their LD (r$^2$) relative to the variant with the highest colocalization posterior probability (chr9_133252613_G_C_b38; CLPP=0.23) (**c**) or the lead GWAS variant (chr9_133255801_C_T) (**d**). **e-f**, LocusZoom plots of -log$_{10}$(P-value) of IOP GWAS meta-analysis (**e**) and *ABO* Skin sun exposed lower leg eQTL (**f**) for the IOP locus rs10793962 (chr9_133253728_T_A_b38). Variants are color-coded by LD (r$^2$) relative to the lead GWAS variant (chr9_133253728_T_A_b38). **g-h,** LocusCompare plots of -log$_{10}$(P-value) of IOP GWAS meta-analysis compared to *ABO* Skin sun exposed lower leg eQTL that significantly colocalized with the IOP GWAS locus rs10793962. Points are color-coded based on their LD (r$^2$) relative to the variant with the highest colocalization posterior probability (chr9_133233283_G_A_b38; CLPP=0.02) (**f**) or the lead GWAS variant (chr9_133253728_T_A_b38) (**h**). **i-j,** Violin plot of normalized *ABO* expression values in Skin sun exposed (lower leg) as a function of the genotype of the eVariant chr9_133252613_G_C_b38 or chr9_133233283_G_A_b38 that displayed the highest colocalization posterior probability for POAG cross-ancestry (**i**) and IOP (**j**) GWAS, respectively, taken from the GTEx portal (https://gtexportal.org). The effect size of this *ABO* eVariant relative to the alternative allele, *beta* = 1.35 and -0.67 for POAG (**i**) and IOP (**j**), respectively is in the same direction relative to POAG risk (POAG cross-ancestry GWAS beta = 0.096) or IOP level (GWAS beta = -0.094), respectively, suggesting that decreased *ABO* expression may be protective for POAG.

**Supplementary Figure 11. e/sQTL colocalization results for *CDKN2A/B* POAG cross-ancestry GWAS locus**

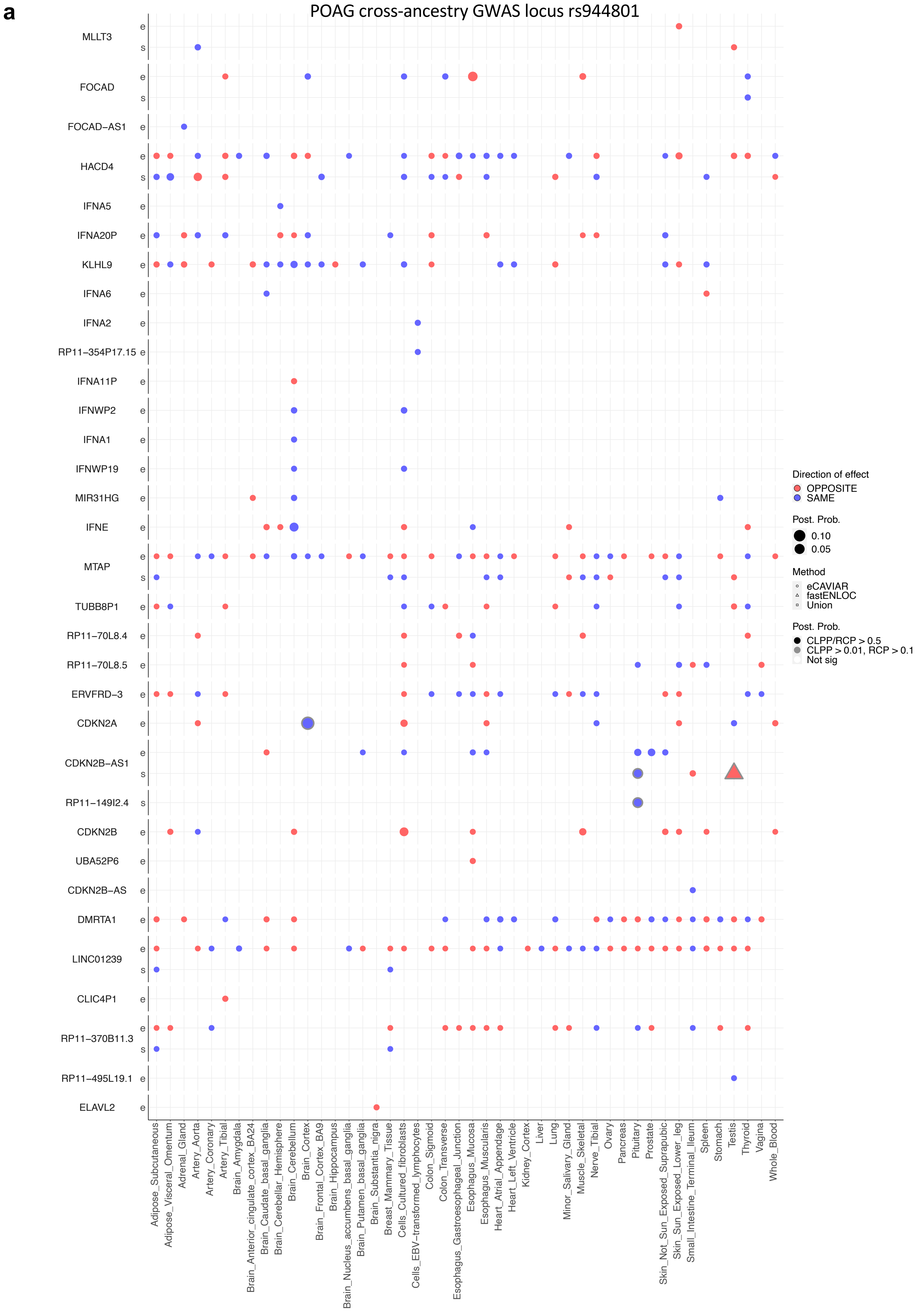

#### Supplementary Figure 11. e/sQTL colocalization results for *CDKN2A/B* POAG cross-ancestry GWAS locus

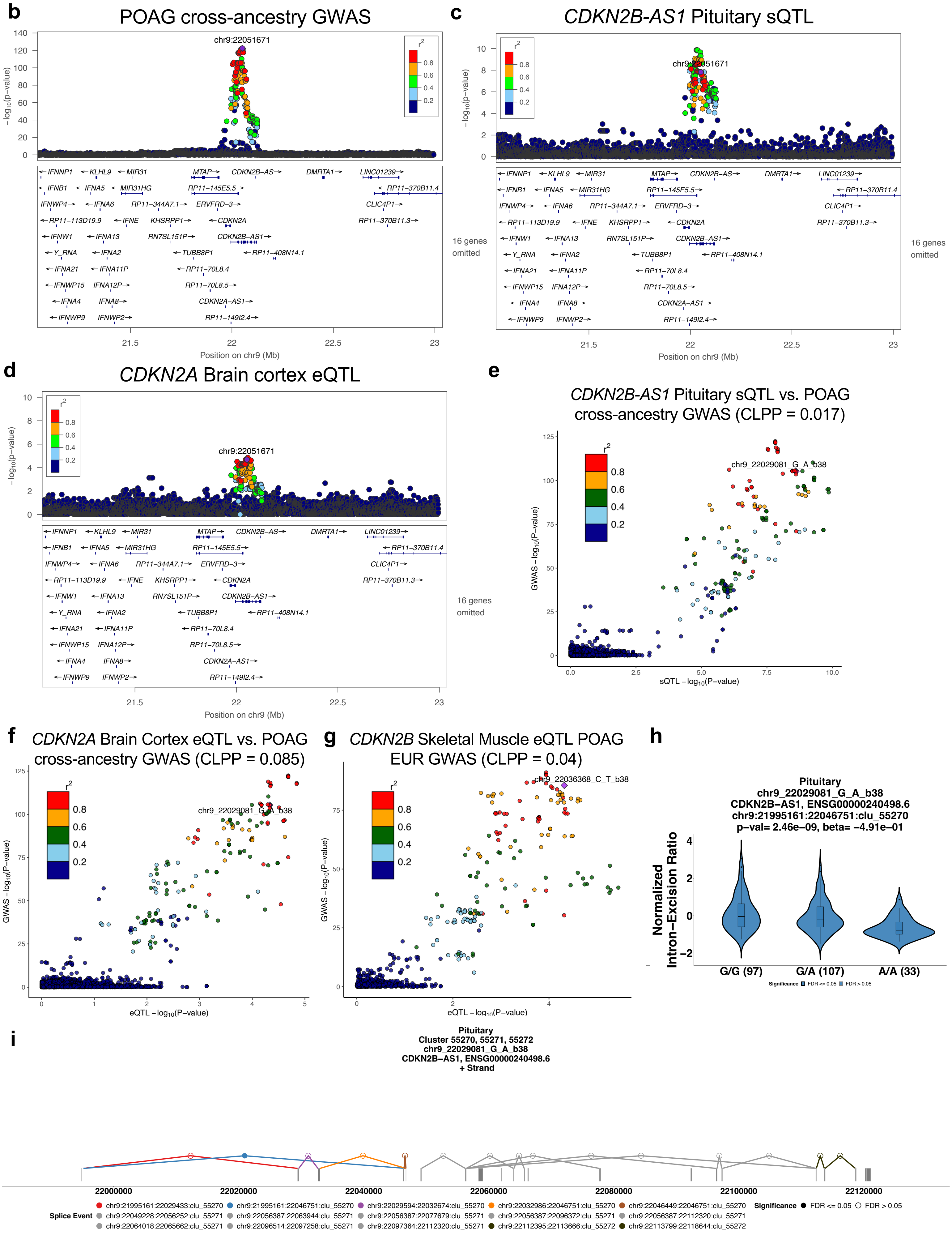

**Supplementary Figure 11. e/sQTL colocalization results for *CDKN2A/B* POAG cross-ancestry locus. a,** Colocalization results for all genes tested in the POAG cross-ancestry GWAS locus rs944801 (chr9_22051671_G_C_b38) LD interval that had at least one significant eQTL or sQTL result across 49 GTEx tissues and peripheral retina. Genes were ordered by chromosome position. Size of points is proportional to the maximum colocalization posterior probability of all e/sVariants tested for the given gene, QTL type and tissue combination. Points are color-coded by direction of effect (blue if increased expression or splicing increases POAG risk or vice versa; red if increased expression or splicing decreases POAG risk or vice versa). Shape of points indicates colocalization method used: circle (eCAVIAR), triangle (*enloc*), and square (tested in both methods; results shown for method with maximum posterior probability). Grey or black border denote variant-gene-tissue-QTL combination that passed QC filtering (see Methods) and a colocalization posterior probability cutoff above 0.01/0.1 (CLPP/RCP) or 0.5, respectively. White or black asterisk in the square indicates whether the second method tested passed a posterior probability cutoff of 0.01/0.1 (CLPP/RCP) or 0.5, respectively. **b-d**, LocusZoom plots for the POAG GWAS locus rs944801 showing the $-\log_{10}$(P-value) of the POAG cross-ancestry GWAS (b), *CDKN2B-AS1* pituitary sQTL (c), and *CDKN2A* Brain cortex eQTL (**d**) in the GWAS locus LD interval. Points are color-coded by LD relative to the lead GWAS variant (chr9_22051671_G_C_b38). **e-g,** LocusCompare plots of $-\log_{10}$(P-value) of the POAG cross-ancestry GWAS meta-analysis relative to the $-\log_{10}$(P-value) of the *CDKN2B-AS1* pituitary sQTL (**e**) or *CDKN2A* Brain cortex eQTL (**f**), and of the POAG European subset GWAS versus the *CDKN2B* Skeletal muscle eQTL (**g**), all of which showed significant colocalization results. Points are color-coded based on LD ($r^2$) relative to the e/sVariant with the highest eCAVIAR colocalization posterior probability (CLPP). **h,** Violin plot of normalized intron-excision ratio for chr9:21995161-22046751 computed with Leafcutter for *CDKN2B-AS1* in Pituitary as a function of the genotype of the sVariant rs679038 (chr9_22029081_G_A_b38) that displayed the highest CLPP (0.017) for this sQTL and POAG cross-ancestry GWAS signal in locus rs944801 (chr9_22051671_G_C_b38). This sVariant also significantly colocalized with the POAG European subset GWAS locus rs6475604 (chr9_22052735_T_C_b38) (CLPP=0.11). The effect size of the sQTL relative to the alternative allele (*beta* = -0.49) is in same direction relative to POAG risk (POAG cross-ancestry GWAS beta = -0.22; POAG European GWAS beta = -0.24), suggesting that decrease in splicing between chr9:21995161-22046751 may decrease POAG risk. **i,** Gene model for *CDKN2B-AS1* in GTEx Pituitary showing all splicing events detected with Leafcutter for *CDKN2B-AS1* in this tissue (positive strand). Intron excision cluster 55270 is colored because it has a significant splicing event, chr9:21995161-22046751 (color Blue), which skips exon 2 and 3 that is associated with the sVariant rs679038 (chr9_22029081_G_A_b38) that colocalized with the POAG cross-ancestry GWAS locus rs944801 (chr9_22051671_G_C_b38), as well as with the POAG European GWAS locus rs6475604 (chr9_22052735_T_C_b38). Retention of exon 2 and 3 in *CDKN2B-AS1* is protective for POAG (**h**).

#### Supplementary Figure 12. *EFEMP1* sQTL colocalizing with POAG GWAS locus.

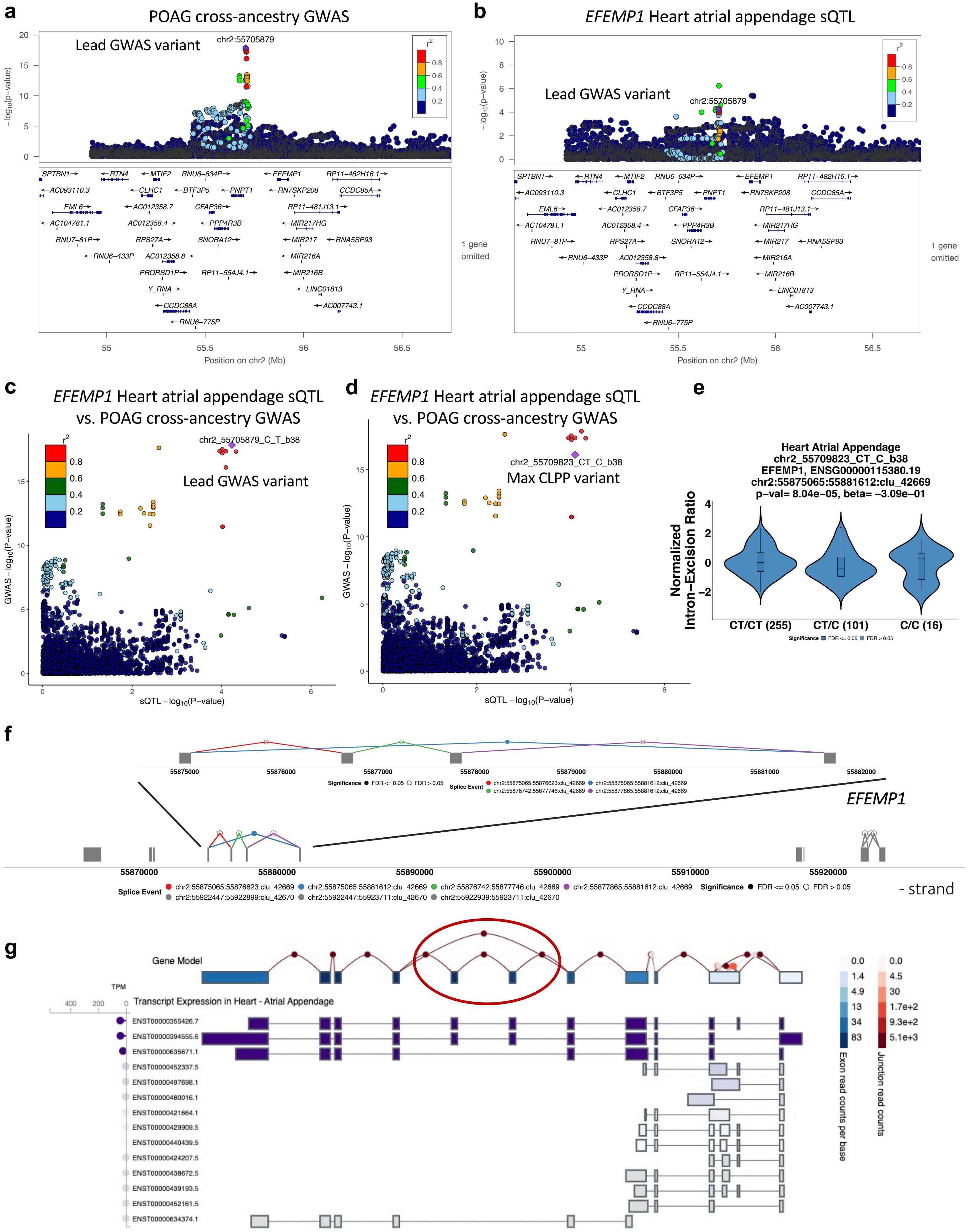

**Supplementary Figure 12. *EFEMP1* sQTL colocalizing with POAG GWAS locus. a-b**, LocusZoom plots of -$\log_{10}$(P-value) of POAG cross-ancestry GWAS meta-analysis (**a**) and *EFEMP1* Heart atrial appendage sQTL (**b**) that colocalized with the POAG cross-ancestry association in locus rs2627761 (chr2_55705879_C_T_b38). Variants are color-coded by LD ($r^2$) relative to the lead GWAS variant (chr2_55705879_C_T_b38). **c-d,** LocusCompare plots of -$\log_{10}$(P-value) of the POAG cross-ancestry GWAS meta-analysis compared to the *EFEMP1* Heart atrial appendage sQTL -$\log_{10}$(P-value). Points are color-coded based on their LD ($r^2$) relative to the lead GWAS variant (chr2_55705879_C_T_b38) (**c**) or the sVariant with the highest colocalization posterior probability (chr2_55709823_CT_C_b38**;** CLPP=0.51) (**d**). **e**, Violin plot of normalized intron-excision ratio for chr2:55875065-55881612 computed with Leafcutter for *EFEMP1* in Heart Atrial Appendage as a function of the genotype of the sVariant with the highest CLPP for this sQTL (rs35017406, chr2_55709823_CT_C_b38) and POAG GWAS locus rs2627761 (CLPP=0.51). The effect size of the sQTL relative to the alternative allele (*beta* = -0.31) is in opposite direction relative to the POAG cross-ancestry association (*beta* = 0.10), suggesting that increase in splicing between chr2:55875065-55881612 decreases risk of POAG. **f,** Gene model for *EFEMP1* in GTEx Heart Atrial Appendage on the negative strand, showing all splicing events in the gene (open circles), and zooming in on the splice event chr2:55875065-55881612 (cluster:42669) from exon 5 to exon 8 (blue line and closed circle) that has an sQTL (chr2_55709823_CT_C_b38) that significantly colocalized with POAG cross-ancestry association in the locus. From panel **e** and Supplementary Table 2, it can be understood that skipping of exons 6 and 7 of *EFEMP1* decreases risk of POAG. **g,** *EFEMP1* gene model and transcripts expressed in GTEx Heart Atrial Appendage taken from the GTEx portal (URLs). In the gene model, exon boxes are color-coded by exon read counts per base (blue) and lines connecting exons by exon-exon junction read counts (red). All splicing events observed in the tissue are shown, including the alternative splicing between exon 5 and exon 8 in *EFEMP1* whose genetic regulation colocalized with POAG (**d**). Below the gene model, transcripts expressed in Heart Atrial Appendage in Transcripts per Million (TPM), computed with RSEM, are shown in descending order.

#### **Supplementary Figure 13.** Top GWAS POAG locus with *MYOC* mutation colocalizes with *PIGC* sQTL.

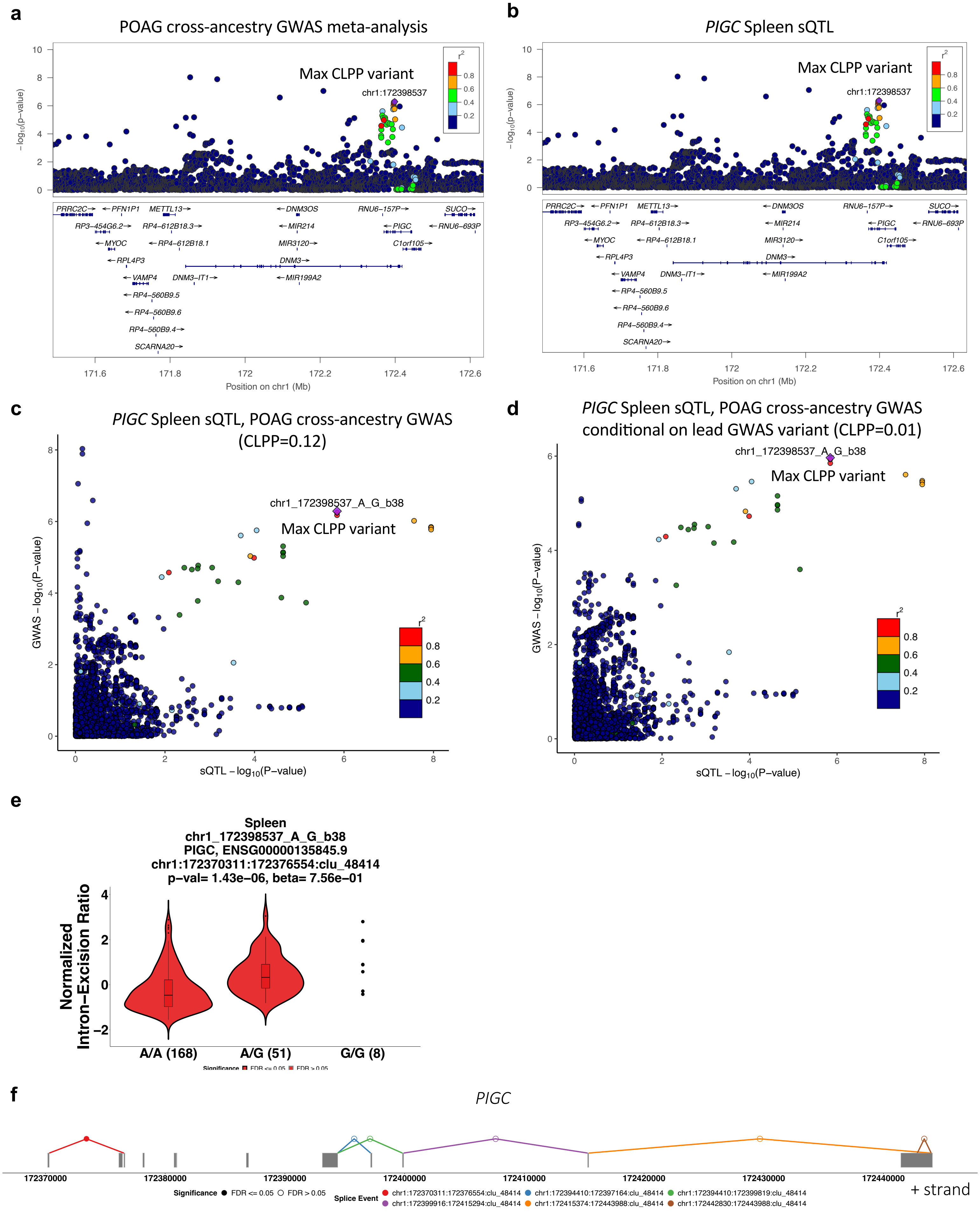

**Supplementary Figure 13. Top GWAS POAG locus with *MYOC* mutation colocalizes with *PIGC* sQTL. a-b**, LocusZoom plots of -$log_{10}$(P-value) of POAG cross-ancestry GWAS meta-analysis (**a**) and *PIGC* Spleen sQTL (**b**) that colocalized with the POAG cross-ancestry association in locus rs74315329 (chr1_171636338_G_A_b38). Variants are color-coded by LD ($r^2$) relative to the sVariant with the highest colocalization posterior probability (chr1_172398537_A_G_b38; CLPP=0.12). **c-d,** LocusCompare plots of -$log_{10}$(P-value) of the *PIGC* Spleen sQTL compared to POAG cross-ancestry GWAS meta-analysis before (**c**) and after (**d**) conditional analysis on the lead GWAS variants 74315329 (see Methods). Points are color-coded based on their LD ($r^2$) relative to the variant with the highest CLPP (chr1_172398537_A_G_b38). **e**, Violin plot of normalized intron-excision ratio for chr1:172370311-172376554 computed with Leafcutter for *PIGC* in Spleen as a function of the genotype of the sVariant rs10911684 (chr1_172398537_A_G_b38) with the highest CLPP for this sQTL and POAG locus. The effect size of the sQTL relative to the alternative allele (*beta* = 0.76) is in opposite direction relative to the POAG cross-ancestry association (*beta* = -0.085), suggesting that increased exon 2 skipping is protective of POAG. **f**, Gene model for *PIGC* in GTEx Spleen on positive strand, showing all splicing events in the gene (open circles), including the splice event chr1:172370311-172376554 (from exon 1 to exon 3) with a significant sQTL (chr1_172398537_A_G_b38) that colocalized with POAG locus rs74315329 (Red line and closed circle).

**Supplementary Figure 14. Top gene sets enriched for e/sGenes that colocalized with POAG and IOP GWAS loci.**

**a**

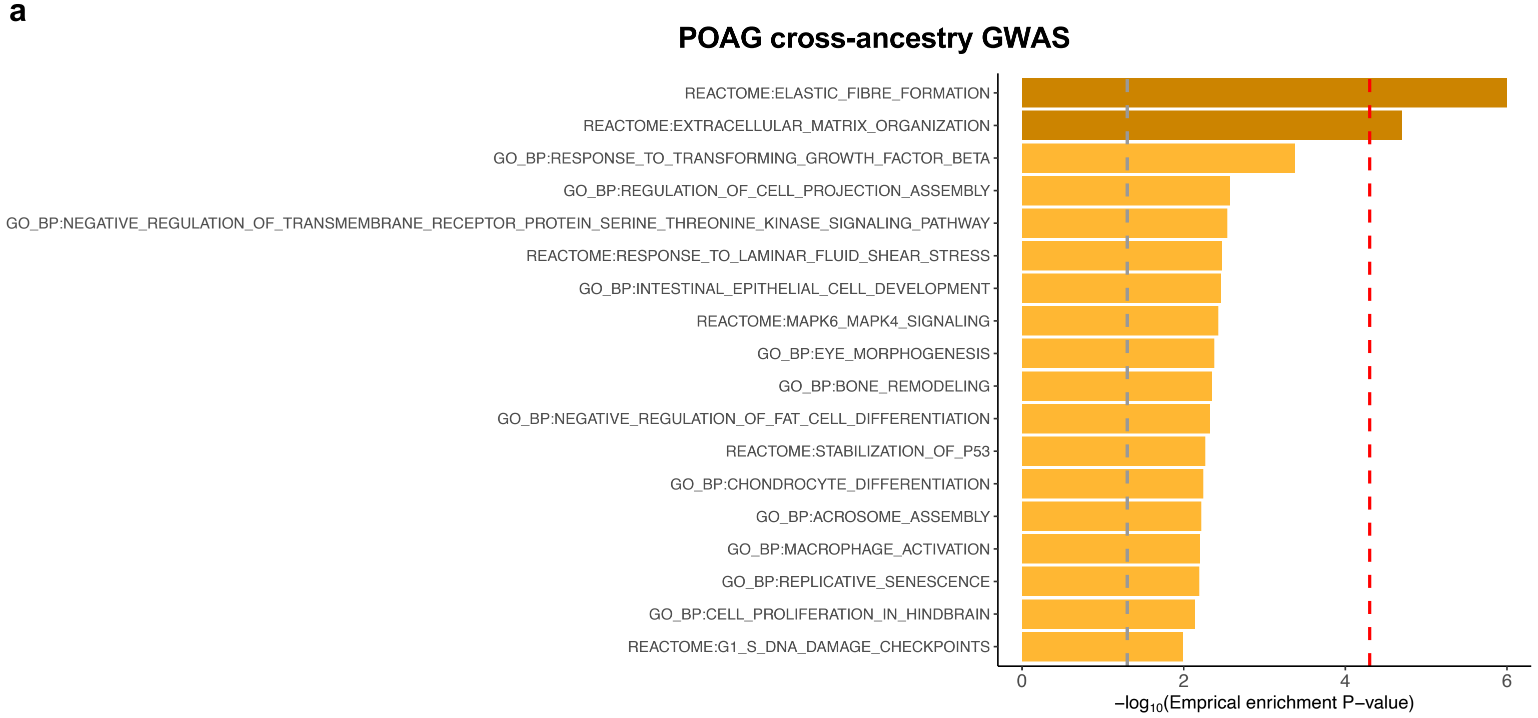


**b**

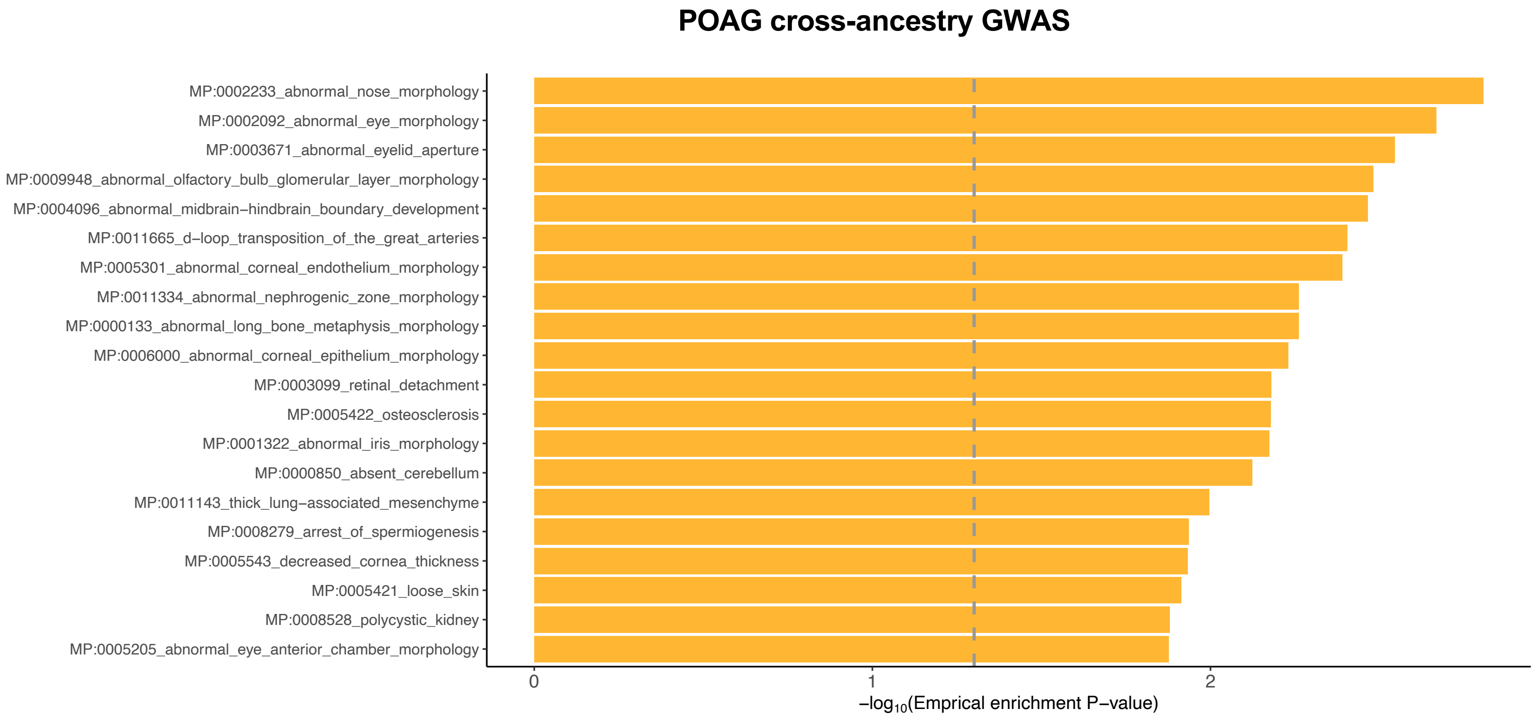

**Supplementary Figure 14. Top gene sets enriched for e/sGenes that colocalized with POAG and IOP GWAS loci.**

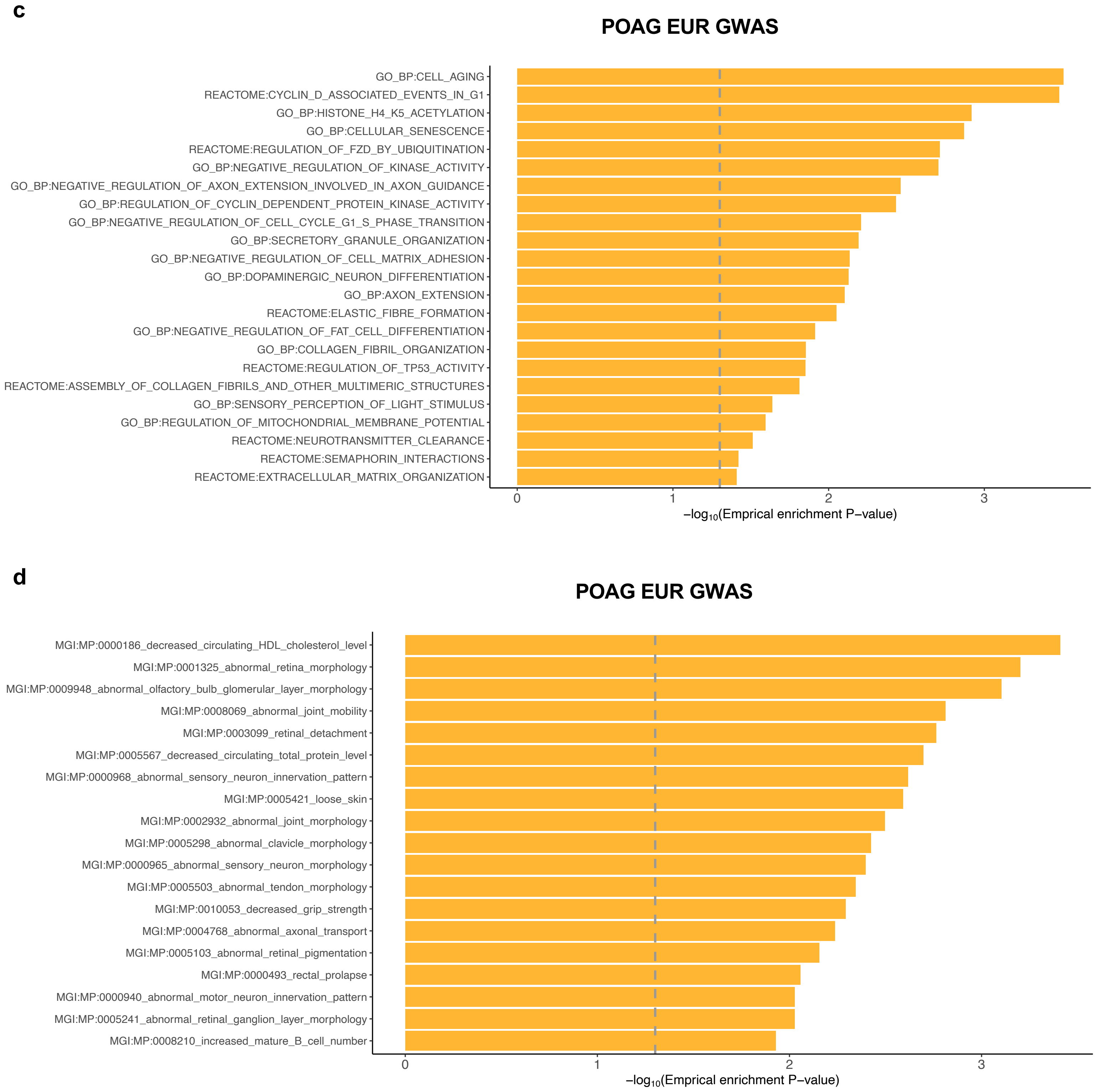

**Supplementary Figure 14. Top gene sets enriched for e/sGenes that colocalized with POAG and IOP GWAS loci.**

**e**

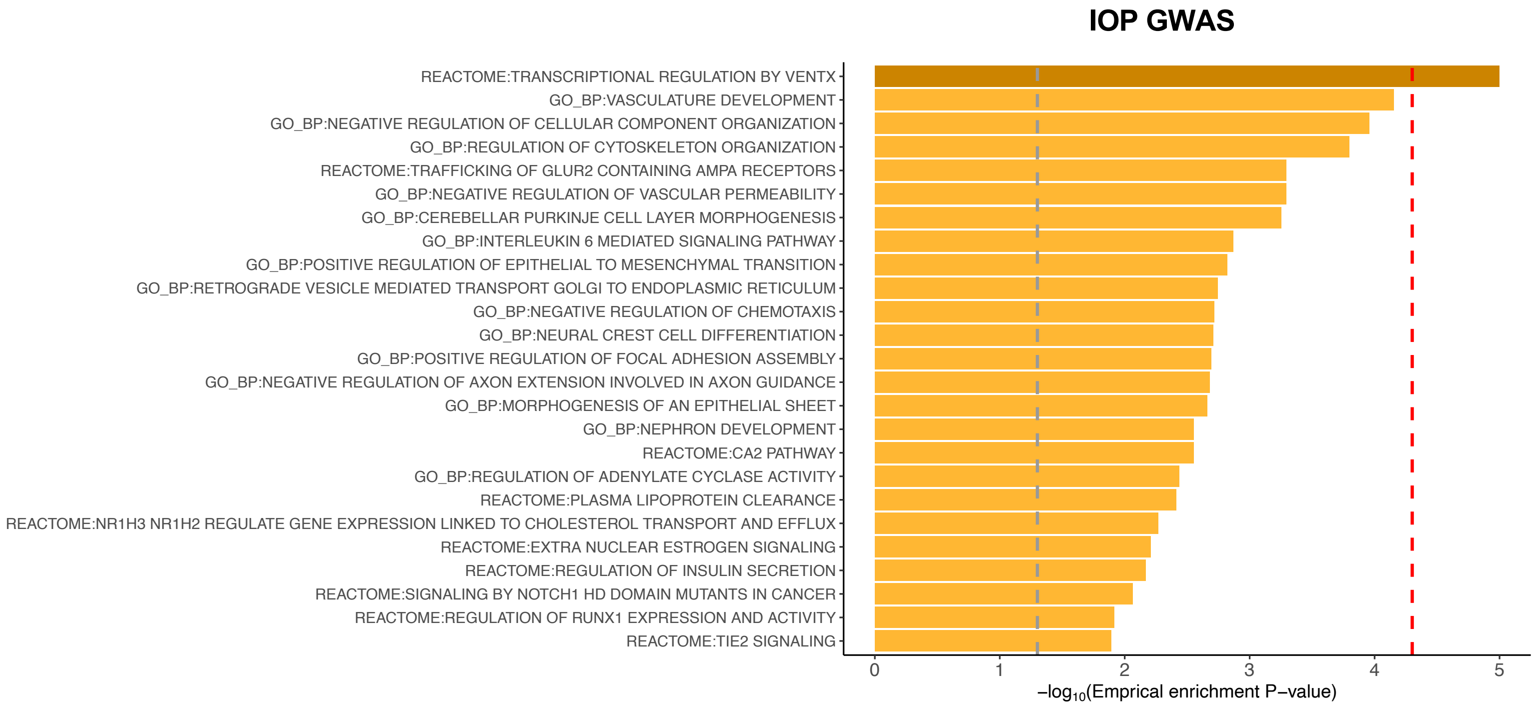


**f**

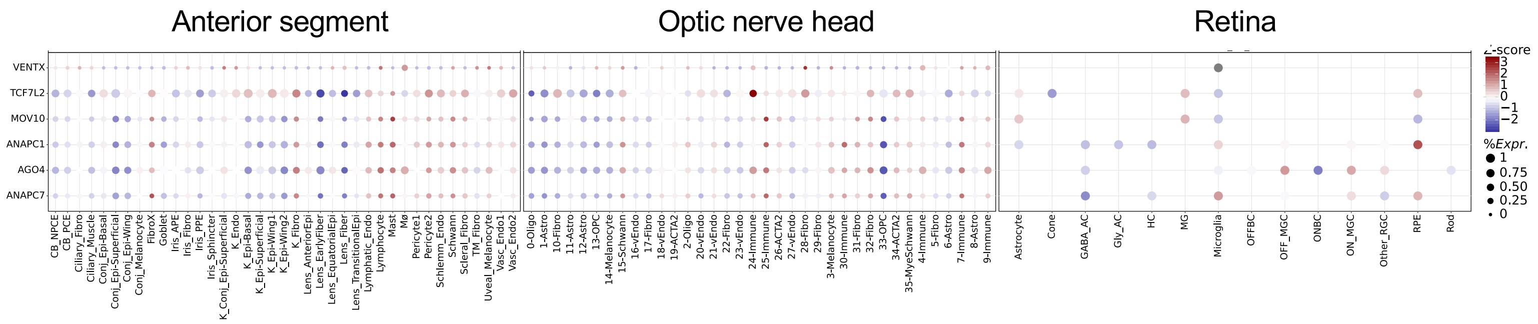


**g**

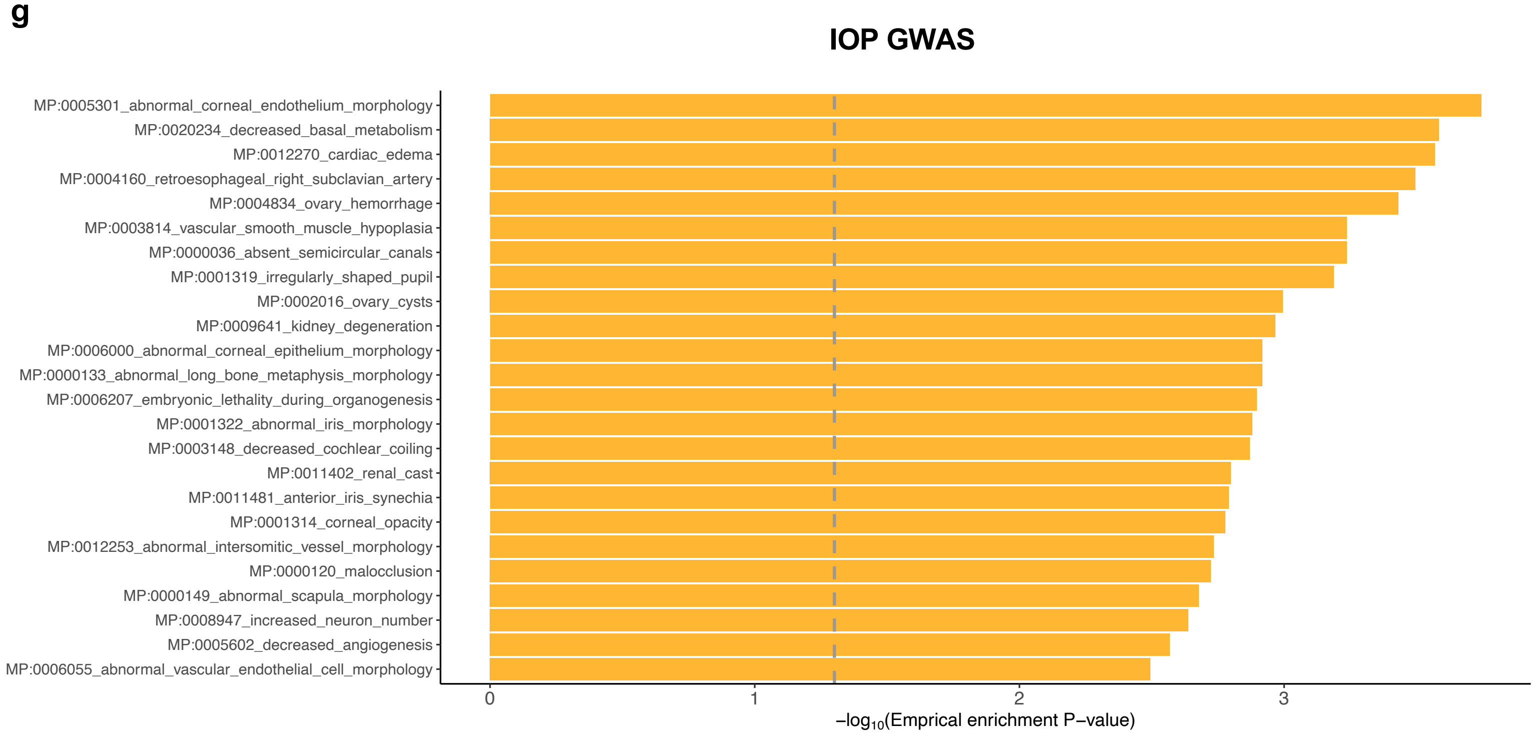

**Supplementary Figure 14. Top gene sets enriched for e/sGenes that colocalized with POAG and IOP GWAS loci.** Barplots of top ranked gene sets enriched for target genes of colocalizing e/sQTLs with POAG Cross-ancestry (**a,b**), POAG European ancestry subset (**c,d**), and IOP (**e,g**) GWAS loci are shown for biological processes (Reactome and gene ontology) (**a,c,e**) and mouse phenotype ontology (**b,d,g**) gene sets separately. An empirical gene set enrichment p-value computed with *GeneEnrich* is shown on $-\log_{10}$ scale. Dark orange bars indicate gene sets that pass Bonferroni correction (Emp. $P < 2\times10^{-5}$; red dashed line), and yellow bars indicate gene sets with a nominal Emp. $P < 0.05$ (dashed grey line). **f,** Bubble map displaying the expression of VENTX and five VENTX target genes, whose e/sQTLs colocalized with IOP and were enriched in the regulation by VENTX gene set (shown in panel **e**), across all cell types in the anterior segment, optic nerve head, and retina. The colorbar represents z-scores computed by comparing each gene's average expression in a given cell type to its per cell type average expression across all types divided by the standard deviation of all cell type expression averages. Bubble size is proportional to the percentage of cells expressing the given gene (log(TPK+1)>1). Cell type abbreviations are described in Supplementary Table 38.

**Supplementary Figure 15. Cell type enrichment of e/sQTL-mapped genes in POAG and IOP GWAS loci in the anterior segment.**

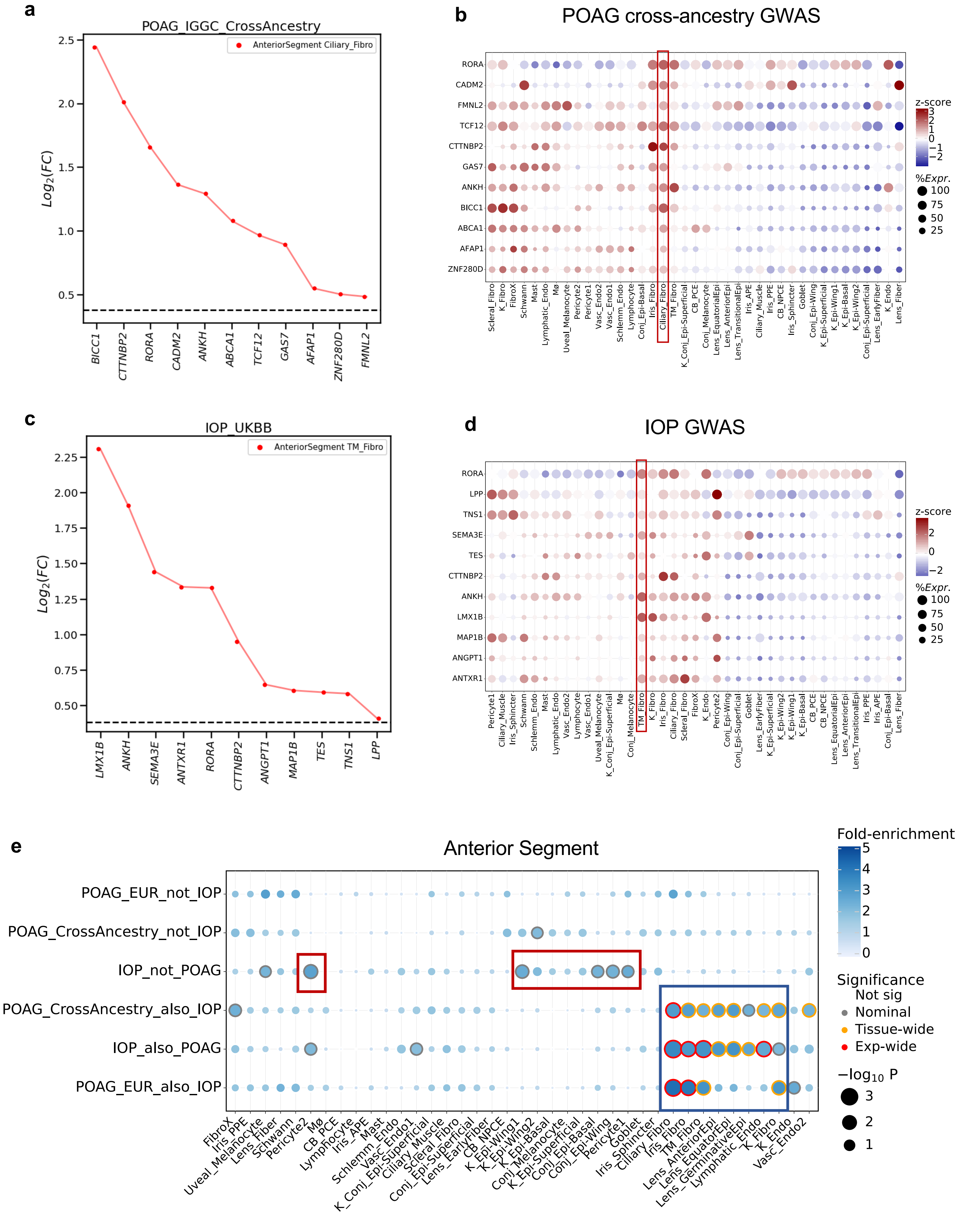

**Supplementary Figure 15. Cell type enrichment of e/sQTL-mapped genes in POAG and IOP GWAS loci in the anterior segment. a,** Differential expression ($log_2$(Fold-change), y axis) of the e/sGenes mapped to POAG cross-ancestry GWAS loci based on colocalization analysis, which are driving the enrichment signal in ciliary fibroblasts (P=0.01) in the anterior segment compared to all other cell types in the anterior segment. Horizontal dashed line represents $log_2$(Fold-change) of 0.375 (FC=1.3) and FDR<0.1 that was used as the cell type-specificity enrichment cutoff. **b**, Bubble map displaying the expression of the e/sGenes driving the POAG enrichment in ciliary fibroblasts across all cell types in the anterior segment. Colorbar represents gene expression z-scores computed by subtracting each gene's average across all cell types from its average expression in a given cell type, divided by the standard deviation of the gene's average expression across all cell types. Bubble size is proportional to the percentage of cells expressing the given gene (log(TPK+1)>1). Cell type abbreviations are described in Supplementary Table 38. Ciliary fibroblasts displayed the most significant enrichment of all cell types in the anterior segment for POAG cross-ancestry GWAS. **c,** Differential expression ($log_2$(Fold-change)) of the e/sGenes mapped to IOP GWAS loci in Trabecular meshwork (TM) fibroblasts compared to all other cell types in the anterior segment. Specifications similar to panel **a**. **d**, Bubble map displaying the expression of the e/sGenes driving the IOP enrichment in TM fibroblasts across all cell types in the anterior segment. Specifications similar to panel **b**. **e,** Significance (circle size, -log10(P-value)) and fold-enrichment (circle color) of the cell type specificity of GWAS locus sets for POAG and IOP independent and shared loci, based on ECLIPSER, for all cell types in the anterior segment. Traits (rows) and cell types (columns) were clustered based on hierarchical clustering of the euclidean distance between GWAS locus set cell type-specificity enrichment scores. Red boxes point to cell type enrichment in IOP only loci. Blue boxes point to cell type enrichment in POAG and IOP shared loci.

**Supplementary Figure 16. Cell type enrichment of e/sQTL-colocalizing genes in POAG and IOP GWAS loci in two separate retina snRNA-seq studies.**

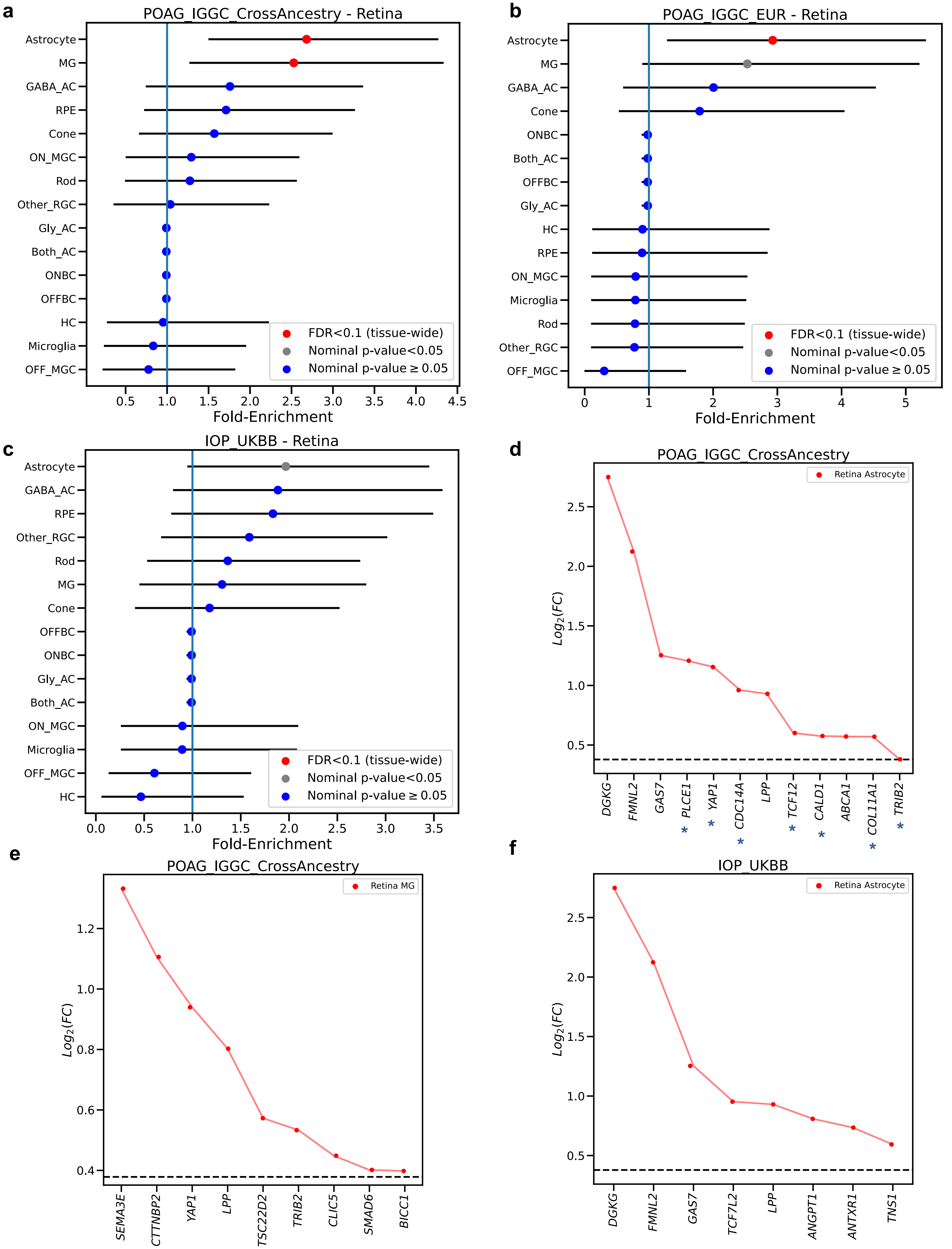

**g**

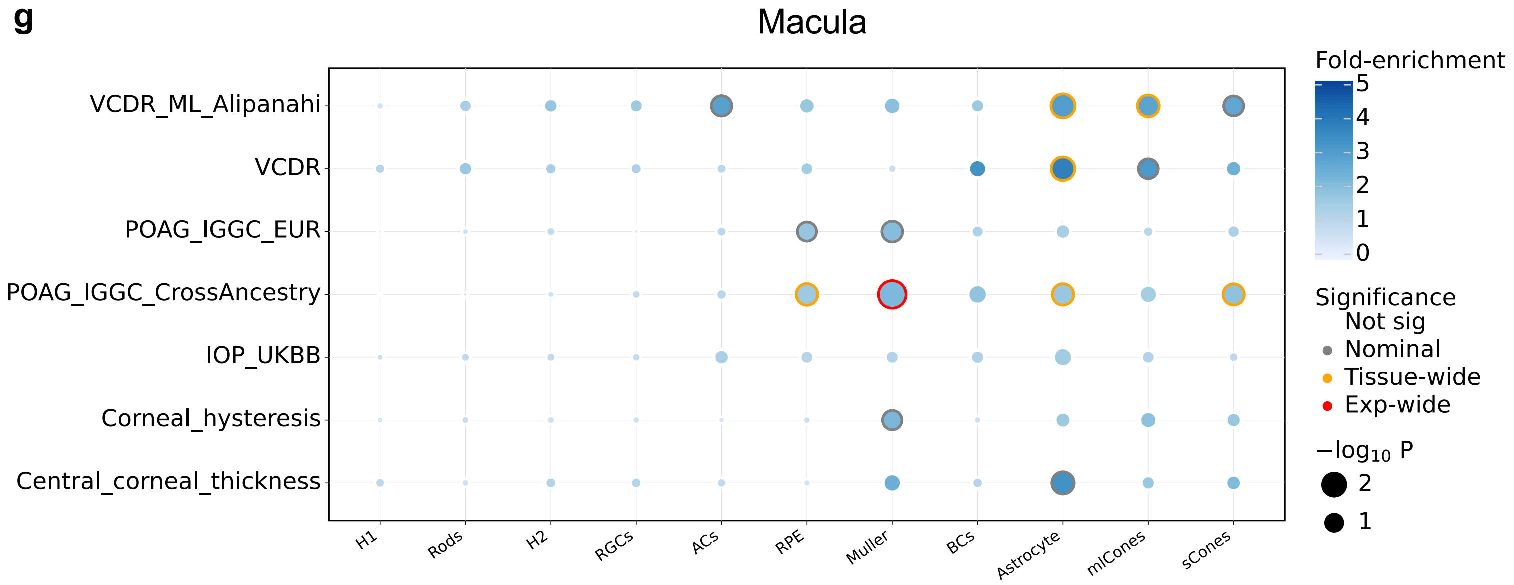


**Supplementary Figure 16. Cell type enrichment of e/sQTL-colocalizing genes in POAG and IOP GWAS loci in two separate retina snRNA-seq studies. a-c,** Cell type specificity fold-enrichment (x-axis) based on ECLIPSER in retina cell types ranked in descending order for the POAG cross-ancestry (**a**), POAG European subset (**b**), and IOP (**c**) GWAS locus sets. Red: tissue-wide significant (FDR<0.1); Grey: nominal significant (P<0.05); Blue: non-significant (P≥0.05). **d-f,** Differential expression ($\log_2$(Fold-change), y axis) of the GWAS colocalizing e/sGenes driving the enrichment signal in astrocytes (**d,f**) and Müller Glia cells (**e**) for POAG cross-ancestry GWAS loci **(d-e**) and IOP (**f**) compared to all other cell types in the retina. Horizontal dashed line represents $\log_2$(Fold-change) of 0.375 (FC=1.3) and FDR<0.1 that was used as the cell type-specificity enrichment cutoff. **g,** Significance (circle size, -log10(P-value)) and fold-enrichment (circle color) of the cell type specificity of GWAS locus sets for POAG cross-ancestry, POAG European subset, IOP, physician-defined Vertical-cup-to-disc ratio (VCDR), machine learning (ML)-defined ver (VCDR_ML_Alipanahi), central cornea thickness, and corneal hysteresis GWAS, based on ECLIPSER, are shown for each of the 11 cell types found in macula, a separate single nucleus RNA-seq dataset than the retina dataset in **a-f**. Traits (rows) and cell types (columns) were clustered based on hierarchical clustering of the euclidean distance between GWAS locus set cell type-specificity enrichment scores. Red rings: experiment-wide significant (Benjamini Hochberg (BH) FDR<0.1); Yellow rings: tissue-wide significant (BH FDR<0.1); Grey rings: nominal significant (P<0.05). H1, horizontal cells cluster 1; H2, horizontal cells cluster 2; RGCs, retinal ganglion cells; ACs, amacrine cells; RPE, retinal pigment epithelium; MG, Müller Glia; BCs, bipolar cells; mlCones, M-cone and L-cone photoreceptors; sCones, S-cone photoreceptors.

**Supplementary Figure 17. Cell type enrichment of e/sQTL-colocalizing genes in POAG and IOP independent or common loci in the retina and macula.**

**a**

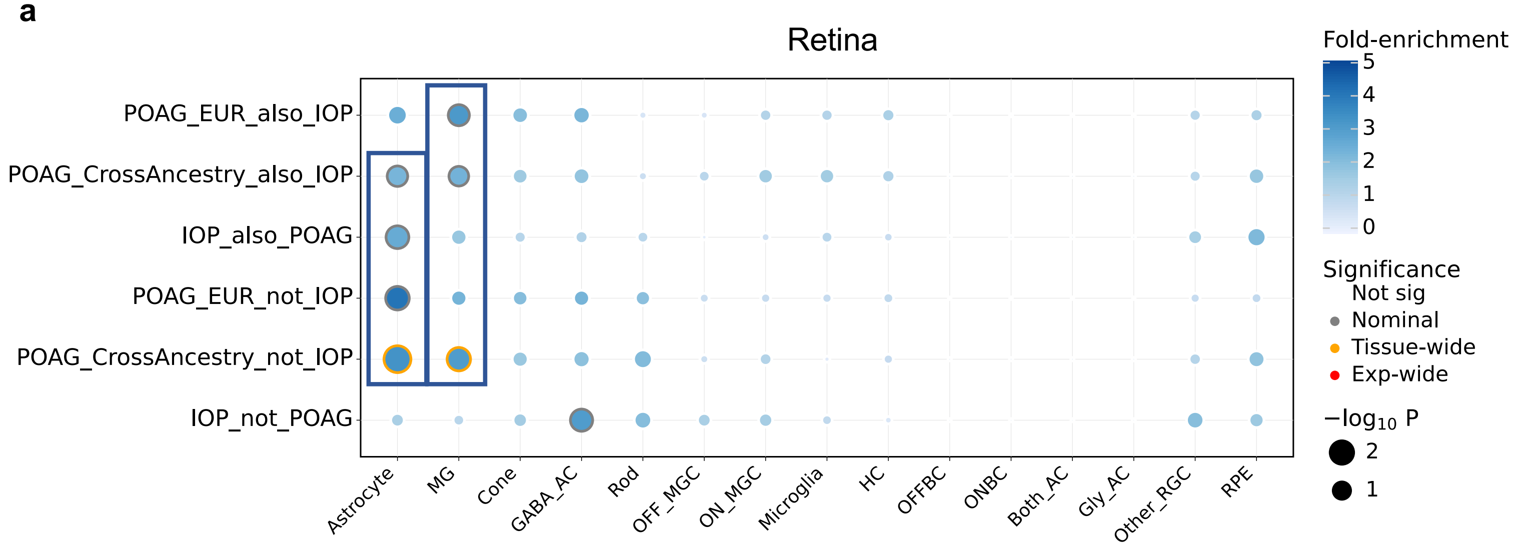


**b**

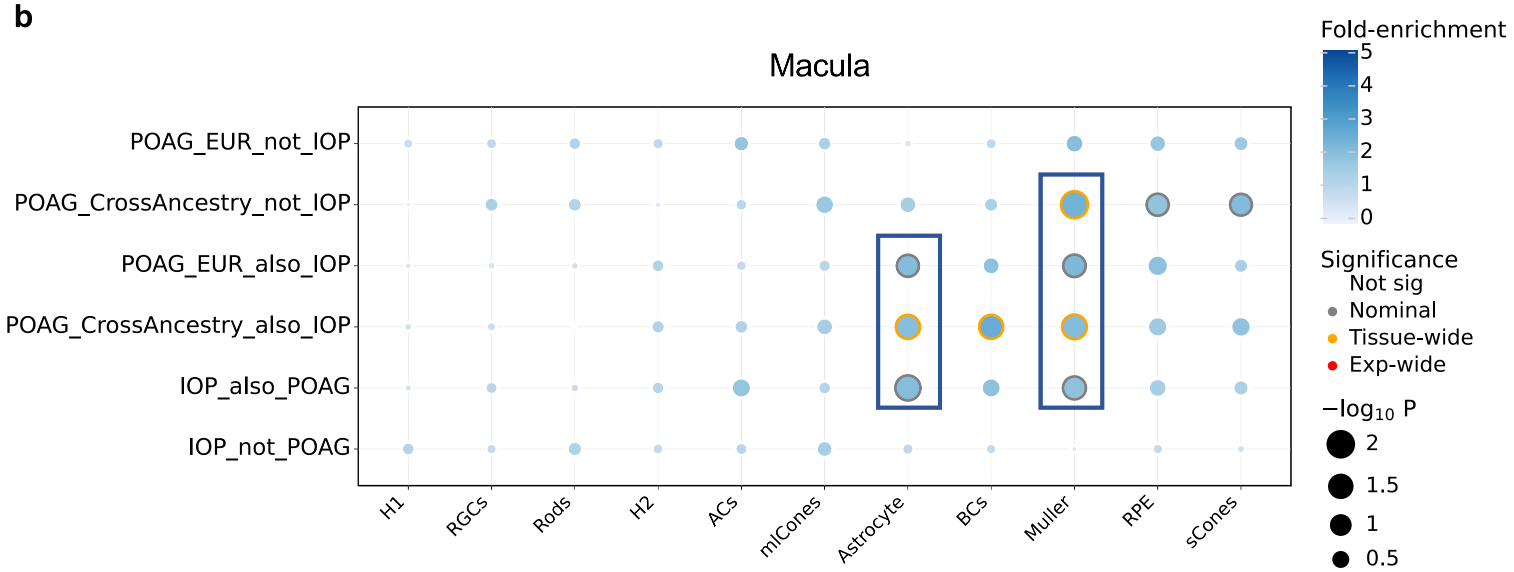


**Supplementary Figure 17. Cell type enrichment of e/sQTL-colocalizing genes with POAG and IOP independent and shared GWAS loci in retina and macula.** Significance (circle size, -log10(P-value)) and fold-enrichment (circle color) of the cell type specificity of GWAS locus sets for POAG and IOP independent and shared loci, based on ECLIPSER, for all cell types in peripheral and macular retina (**a**) and in separate macula (**b**) snRNA-seq studies. Traits (rows) and cell types (columns) were clustered based on hierarchical clustering of the euclidean distance between GWAS locus set cell type-specificity enrichment scores. Blue boxes point to cell type enrichment in POAG only or POAG and IOP shared loci. GABA_AC, GABAergic amacrine cells; Gly_AC, Glycinergic amacrine cells; Both_AC, amacrine cells with both glycinergic and GABAergic markers; ONBC, ON-bipolar cells; OFFBC, ON-bipolar cells; RPE, retinal pigment epithelium; ON_MGC, ON-midget ganglion cells; OFF_MGC, OFF-midget ganglion cells; Other_RGC, All the retinal ganglion cells (RGCs) other than the ON- and OFF-MGC (over 90% belong to these two groups); HC, horizontal cells; MG, Müller Glia. H1, horizontal cells cluster 1; H2, horizontal cells cluster 2; RGCs, retinal ganglion cells; ACs, amacrine cells; Muller, Müller Glia; BCs, bipolar cells; mlCones, M-cone and L-cone photoreceptors; sCones, S-cone photoreceptors.

**Supplementary Figure 18. Cell type enrichment of e/sQTL-colocalizing genes with POAG and IOP GWAS loci in optic nerve head and surrounding posterior tissues.**

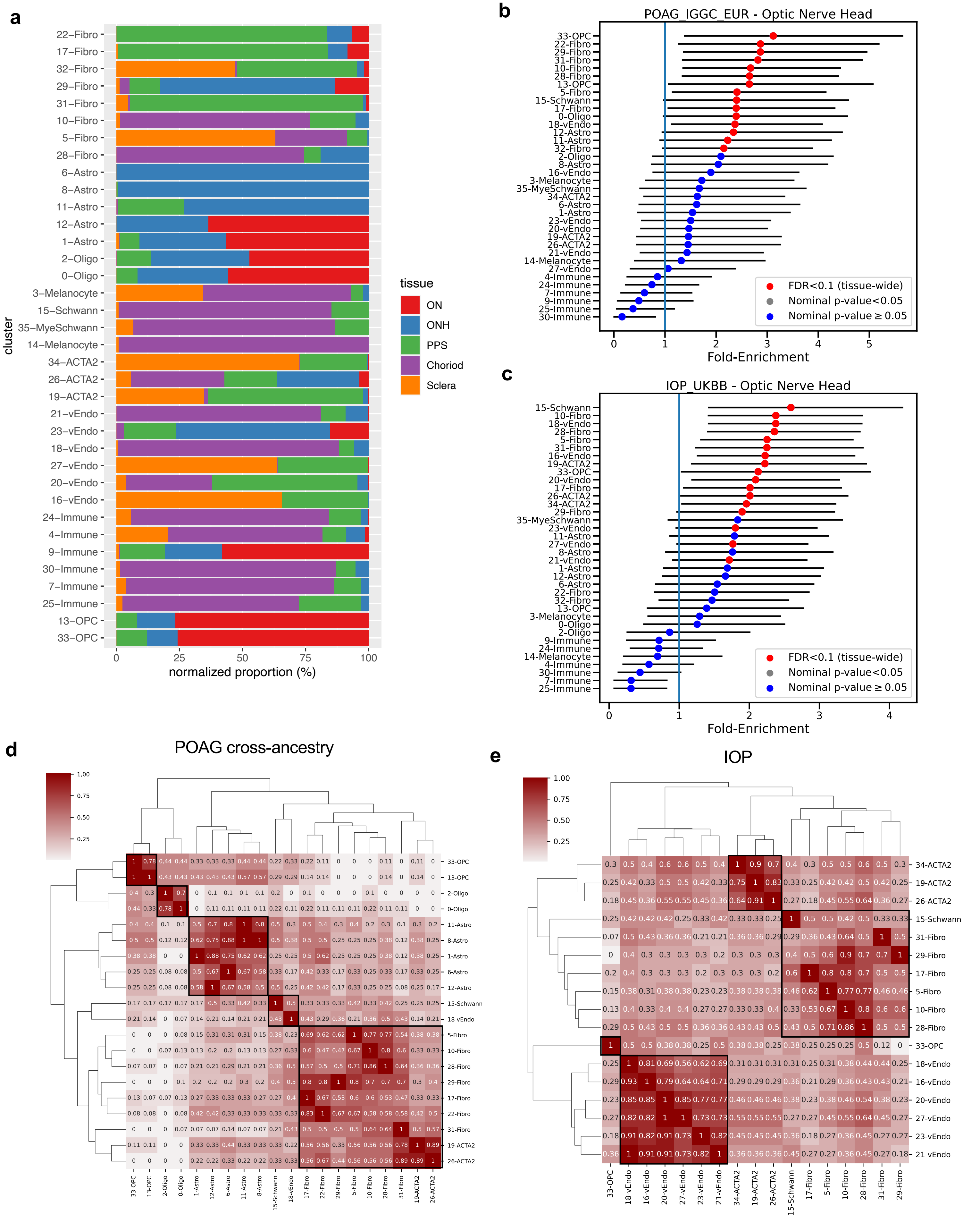

**Supplementary Figure 18. Cell type enrichment of e/sQTL-colocalizing genes with POAG and IOP GWAS loci in optic nerve head and surrounding posterior tissues.**

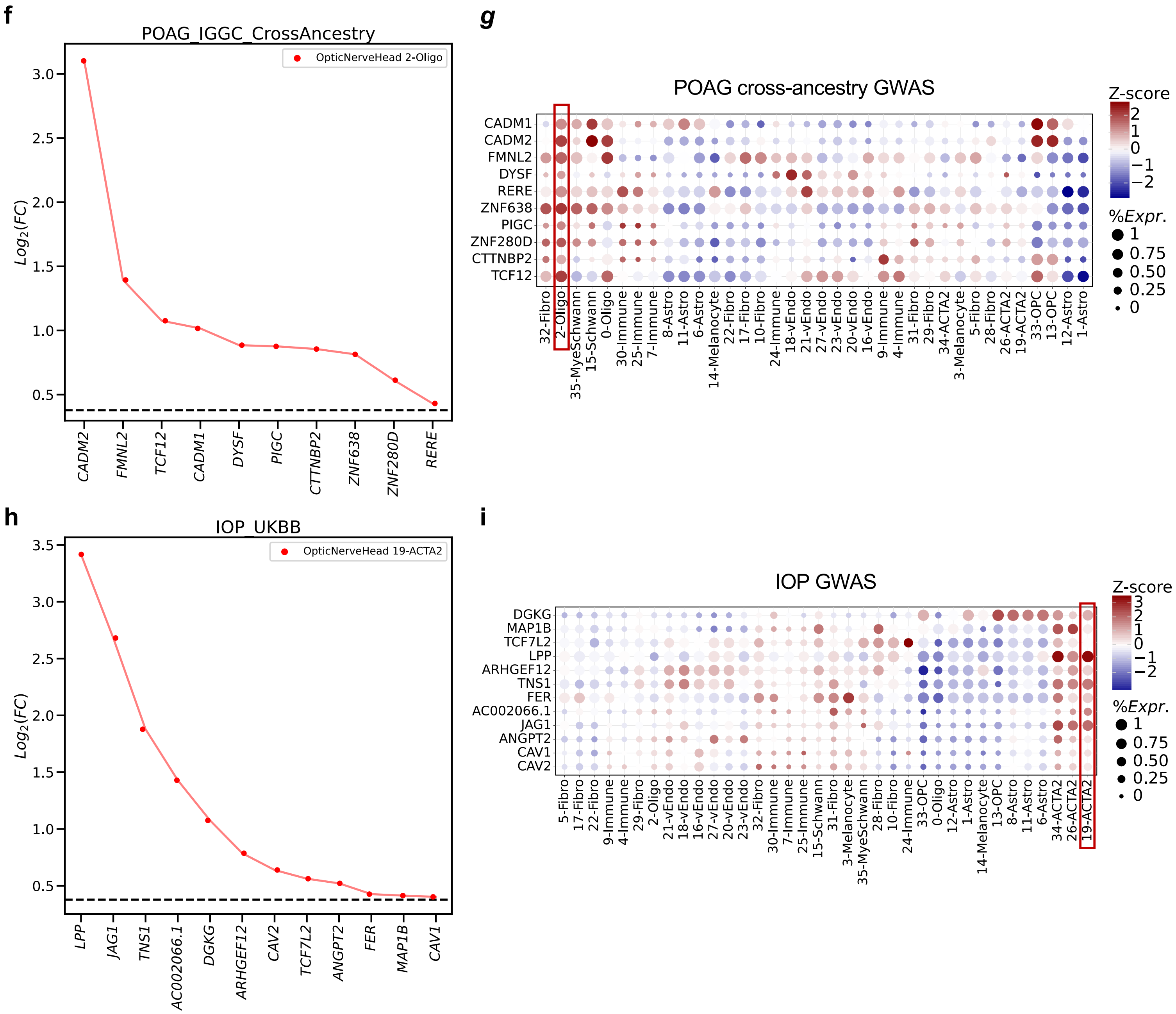

**Supplementary Figure 18. Cell type enrichment of e/sQTL-colocalizing genes with POAG and IOP GWAS loci in optic nerve head and surrounding posterior tissues. a,** Barplot displaying the proportion distribution of each cell type in the optic nerve head (ONH), optic nerve (ON), peripapillary sclera (PPS), peripheral sclera, and choroid adapted from Monavarfeshani*, Yan* *et al.,* bioRxiv 2023[37]. **b,c,** Cell type specificity fold-enrichment (x-axis) based on ECLIPSER in the ONH and surrounding posterior tissue cell types ranked in descending order for the POAG European (**b**) and IOP (**c**) GWAS locus sets. Red: tissue-wide significant (FDR<0.1); Grey: nominal significant (P<0.05); Blue: non-significant (P≥0.05). **d,e,** Heatmap of fraction of genes that overlap between the e/sGenes driving the enrichment signal for top ranked cell types (P<0.05) in the posterior tissues for POAG cross-ancestry (**d**) and IOP (**e**) GWAS loci. Numbers refer to the fraction of e/sGenes driving the cell type enrichment on each row that overlaps with the genes driving the cell type enrichment on the corresponding column. Hierarchical clustering was performed on both rows and columns using the euclidean distance between fractions. **f,h,** Differential gene expression ($\log_2$(Fold-change), y axis) in an enriched cell type compared to all other cell types for the set of genes (x axis) driving the enrichment signal of POAG cross-ancestry GWAS loci in oligodendrocytes in ONH and ON **(f)** and IOP GWAS loci in vascular smooth muscle cells (19-ACTA2) **(h).** Horizontal dashed line represents $\log_2$(Fold-change) of 0.375 (FC=1.3) and FDR<0.1 that was used as the cell type-specificity enrichment cutoff. **g,i,** Bubble maps displaying the expression of the e/sGenes driving the POAG gene enrichment in oligodendrocytes **(g)** or IOP gene enrichment in in vascular smooth muscle cells **(i)** across all cell types in the ONH and adjacent posterior tissues. The colorbar represents gene expression z-scores computed by comparing each gene's average expression in a given cell type to its per cell type average expression across all types divided by the standard deviation of all cell type expression averages. Bubble size is proportional to the percentage of cells expressing the given gene (log(TPK+1)>1). OPC, oligodendrocyte precursor cells; 19-ACTA2 and 34-ACTA2, vascular smooth muscle cell types in the PPS and sclera. Cell type abbreviations are described in Supplementary Table 38.

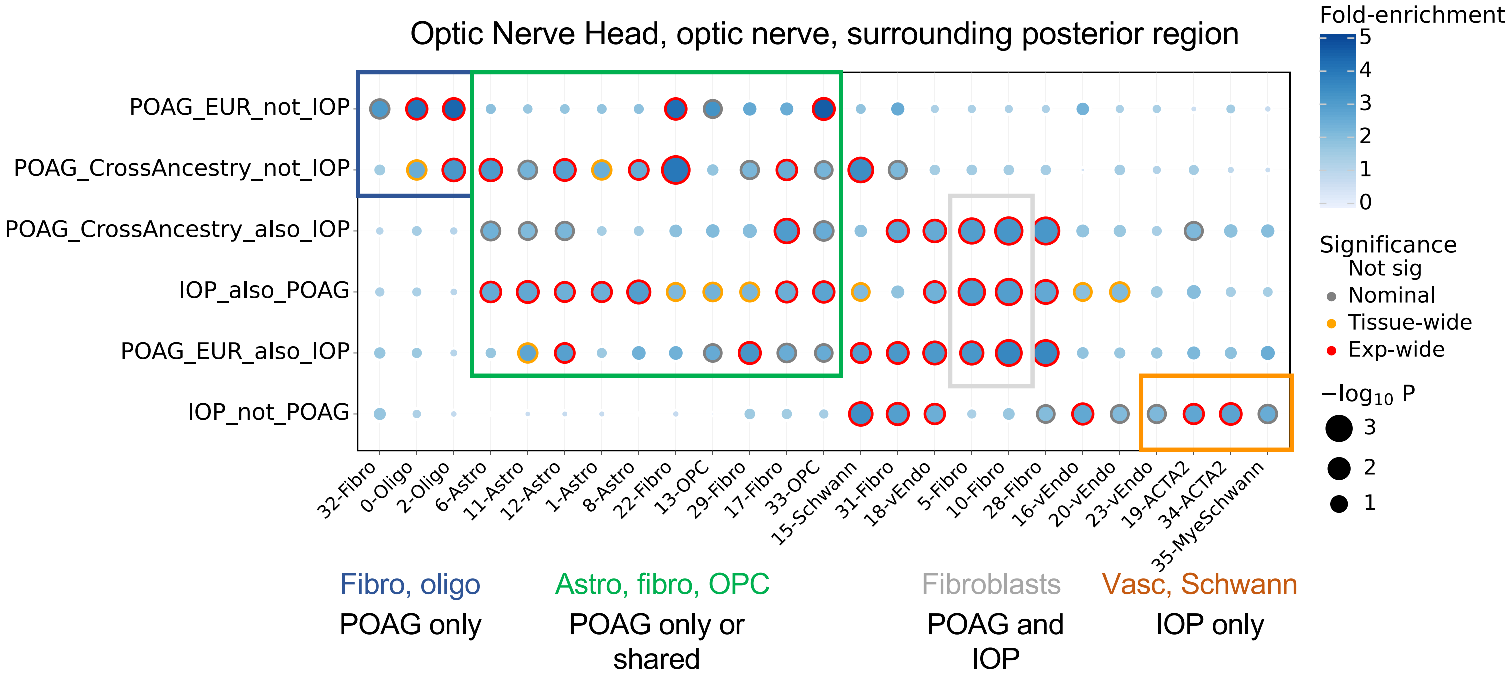


**Supplementary Figure 19. Cell type enrichment of e/sQTL-colocalizing genes with POAG and IOP independent and shared GWAS loci in optic nerve head and surrounding posterior tissues.** Significance (circle size, -log10(P-value)) and fold-enrichment (circle color) of the cell type specificity of GWAS locus sets for POAG and IOP independent and shared loci, based on ECLIPSER, for all cell types with at least one significant result in the optic nerve head, optic nerve, peripapillary sclera (PPS), peripheral sclera, and choroid. Traits (rows) and cell types (columns) were clustered based on hierarchical clustering of the euclidean distance between GWAS locus set cell type-specificity enrichment scores. OPC, oligodendrocyte precursor cells; 19-ACTA2 and 34-ACTA2, vascular smooth muscle cell types in the PPS and sclera. Cell type abbreviations are described in Supplementary Table 38.

**Supplementary Figure 20. Cell type specific expression of e/sQTL-colocalizing genes with POAG and IOP GWAS loci in glaucoma-relevant eye tissues.**

**a**

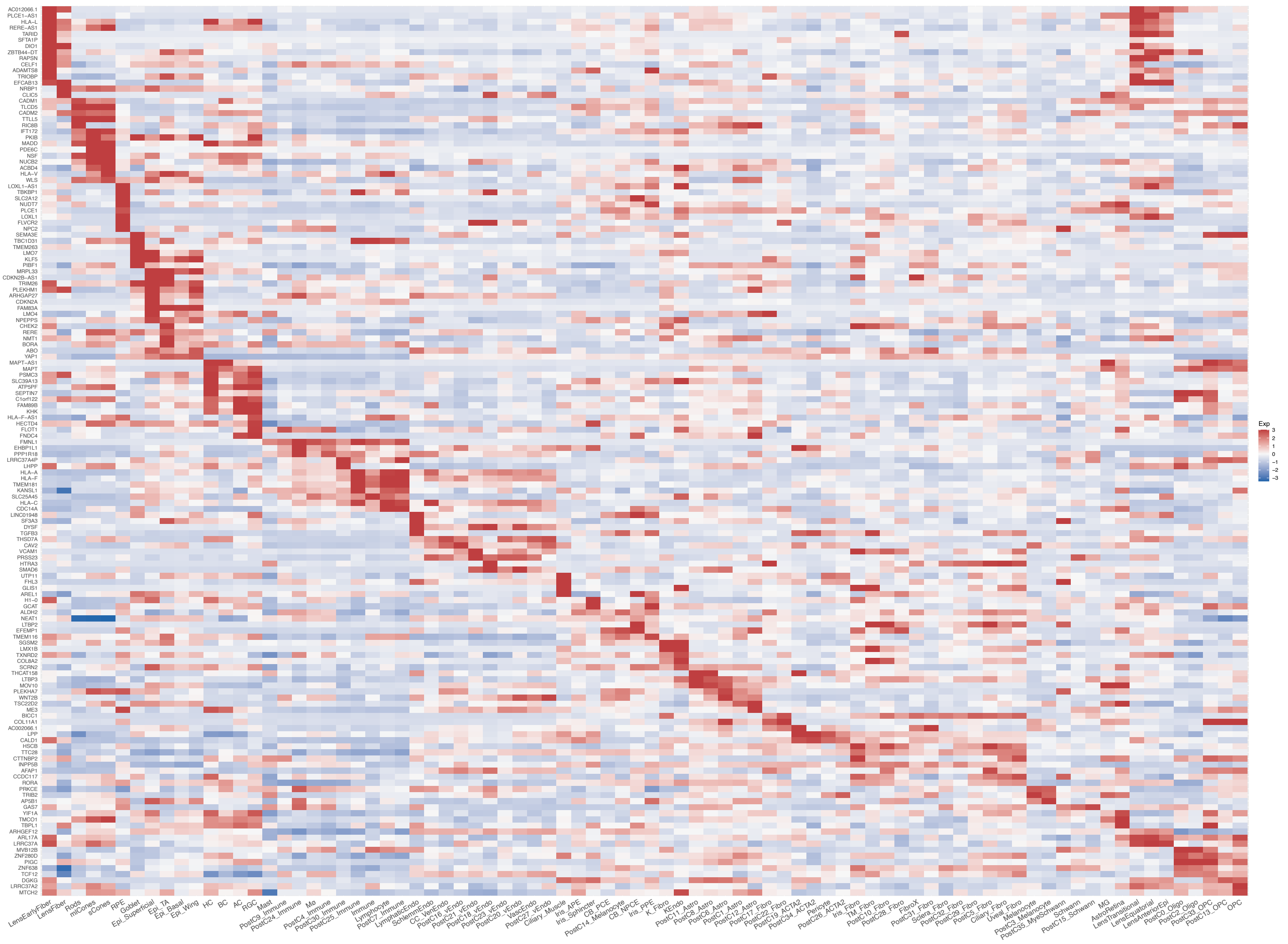

**Supplementary Figure 20. Cell type specific expression of e/sQTL-colocalizing genes with POAG and IOP GWAS loci in glaucoma-relevant eye tissues.**

**b**

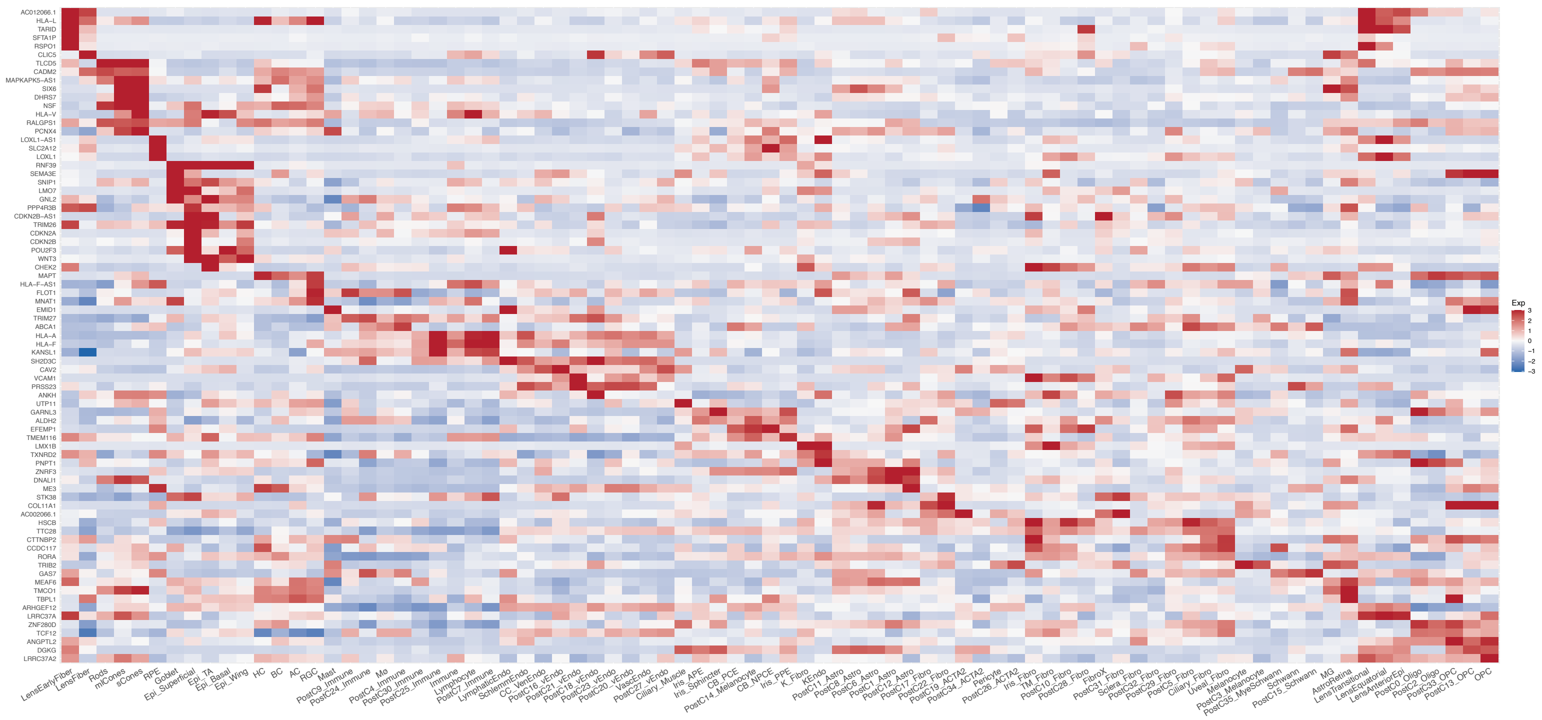

**Supplementary Figure 20. Cell type specific expression of e/sQTL-colocalizing genes with POAG and IOP GWAS loci in glaucoma-relevant eye tissues.**

**c**

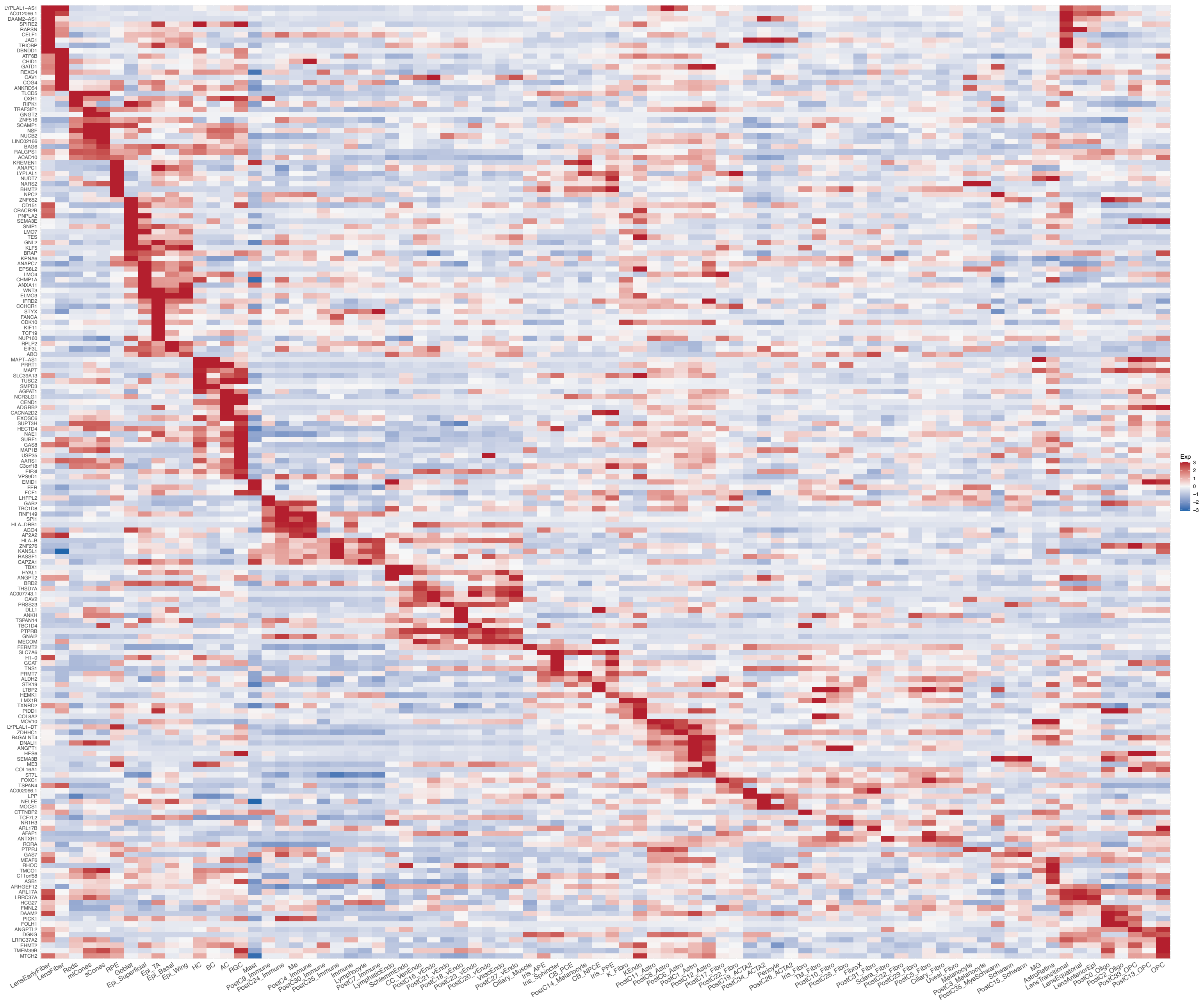

**Supplementary Figure 20. Cell type specific expression of e/sQTL-colocalizing genes with POAG and IOP GWAS loci in glaucoma-relevant eye tissues. a-c**, Heatmap displaying scaled mean expression (z-scores) of the genes whose e/sQTLs significantly colocalized with POAG cross-ancestry (**a**), POAG European subset (**b**) and IOP (**c**) GWAS loci across anterior segment, retina, and optic nerve head/optic nerve/peripapillary sclera/sclera/choroid (Post) cell types. Only genes expressed in more than 10% of cells in any cell type are shown (146, 77 and 179 genes for POAG cross-ancestry, POAG EUR and IOP loci, respectively). Colorbar represents gene expression z-scores computed by subtracting each gene's average across all cell types from its average expression in a given cell type, divided by the standard deviation of the gene's average expression across all cell types. Hierarchical clustering was performed on both rows (genes) and columns using the Euclidean distance between z-score vectors. Some of these data are replotted from Figure S6B in Monavarfeshani *, Yan* *et al.,* bioRxiv 2023.

**Supplementary Figure 21. Cell type specific enrichment of e/sQTL-mapped genes to negative control trait loci in anterior and posterior eye tissues.**

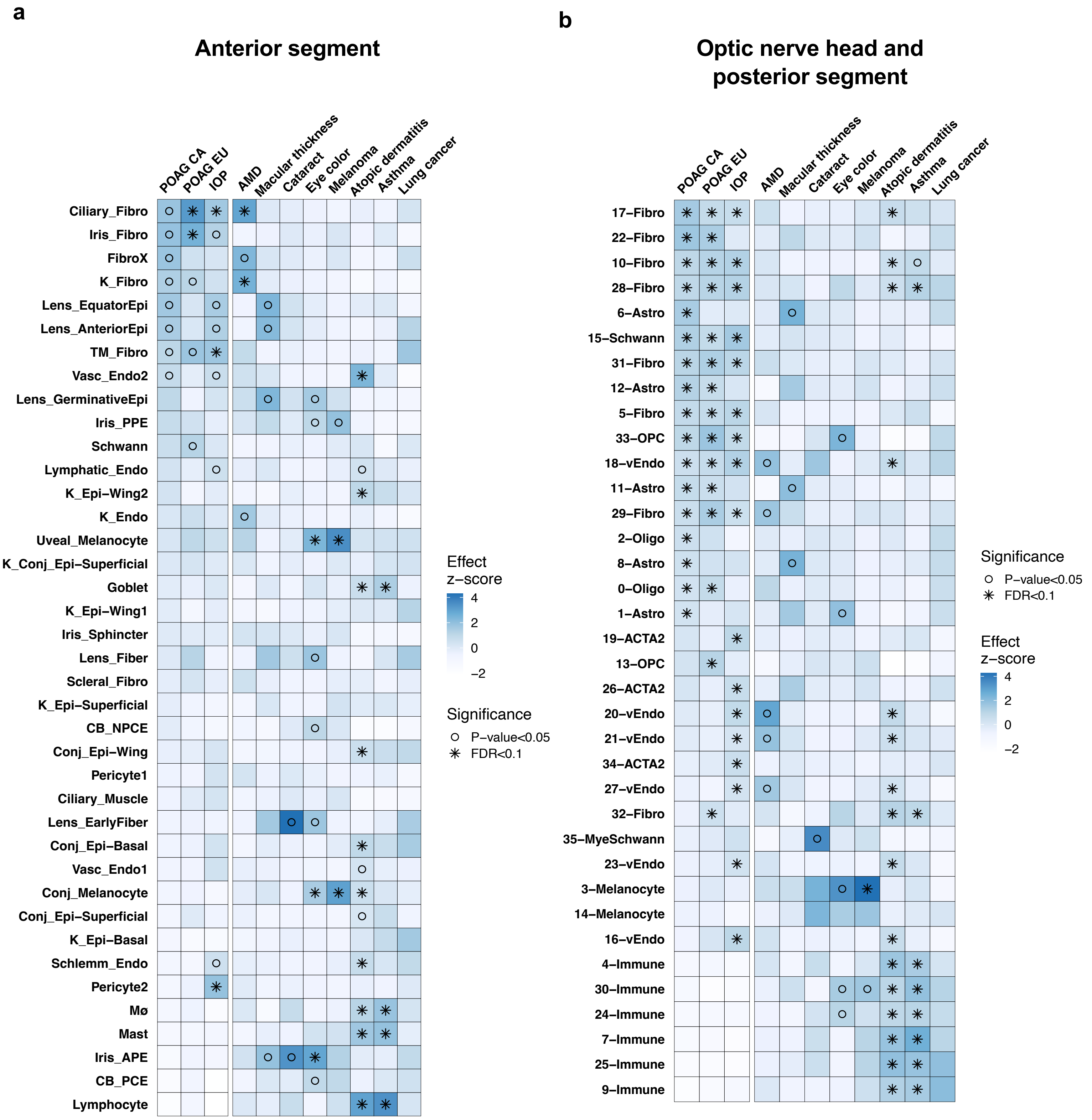

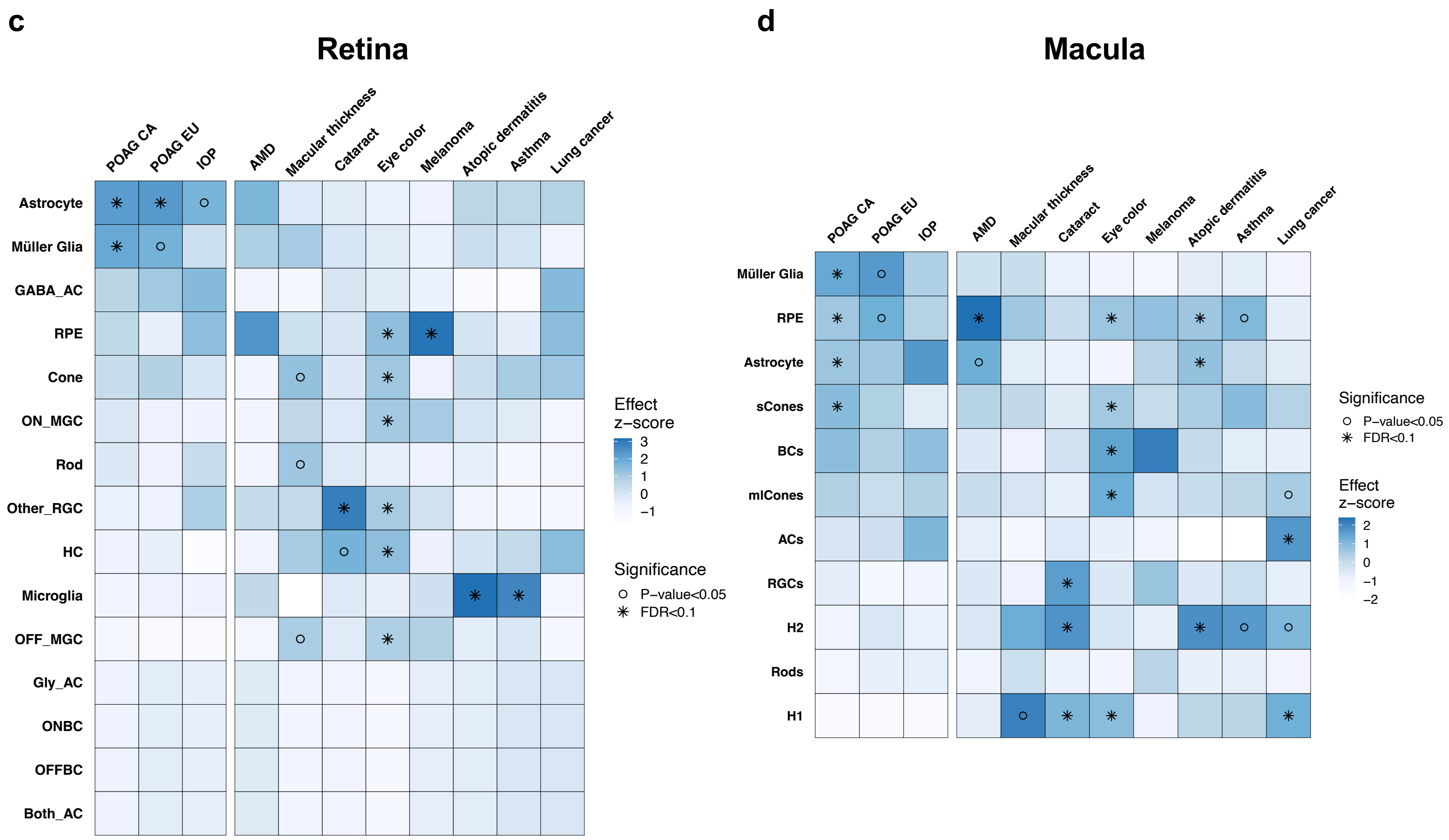


**Supplementary Figure 21. Cell type specific enrichment of e/sQTL-mapped genes to negative control trait loci in eye tissues. a-d,** Heatmaps displaying scaled ECLIPSER effect size (z-scores) of cell type fold-enrichment for POAG cross-ancestry (CA), POAG European subset (EU), and IOP GWAS loci, followed by eight negative control traits using four single nucleus RNA-sequencing datasets: the anterior segment **(a),** optic nerve head, optic nerve and surrounding posterior tissues **(b),** peripheral and macular retina **(c),** and macula **(d).** The blue colormap is proportional to the effect z-score. Rows are ordered by the POAG CA effect sizes per cell, sorted in descending order. Asterisks represent significant cell types at Benjamini-Hochberg FDR below 0.1, and circles represent nominal significance (P<0.05). Genes were mapped to POAG and IOP GWAS loci based on colocalization analysis, and to the negative control traits based on the target genes of GTEx and retina e/sQTLs that are in linkage disequilibrium (LD; r2>0.8) to the lead GWAS variants. Cell type abbreviations are described in Supplementary Table 38. AMD, age-related macular degeneration.

**Supplementary Figure 22. Cell type specific enrichment of POAG and IOP associations in anterior and posterior eye tissues based on three different methods.**

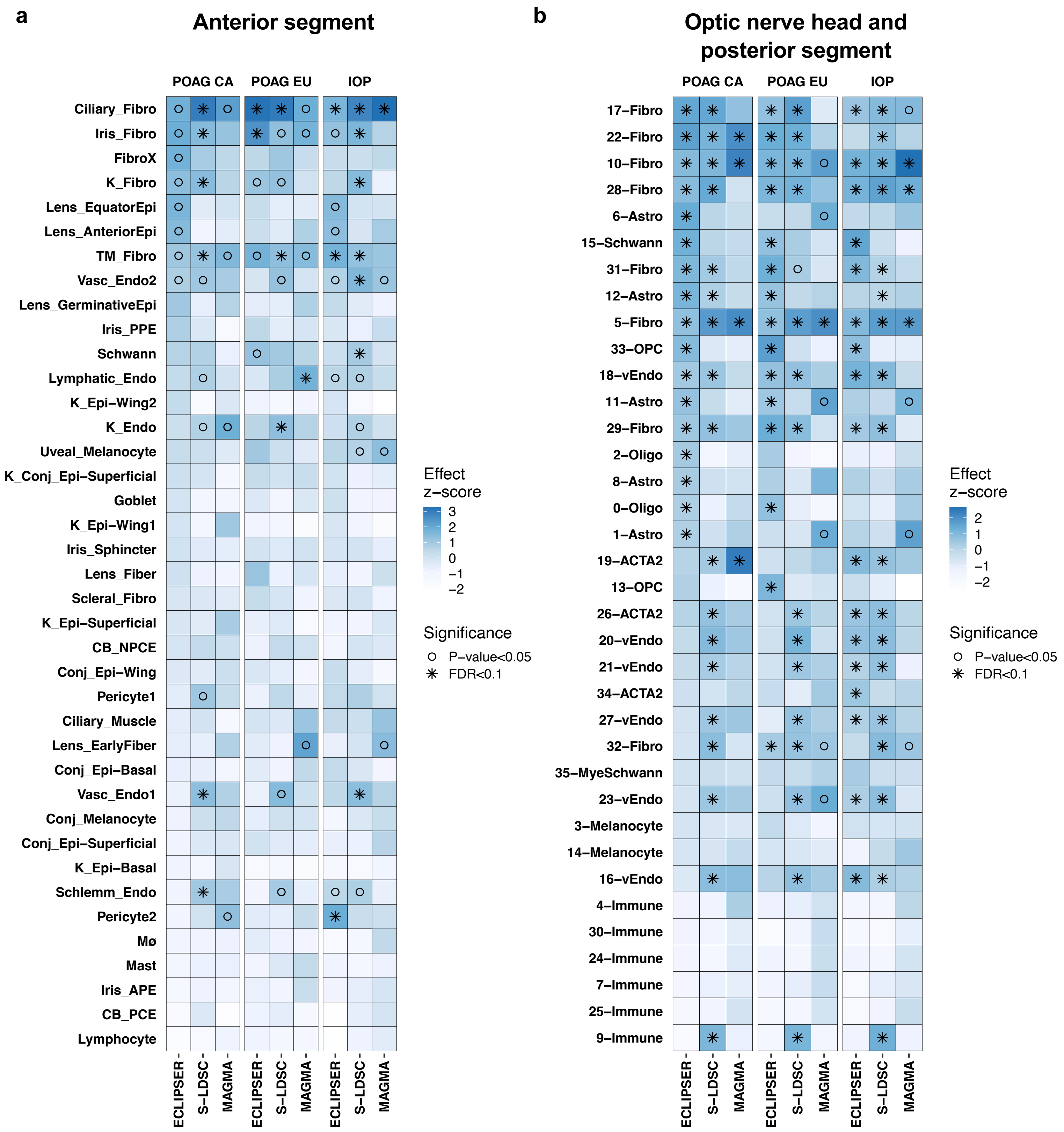

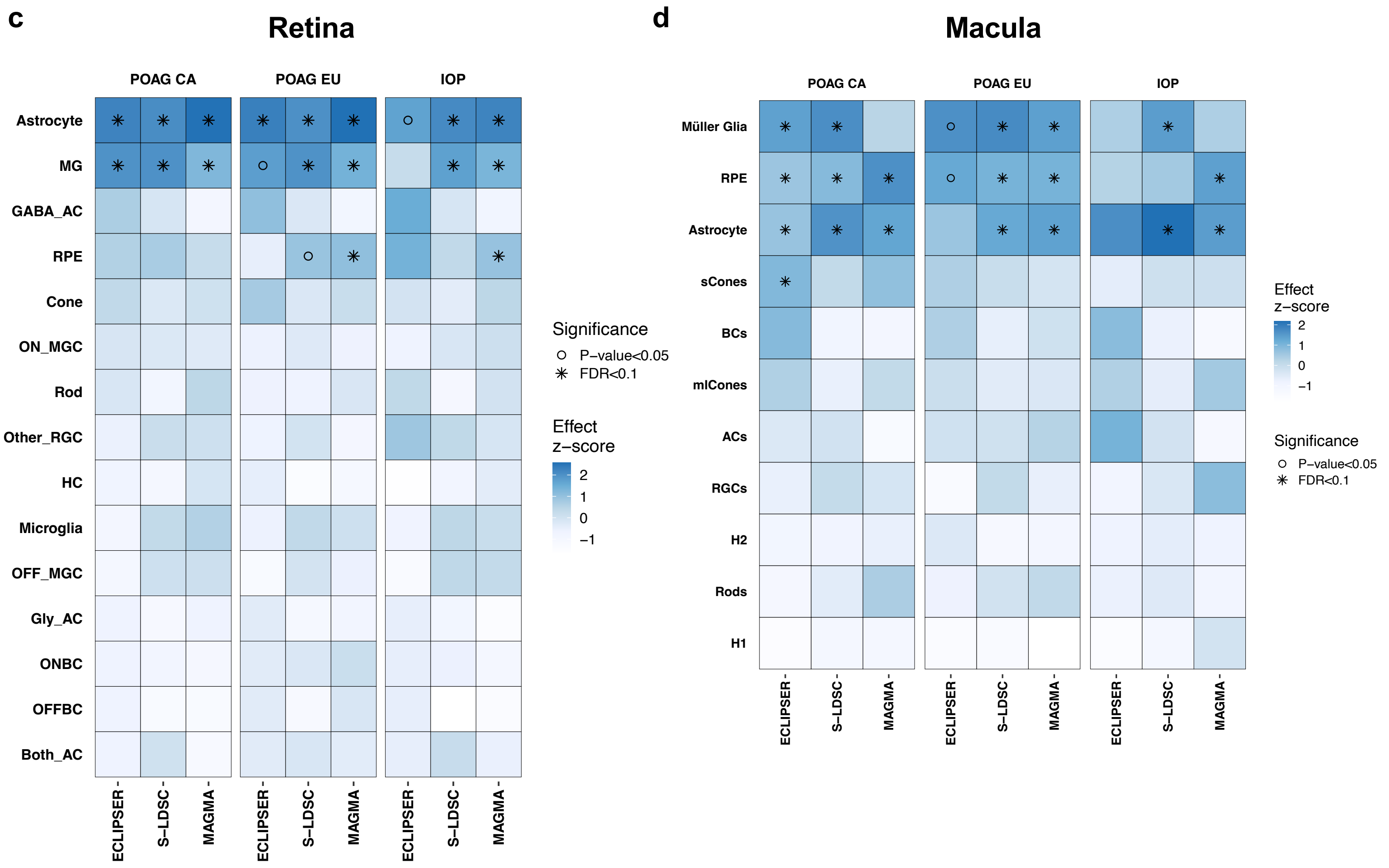


**Supplementary Figure 22. Cell type specific enrichment of POAG and IOP associations in anterior and posterior eye tissues based on three different methods.** Heatmaps displaying scaled cell type enrichment effect sizes (z-scores) of POAG cross-ancestry (CA), POAG European subset (EU), and IOP GWAS associations, based on ECLIPSER, stratified LD score regression (S-LDSC) and MAGMA methods for four single nucleus RNA-sequencing datasets: the anterior segment **(a),** optic nerve head, optic nerve and surrounding posterior tissues **(b),** peripheral and macular retina **(c),** and macula **(d).** The blue colormap is proportional to the effect z-score. Rows are ordered by the POAG CA effect sizes per cell with ECLIPSER, sorted in descending order. Asterisks represent significant cell types at Benjamini-Hochberg FDR below 0.1, and circles represent nominal significance ($P<0.05$). Genes were mapped to POAG and IOP GWAS loci based on colocalization analysis for the ECLIPSER analysis, and genome-wide GWAS associations were used in S-LDSC and MAGMA cell type enrichment analysis. Cell type abbreviations are described in Supplementary Table 38. LD, linkage disequilibrium.

**Supplementary Figure 23. Cell type enrichment of genes mapped to VCDR and cornea-related trait GWAS loci in glaucoma-relevant ocular tissues.**

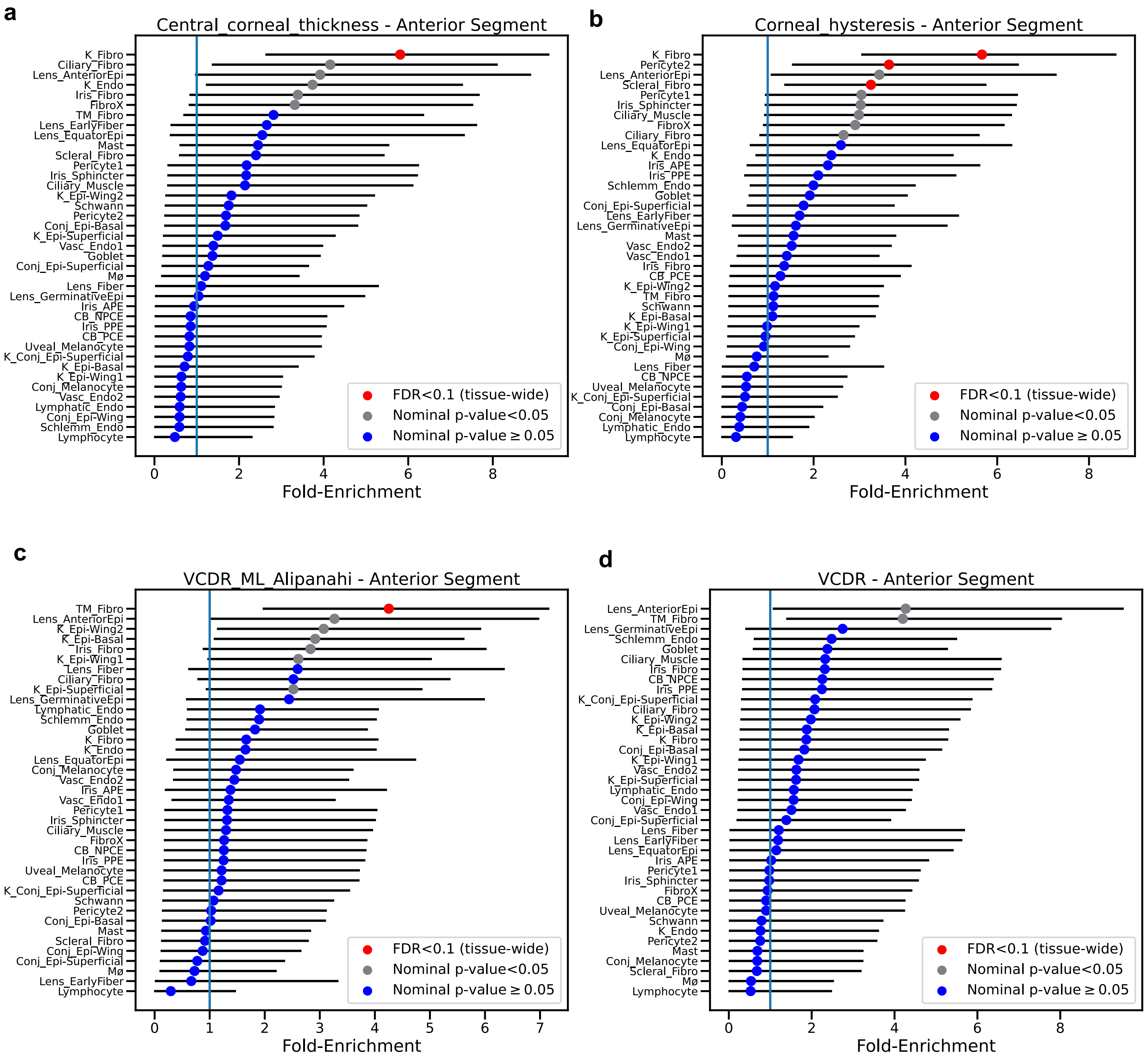

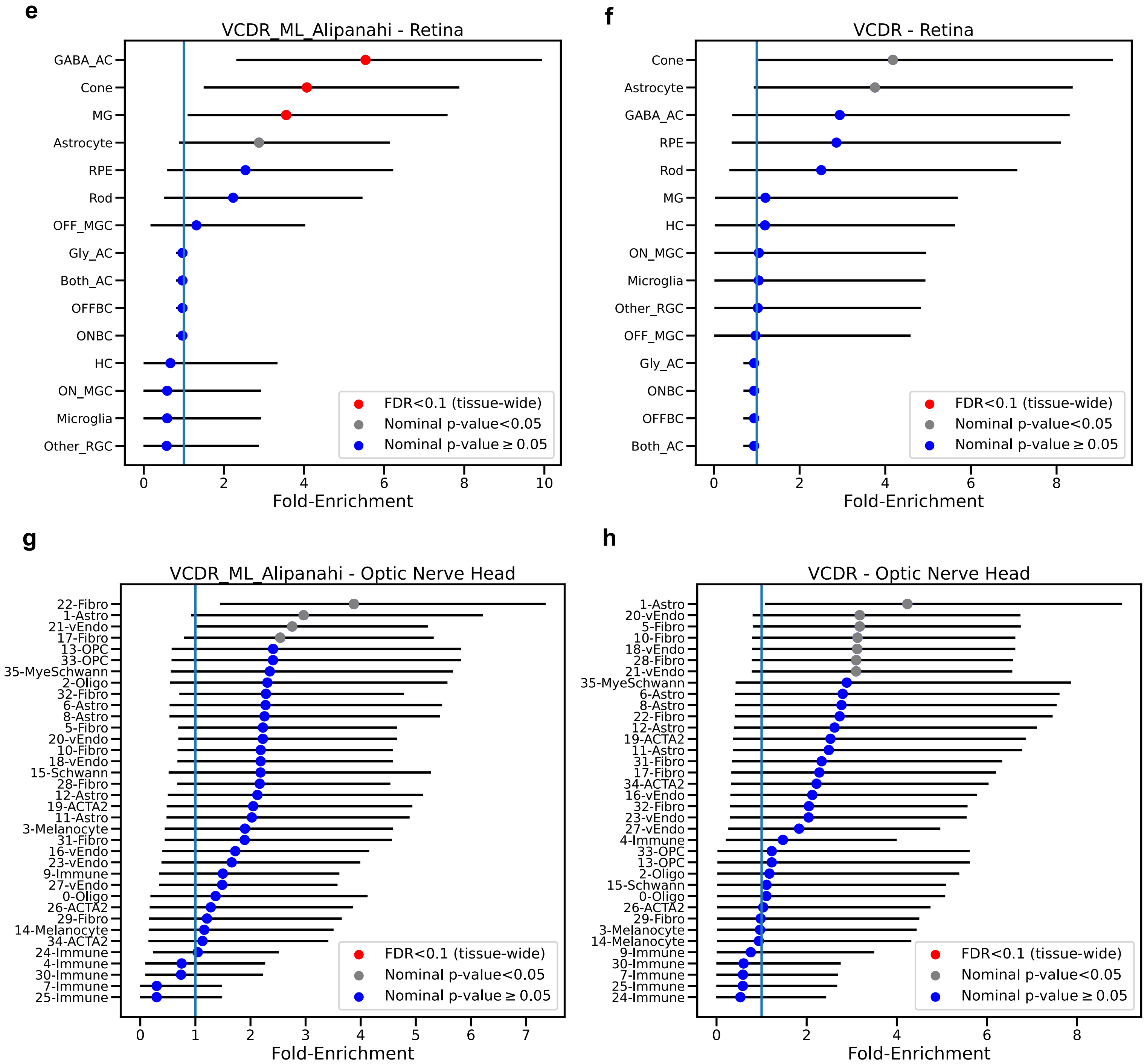


**Supplementary Figure 23. Cell type enrichment of genes mapped to VCDR and cornea-related trait GWAS loci in glaucoma-relevant ocular tissues. a-i,** Cell type specificity fold-enrichment (x-axis) based on ECLIPSER in the anterior segment (**a-d),** retina (**e-f**), and optic nerve head and surrounding posterior tissues (**g-h**) cell types ranked in descending order for the central corneal thickness (**a**), corneal hysteresis (**b**), machine learning (ML)-defined vertical-cup-to-disc ratio (VCDR) (**c,e,g**), and physician-defined VCDR (VCDR) (**d,f,h**) GWAS locus sets. Red: tissue-wide significant (FDR<0.1); Grey: nominal significant (P<0.05); Blue: non-significant (P≥0.05). Cell type abbreviations are described in Supplementary Table 38.
